# Trends in incidence and antimicrobial resistance for five major causes of bacteraemia in a Canadian metropolitan area, 2006–22: a genomic and antimicrobial use cohort study

**DOI:** 10.64898/2026.08.27.26361471

**Authors:** Thi Mui Pham, Joshua T. Smith, Tatum D. Mortimer, Yonatan H. Grad, Ashlee M. Earl, Ian Lewis, the PRIME Consortium

**Affiliations:** Department of Epidemiology, Harvard T.H. Chan School of Public Health, 02115, Boston, MA, USA; Center for Communicable Disease Dynamics, Harvard T.H. Chan School of Public Health, 02115, Boston, MA, USA; Department of Immunology and Infectious diseases, Harvard T.H. Chan School of Public Health, 02115, Boston, MA, USA; Infectious Disease and Microbiome Program, Broad Institute of MIT & Harvard, Cambridge, MA, 02142, USA; Population Health, University of Georgia, Athens, GA, 30602, USA; Alberta Centre for Advanced Diagnostics, Department of Biological Sciences, University of Calgary, Calgary, Alberta, Canada

**Author notes:** Corresponding author: Ian Lewis, Alberta Centre for Advanced Diagnostics, Department of Biological Sciences, University of Calgary, Calgary, Alberta, Canada. These authors contributed equally to this work and are co-first authors (Thi Mui Pham, Joshua T. Smith, Tatum D. Mortimer). These authors contributed equally to this work and are co-senior authors (Yonatan H. Grad, Ashlee M. Earl, Ian Lewis).

## Abstract

**Background:** Using a population-based cohort from the Calgary Health Zone (CHZ), Canada, we integrated longitudinal antimicrobial susceptibility and prescribing data with the whole genome sequences of five major pathogens. We aimed to assess how antimicrobial resistance (AMR) responds to prescribing changes and determine which bacterial strains shape these dynamics.

**Methods:** We analysed antibiotic prescribing rates, clinical and genomic data from 7,271 *Staphylococcus aureus*, 1,609 *Enterococcus faecalis*, 801 *Enterococcus faecium*, 11,363 *Escherichia coli*, and 2,319 *Klebsiella pneumoniae* isolates, associated with bacteraemia episodes in the CHZ between 2006–2022. Genomic clusters (referred to as strains) were identified using StrainGST and assigned to known sequence types (STs) or clonal complexes (CCs). Strain-level incidence, stratified by community-onset (isolates collected ≤48h after admission) and hospital-onset (>48h after admission), AMR phenotypes, and prescribing rates were modelled using negative-binomial and binomial regression. Temporal trends were quantified using average annual percentage change (AAPC).

**Findings:** Between 2010–2022, fluoroquinolone prescribing declined in both community (AAPC=−6·8% [95% CI −8·1, −5·4]; p<0·0001) and hospital settings (AAPC=−5·1% [−6·5, −3·7]; p<0·0001). This was accompanied by a significant reduction in fluoroquinolone resistance among Gram-positive species. Specifically, *S aureus* bacteraemia resistant to clinically important antibiotics, cloxacillin, ciprofloxacin, erythromycin, and clindamycin, declined from 2006 to 2022, mostly in hospital-onset cases (AAPC=−16·0%, [−19·3%, −12·7%], p<0·0001). In *E coli,* ceftriaxone and ciprofloxacin resistance were clustered in ST131 and the emerging ST1193; the latter increased steadily, particularly in community-onset cases (AAPC=17·7%, [9·0%, 30·0%], p<0·0001). CTX-M-27-producing *E coli* ST131 strains increased (AAPC=23·8%, [17·4%, 30·5%], p<0·0001) between 2008–2022, while CTX-M-14-producing *E coli* ST131 declined (AAPC=−15·9%, [−21·3%, −10·2%], p<0·0001) between 2013–2022. These trends were paralleled by an increase in community cephalosporin prescribing (AAPC=7·3%, [4·2%, 10·5%], p<0·0001) between 2010–2022. For *K pneumoniae*, hypervirulent ST23 was most common (N=88) with an increasing trend in incidence (AAPC=2·3%, [−4·4%, 9·4%]) between 2006–2019.

**Conclusions:** The contrasting resistance trends between Gram-positive and Gram-negative species underscore the complexity of AMR control efforts. Effective strategies will require stewardship efforts targeting multiple drug classes, genomic surveillance for emerging resistant strains, and interventions extending beyond hospital settings.

**Funding:** This study was funded by a Large Scale Applied Research Project competition award (2017) from Genome Canada, administered via Genome Alberta. IAL is supported by an Alberta Innovates Translational Health Chair and a UCalgary Research Excellence Chair, with additional support from the Canada Foundation for Innovation (CFI-JELF 34986), the Natural Sciences and Engineering Research Council (DG 04547). Microbial isolates were prepared for analysis at the Alberta Centre for Advanced Diagnostics, which is supported by PrairiesCan (000022734). This work was also supported by federal funds from the US National Institute of Allergy and Infectious Diseases, National Institutes of Health, Department of Health and Human Services, under grant number U19AI110818 to the Broad Institute. JTS is additionally supported by the NIH/NIAID F32AI179151.

**Research in context:** *Evidence before this study:* We searched PubMed for articles published in English up to Dec 23, 2025, using (antibiotic[tiab] OR antimicrobial[tiab]) AND (use[tiab] OR prescribing[tiab] OR consumption[tiab]) AND (resistance[tiab] OR “multidrug resistance“[tiab]) AND (relationship[tiab] OR association[tiab] OR correlation[tiab] OR effect[tiab] OR impact[tiab] OR influence[tiab] OR regression[tiab]) AND (genomic OR genetic OR “”molecular epidemiology) AND (“Staphylococcus aureus” OR “Escherichia coli” OR “Klebsiella pneumoniae” OR “Enterococcus faecalis” OR “Enterococcus faecium”) NOT animal. This search yielded 739 articles. Numerous studies have characterised the molecular epidemiology of these species over the past two decades: For *Staphylococcus aureus*, extensive genomic surveillance has revealed that a limited number of sequence types have dominated global infections over the past two to three decades. Pandemic hospital-associated methicillin resistant *S aureus* (MRSA) strains such as CC5, CC22, CC80, and CC30 were widely established in the 1990s and early 2000s, often carrying large multidrug-resistant staphylococcal cassette chromosome *mec* (SCCmec) elements. Since then, community-associated MRSA strains, including ST8 (CC8, USA300), ST59, and ST80, have emerged and expanded, frequently harbouring SCCmec IV or V. Livestock-associated MRSA ST398 has also become widespread, particularly in Europe and North America. While vancomycin resistance in *S aureus* remains rare, reduced susceptibility has been reported sporadically, especially in ST5 and ST8. Overall, the distribution of *S aureus* sequence types continues to shift regionally, with the spread of community-associated strains into hospitals. These patterns largely reflect data from high-income countries whereas strain distribution tends to be more heterogeneous in low-and middle-income countries. In *Enterococcus faecium*, genomic analyses have revealed a split population structure: clade A1 (clonal complex 17) comprises the vast majority of clinical, multidrug-resistant isolates, while clade B includes largely commensal, less drug-resistant strains circulating in the community. Globally, most clinical isolates now belong to clade A1 (CC17), which is characterised by near-universal ampicillin resistance, frequent vancomycin resistance, and additional resistance to aminoglycosides and fluoroquinolones. Since its emergence, CC17 *E faecium* has disseminated globally, with distinct subclones predominating in specific regions and time periods. In Europe and North America, ST17 and ST117 have become the predominant sequence types, commonly associated with vancomycin resistance and implicated in bloodstream infections. Although vancomycin resistance rates remain high in the USA and low in Canada, they have been relatively stable in recent years. Genomic epidemiology has shown that *Enterococcus faecalis* is more genetically diverse than *E faecium*, with a highly recombinogenic genome and no clear separation into distinct clades. Early vancomycin-resistant *E faecalis* reports in the 1990s were sporadic and genetically diverse, but by the 2000s and 2010s, expansions of hospital-associated strains such as ST6, ST87, and ST179 became more prominent, particularly in Europe and North America, frequently producing beta-lactamase and exhibiting resistance to vancomycin, and high-level gentamicin. For *Klebsiella pneumoniae* and *Escherichia coli,* studies have focused on the surveillance of extended-spectrum-beta-lactamase (ESBL) producing clones. The most common epidemic clones associated with multidrug resistance in *E coli* were ST131, ST1193, and ST69. The majority of ESBL-producing strains are attributed to CTX-M alleles with CTX-M-15 and CTX-M-14 being the predominant genotypes. Since 2000, the emergence of CTX-M-27, a single-nucleotide variant of CTX-M-14, has been increasingly reported. ESBL rates among hospital-associated *E coli* infections showed increasing trends in North America, Europe, and in many countries in Southeast and East Asia. ESBL rates varied considerably for *K pneumoniae* across different countries (15%–60%) with high rates primarily detected in Asia and in Southern and Eastern Europe. *K pneumoniae* bacteraemia was characterized by diverse strains with only a few associated with clonal outbreaks and hypervirulence, such as ST20, ST23, ST258, ST1. Only one study has comprehensively integrated clinical, genomic, and antibiotic prescribing data. Pöntinen and colleagues (2024) examined *E coli* bloodstream infections, showing that non-penicillin β-lactam use influenced the success of widespread multidrug-resistant, extended-spectrum beta-lactamase (ESBL)-producing *E coli* clones. However, no previous study has combined longitudinal clinical, genomic, and antibiotic use data across multiple major pathogens.

*Added value of this study:* To our knowledge, this study is the first to integrate longitudinal clinical, genomic, and antibiotic prescribing data for five key bacterial pathogens for an entire region over almost two decades. This combined dataset provides a unique opportunity to analyse the impact of changing antibiotic use on antimicrobial resistance trends across multiple organisms, as well as explore how pathogen-specific strains adapt to these changes over time.

*Implications of all the available evidence:* This study underscores the critical importance of integrated, pathogen-specific surveillance programs for guiding antimicrobial stewardship. The observed association between declining fluoroquinolone prescribing and reduced resistance among Gram-positive pathogens illustrates the potential impact of targeted prescribing interventions. At the same time, the heterogeneous responses across bacterial species to this change in practice highlight that stewardship strategies must be tailored, rather than one-size-fits-all. Continuous genomic surveillance is essential for anticipating resistance shifts and enabling timely detection and response to emerging threats, particularly from high-risk clones with the capacity for rapid dissemination. Finally, reducing antimicrobial resistance will require an integrated approach with hospital-based interventions along with coordinated efforts in the community to limit transmission and overall antibiotic exposure.

## Introduction

Antimicrobial resistance (AMR) is a major public health concern^1^ that emerges through the complex interplay of antimicrobial use, host and ecological factors, and bacterial population dynamics. However, how changes in antimicrobial prescribing translate into resistance trends through shifts in circulating bacterial strains is not yet fully understood. Bloodstream infections (BSIs) are a critical setting in which to examine these dynamics: they ranked as the seventh leading cause of death in 2019 and among the top three infectious syndromes linked to drug-resistant organisms.^2^

Understanding what drives AMR dynamics requires integrating epidemiological, genomic, and prescribing data – yet studies combining all three avenues of data remain rare. Most longitudinal cohort studies have used clinical and susceptibility data across multiple pathogens,^1,3–6^ while genomic studies typically focus on only one or two species.^6–9^ Although these studies revealed dynamic strain prevalence patterns over time, the forces driving these shifts remain unclear. Selective pressure from antimicrobial use has long been proposed as a key factor,^10^ and many phenotype-based studies have linked antibiotic use to resistance prevalence.^11–17^ For instance, fluoroquinolone, macrolide, and cephalosporin use were associated with increased resistance in pathogens such as *Staphylococcus aureus, Escherichia coli,* and *Streptococcus pneumoniae*.^11,17^ To date, only one study has comprehensively integrated clinical, genomic, and antibiotic prescribing data for *E coli* BSIs.^18^ Pöntinen and colleagues found that non-penicillin β-lactam use modulated the success of widely disseminated multidrug resistant extended-spectrum beta-lactamase (ESBL)-carrying *E coli* clones.^18^

Antimicrobial use not only shapes the population dynamics of the targeted pathogen but also those of other bacteria through bystander selection.^19^ However, given the lack of contemporaneous studies examining multiple pathogens in a single setting, it remains unclear to what extent shifts in antimicrobial selection drive similar or distinct strain dynamics across different pathogens. This knowledge gap hinders the development of predictive models that account for the complex interplay between antimicrobial use and resistance across bacterial pathogens, limiting our ability to design optimal clinical and public health strategies.

Our objective was to address this gap by integrating clinical, genomic, and antibiotic use data for five major causes of bacteraemia within the Calgary Health Zone (CHZ), Canada’s third-largest metropolitan area (population ∼1·7 million) with a fully integrated provincial healthcare system. We analysed trends in 23,512 routinely collected blood cultures for *S aureus*, *E faecalis*, *E faecium*, *E coli*, and *K pneumoniae* from Jan 1, 2006, to Oct 14, 2022. By examining AMR trends in bacterial genotypes and phenotypes across multiple antibiotic classes, we sought to identify how prescribing patterns and strain-level dynamics shape resistance trajectories, and how these relationships differ among pathogens. We hypothesized that selection pressures from population-level antimicrobial use would have distinct effects on population structure and resistance patterns for different pathogens, shaped by factors such as genetic backgrounds, mutational pathways, fitness costs, ecological niches, and baseline resistance profiles.

## Methods

### Study design and participants

This retrospective cohort study included antimicrobial prescribing, clinical, microbiological, and pathogen genomic data from all positive blood cultures in the CHZ from Jan 1, 2006, to Oct 14, 2022. The CHZ is a centralized health district with 14 hospitals providing inpatient and outpatient services to Calgary residents. Demographic information (age and biological sex) was obtained from birth certificates. The study compared pre–COVID-19 (2006–2019) and COVID-19 periods (2020–2022). Data for *Klebsiella pneumoniae* were limited to the pre–COVID-19 period; other species spanned the entire study period.

This study received ethical approval by the University of Calgary Institutional Review Board under protocol REB18-0233. As the study used de-identified, routinely collected clinical and microbiology data and involved no direct patient contact or intervention, the requirement for written informed consent was waived by both review boards in accordance with federal regulations.

### Procedures

Blood cultures were retrieved from Alberta Precision Laboratories (APL) and clinical records from Alberta Health Services (AHS). A bacteraemia episode included all positive blood isolates of the same species from a patient within 30 days, with the first isolate designated the *index isolate*. Episodes were classified as community-onset (collected within 48 hours of admission), and hospital-onset (between 48 hours and discharge), consistent with U.S. National Healthcare Safety Network’s definitions.^20^

Antimicrobial susceptibility testing (AST) was conducted using VITEK II (bioMérieux, Montreal, Canada) and interpreted according to Clinical and Laboratory Standards Institute (CLSI) breakpoints at the time of testing. Minimum inhibitory concentrations (MICs) were classified as susceptible (S), intermediate (I), or resistant (R); for analyses, intermediate and resistant were grouped. AMR phenotypes were defined based on AST results from pathogen-specific panels of first-and second-line antibiotics, referred to as *key* antibiotics (Appendix pp.7-8).

### Genomic analysis

Whole-genome sequencing (Table 1) was conducted using the Nextera XT DNA Library Preparation Kit (Illumina, San Diego, CA, USA) on the Illumina HiSeqX platform. Raw reads were processed with the seQuoia pipeline for adapter trimming, contamination screening, assembly, variant calling, and resistance/virulence gene detection.^21^ Isolates were considered successfully sequenced if they were cultured, sequenced, and met predefined quality control criteria. Downstream analyses included core genome alignment, recombinant region removal, and phylogenetic reconstruction. Genomic clusters, hereafter referred to as strains, were identified using StrainGST and assigned to known sequence types (STs) or clonal complexes (CCs).^22,23^ Subclades were defined using previously published classifications.^23^ Further details are provided in Appendix pp.9-10.

**Table 1.**
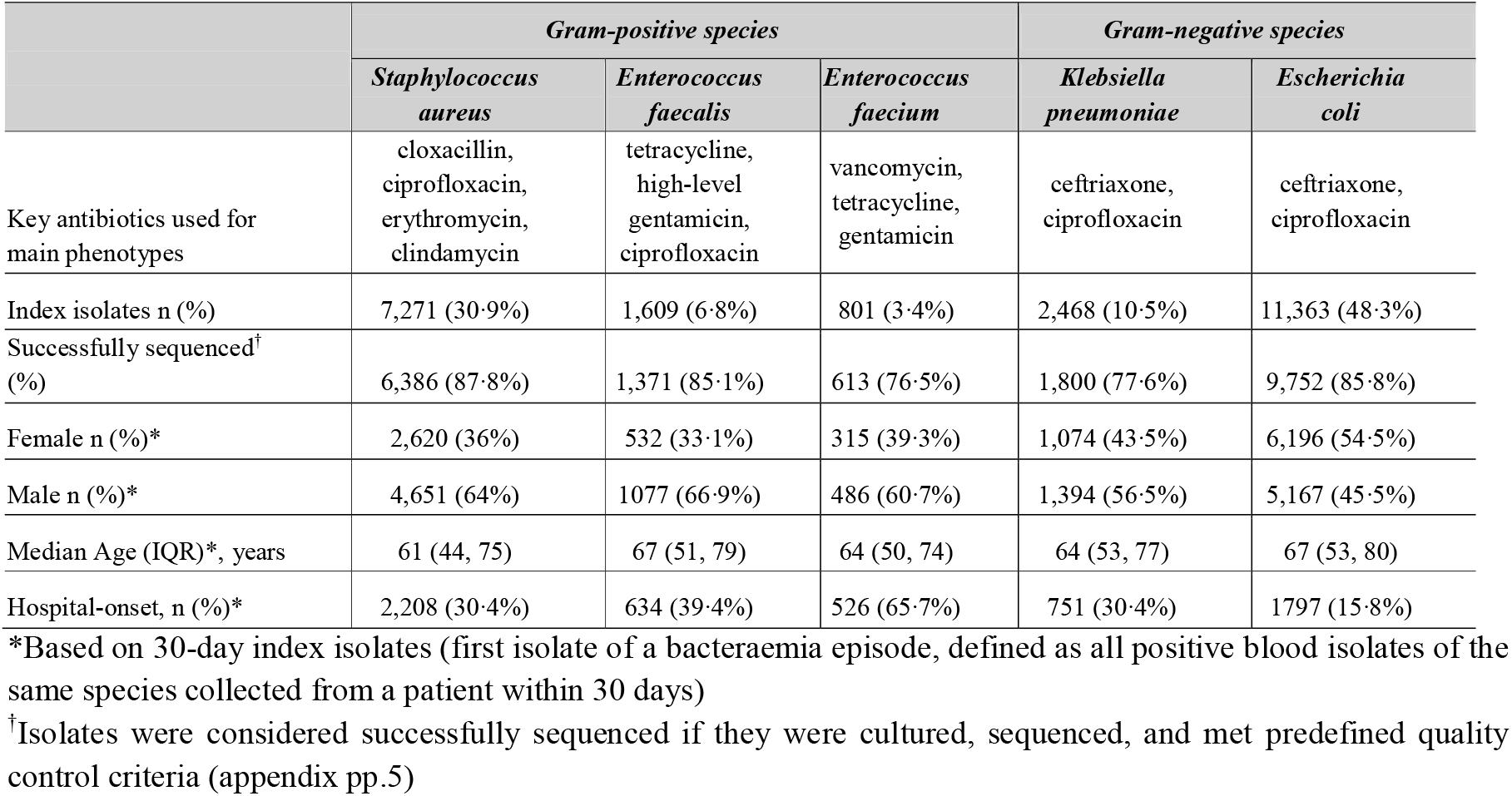
Descriptive statistics of blood cultures by organism, Jan 1, 2006 to Oct 14, 2022. Data are shown *for Staphylococcus aureus, Enterococcus faecalis, Enterococcus faecium, Klebsiella pneumoniae*, and *Escherichia coli*, including the key antibiotics used for AMR phenotypes, the number and percentage of index isolates, proportion successfully sequenced, patient sex distribution, median age with interquartile range (IQR), and proportion of hospital-onset infections. Percentages are calculated out of the total number of 30-day index isolates for each species. Bacteraemia episodes were classified as community-onset if the specimen was collected within 48 hours of admission, and hospital-onset if collected between 48 hours and discharge.

### Antimicrobial prescribing

Inpatient prescribing data were obtained from electronic medical administration records and pharmacy systems of AHS, covering all key antibiotics (Table 1), and converted to Defined Daily Doses (DDDs).^24^ Community prescribing data were sourced from the Pharmaceutical Information Network via AHS.^25^ An Antimicrobial Stewardship Program (ASP) has been active in CHZ since 2002, employing strategies including standardized order sets, cascade susceptibility reporting, and use of targeted prospective audit and feedback on selected wards to optimize prescribing (Appendix p.6).

### Infection prevention and control (IPC) measures

Infection prevention and control policies in the CHZ are described in Appendix pp.4, including changes in MRSA and VRE screening and contact precautions between 2006-2021, and the Alberta Health Services Hand Hygiene Program launched in 2011.

### Outcomes

Primary outcomes included bacteraemia incidence rates stratified by phenotypes and strains for pathogens of interest (Table 1). AMR was assessed using: (1) resistance proportion (percentage of index isolates classified as resistant or intermediate) and (2) phenotypic incidence, defined as the number of resistant isolates per 100,000 Calgary residents (community-onset) or per 100,000 patient days (hospital-onset). Secondary outcomes included community and inpatient prescribing rates, expressed as age-standardized dispensations per 100,000 residents and DDDs per 100 patient-days, respectively.

### Statistical analysis

Incidence rates of bloodstream infections were modelled per year for each organism using negative-binomial or Poisson regression, with Calgary resident counts or patient-days as offsets for community- and hospital-onset, respectively. Segmented regression identified trend change points (Appendix p.7).^26^ Time trends were quantified using average annual percentage change (AAPC) defined as the average rate of change in incidence per year. Shannon diversity indices were calculated based on genomic clusters. Analyses were performed in R version 4.4.0.

## Results

A total of 23,512 bloodstream index isolates were recovered from 20,089 patients between Jan 1, 2006 – Oct 14, 2022. The most isolated species was *E coli* (N=11,363, 48·3%), followed by *S aureus* (N=7,271, 30·9%), *K pneumoniae* (2,468, 10·5%), *E faecalis* (N=1,609, 6·8%), and *E faecium* (N=801, 3·4%) (Table 1). Sequencing success rates ranged from 76·5% for *E faecium* to 87·8% for *S aureus* (Table 1). For most species-antibiotic combinations, susceptible isolates did not encode known resistance determinants, and nearly all resistant isolates encoded at least one resistance-associated gene or allele, except for high-level gentamicin susceptibility/resistance among *Enterococcus* species (Appendix pp.11-18) where limitations of short-read sequencing likely impacted detection of resistance genes.

### Increase in community-onset and decline in hospital-onset bacteraemia

Overall, cumulative incidence of community-onset bacteraemia (COB) across all five species increased between 2006-2018 (AAPC [95%CI]=4·5%, [4·0%, 5·0%], p<0·0001, Figure 1A), followed by a decline between 2019-2022 (AAPC=−10·3%, [−12·5%, −8·0%], p<0·0001) though individual species differed. While COB caused by *S aureus* and *E coli* followed this pattern, *E faecalis* and *K pneumoniae* increased steadily throughout the study period (Appendix p.27). *E faecium* COB increased markedly between 2006–2011 (AAPC=17·7% [1·2, 36·8], *p*=0·0342) before declining after 2012, though the confidence interval included the null (AAPC=–1·0% [–5·1, 3·3], p=0·3070).

**Figure 1.**
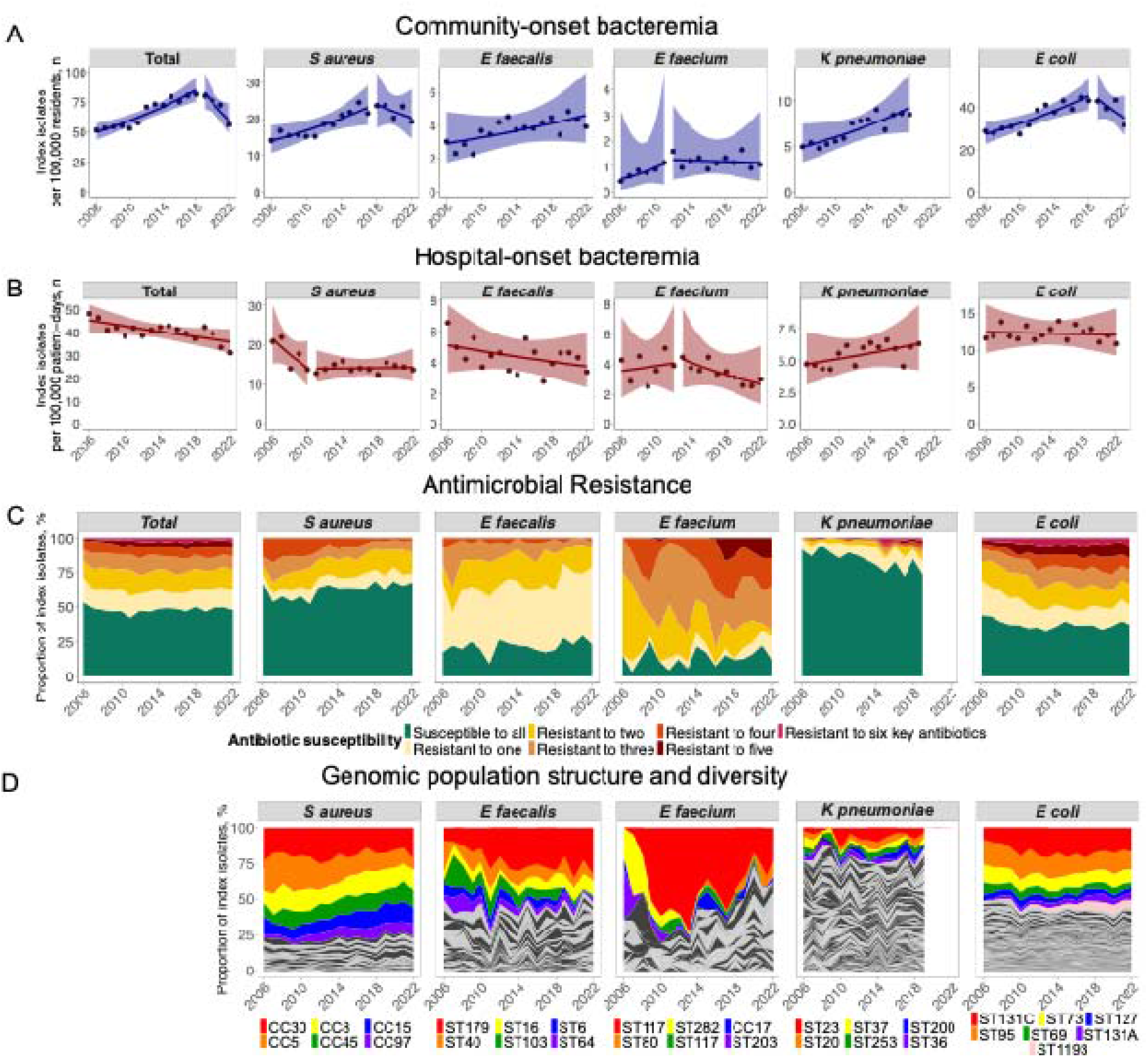
Trends in bacteraemia incidence, antimicrobial resistance, and genomic population structure from Jan 1, 2006 - Oct 14, 2022, for five target bacterial pathogens. (A) The number of 30-day index isolates per 100,000 Calgary residents per pathogen classified as community-onset are represented as points. Time trends were estimated using segmented Poisson regression and are represented as lines and corresponding 95% confidence intervals. (B) Number of 30-day index isolates per 1,000 patient days per organism that were classified as hospital-onset. Isolates were classified as community-onset if the specimen was obtained within 48 hours of admission. Otherwise, it was classified as hospital-onset. (C) Proportion of 30-day index isolates susceptible or resistant to 1, 2, 3, …, 6 antibiotics per organisms. Key antibiotics were chosen based on clinical relevance. *S aureus*: cloxacillin, ciprofloxacin, erythromycin, clindamycin; *E faecalis*: tetracycline, high-level gentamicin, ciprofloxacin; *E faecium*: vancomycin, tetracycline, gentamicin; *K pneumoniae*: ceftriaxone, ciprofloxacin; *E coli*: ceftriaxone, ciprofloxacin. (D) Proportion of 30-day index isolates classified to the 20 most common StrainGST reference clusters per species. StrainGST reference clusters were annotated with the corresponding clonal complex or multilocus sequence type. For *K pneumoniae*, isolates were only collected for Jan 1, 2006 - Dec 31, 2019.

In contrast, hospital-onset bacteraemia (HOB) declined overall (AAPC=−1·5% [−2·1, −0·8], *p*<0·0001; Figure 1B), largely due to Gram-positive organism reductions. The steepest decline was in *S aureus* between 2006-2010 (AAPC=−9·8% [−14·7, −4·6], *p*=0·0003). *K pneumoniae* HOB increased modestly (AAPC=2·0% [0·2, 3·8], *p*=0·0321), while *E coli* HOB remained stable (AAPC=−0·3% [−1·2, 0·7]).

### Decline in multidrug resistance in Gram-positive species

The proportion of multidrug-resistant isolates (resistant to ≥2 key antibiotic classes) generally declined among Gram-positive bacteria from 2006 to 2022 (Figure 1C). In *S aureus*, the proportion resistant to all four key antibiotics (Cloxacillin–Ciprofloxacin–Erythromycin–Clindamycin; R-R-R-R phenotype) fell from 3·4% in 2006 to 0·4% in 2022, with a steeper decline for HOB than for COB (Figure 2A, Appendix p.26). The proportion of methicillin-resistant *S aureus* (MRSA) bacteraemia also declined from 20·1% to 14·1% over the same period (Figure 5B). In *E faecium*, vancomycin resistance proportion increased from 0% in 2006 to a peak of 50·9% in 2013, then declined to 29·2% in 2022 (Figure 5B). In *E faecalis*, high-level gentamicin resistance proportion dropped from 21·0% in 2006 to 10·8% in 2019, before rising again to 14·6% in 2022 (Figure 5B).

**Figure 2.**
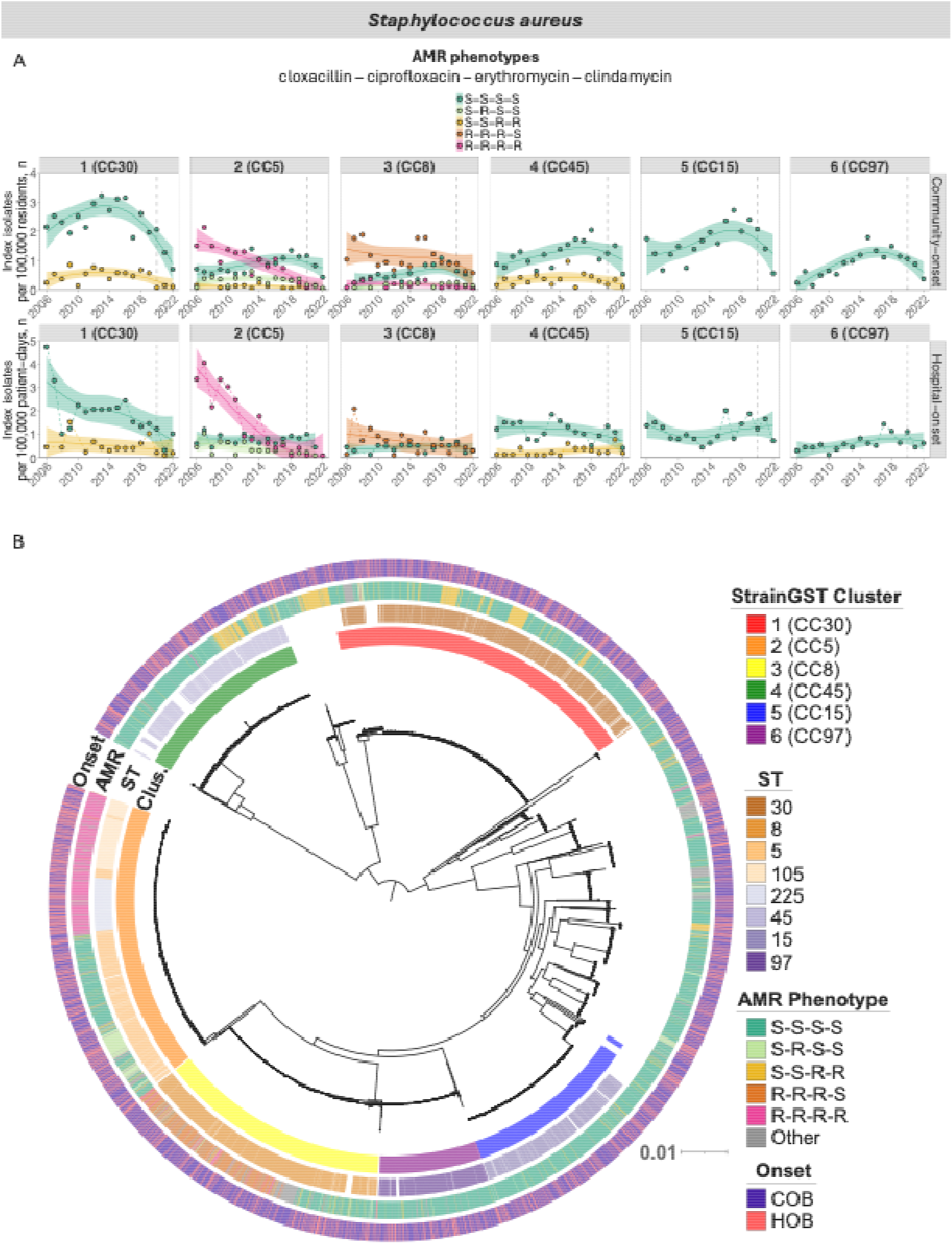
Incidence and genomic population structure for *Staphylococcus aureus* bacteraemia. (A) Incidence of *S aureus* bacteraemia stratified by acquisition type, by five most common AMR phenotypes (colours), and by six most common sequence clusters (and assigned clonal complexes; facets). Cloxacillin, clindamycin, erythromycin, and ciprofloxacin were used for phenotypes. For each sequence cluster, the incidence of phenotypes is shown for which there were at least 20 index isolates. Points represent observed 30-day index isolates per 100,000 Calgary residents/patient days. Lines and shaded areas represent rates and 95% confidence intervals estimated by loess regression. (B) Midpoint-rooted core genome phylogenetic tree of index isolates from unique *S aureus* bacteraemia cases. Inner rings one and two represent sequence clusters and STs, respectively, with legends presented in order from most to least common. The predominant CC associated with each cluster is shown in parentheses. The second outermost ring (ring three) illustrates the phylogenetic relationships of isolates together with their common antimicrobial resistance phenotypes, while the outermost ring (ring four) denotes whether isolates were classified as community-onset or hospital-onset. AMR = antimicrobial resistance; ST = sequence type; CC = clonal complex.

AMR phenotypes were largely present in a few select strains for each Gram-positive species: 80% of sequenced cases were represented by five strains in *S aureus* and *E faecium* and by ten strains in *E faecalis* (Figure 1D, 2B, 3B/D). Among Gram-positives, *E faecium* exhibited the lowest median Shannon strain diversity index, indicating low strain diversity, whereas *E faecalis* exhibited the highest, reflecting greater strain heterogeneity (Appendix p.35).

### Decline of dominant multidrug-resistant MRSA and expansion of susceptible strains

Multidrug resistance in *S aureus* was concentrated in CC5 and CC8. Most isolates with the R-R-R-R phenotype (MRSA co-resistant to three other key antibiotics) belonged to monophyletic subclades of CC5-II B (ST225 and ST105/ST5; Appendix p.19), which declined steadily from 2006–2022, with steeper decreases in HOB than COB (Appendix p.27). Within CC8, the incidence of isolates with the R-R-R-S phenotype also declined, particularly in HOB (AAPC=−6·4% [−11·7, −0·7], *p*=0·0276). These declines occurred during a period of changing MRSA admission screening practices (Appendix p.4): initially restricted to patients with a known history of MRSA (2006– 2011), subsequently broadened to all medical and surgical admissions (2011–2015), and ultimately replaced by a risk-based approach.

Methicillin-susceptible *S aureus* (MSSA) isolates with the fully susceptible phenotype (S-S-S-S) were distributed across the phylogeny (Figure 2B) and increased across multiple clonal complexes (Figure 2A), especially in CC97 (2006–2015), CC8 (2006–2019), and CC5 (2006–2019), but not in CC30 (Appendix pp.27). These shifts were not associated with biological sex, age group, or substance-related disorders (Appendix pp.30-31). MSSA COB incidence subsequently declined across all strains, though with variable timing: declines in CC5, CC8, and CC15 coincided with the COVID-19 period whereas incidence of other strains began declining before 2020 (Appendix pp.30).

### Mixed clonal and diverse population structure of *E faecalis*

In *E faecalis*, the most common phenotype was tetracycline-resistant but susceptible to high-level gentamicin and ciprofloxacin (R-S-S, N = 748/1609, 46·4% of cases) and clustered mainly within ST179, ST40, ST16, and ST64 (Figure 3). Within ST179, the R-R-S phenotype showed an upward trend in HOB from 2006–2014, followed by a sharp decline from 2015–2022 (AAPC=–28·1% [–44·1, –7·4], *p*=0·0025). In contrast, the common R-S-S phenotype increased in ST179 (AAPC=8·2% [3·3, 13·2], *p*=0·0008) but declined in ST40 (AAPC=–4·4% [–9·2, 0·7]). Age distributions differed between these two STs (Appendix p.34), with ST179 underrepresented in children aged 0–9 years (12·2% vs 19·3%, *p*=0·0440) and in adults aged 60–69 years (16·3% vs 24·6%, *p*=0·0352), while no differences were seen by biological sex.

**Figure 3.**
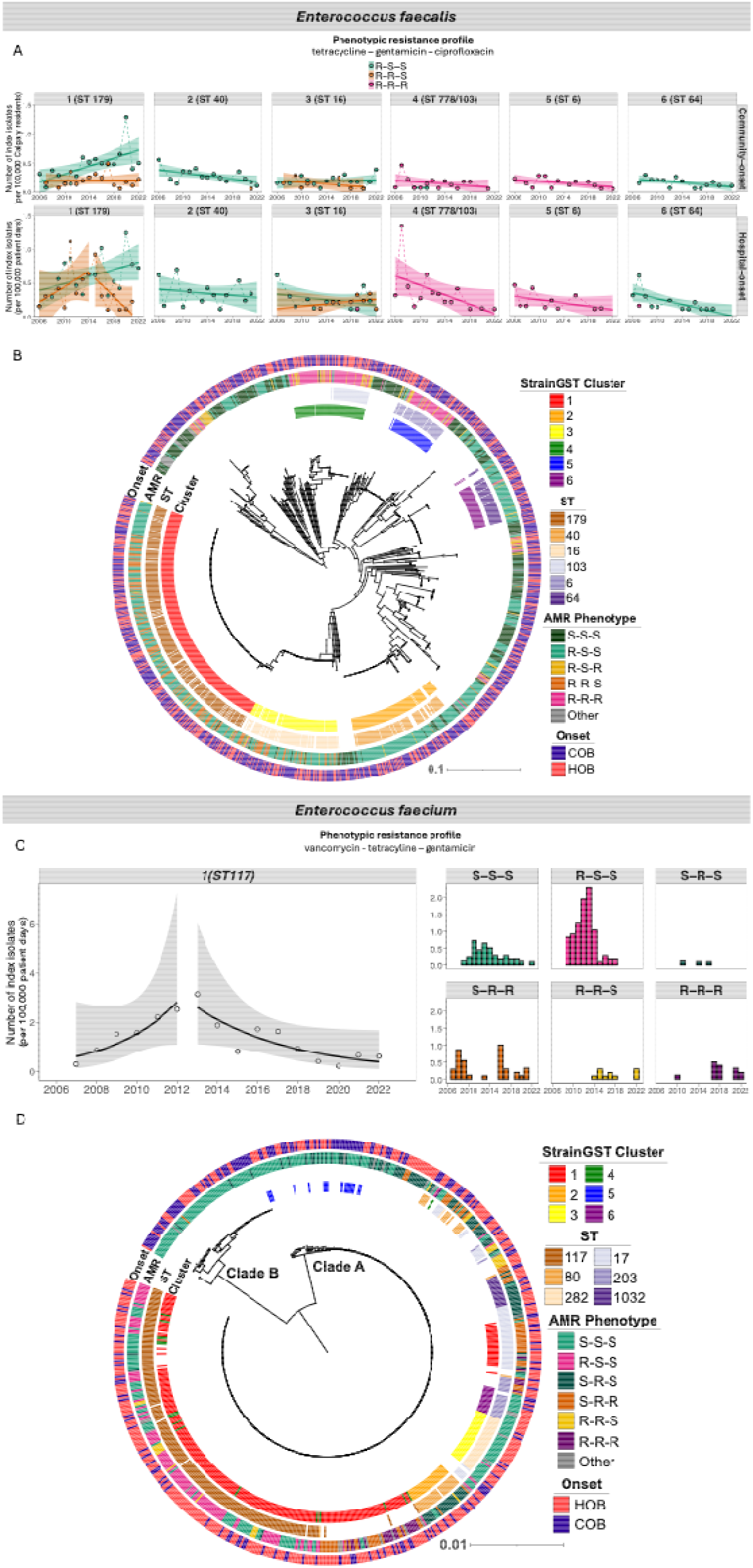
Incidence and genomic population structure for *E faecalis* and *E faecium* bacteraemia. (A) *E faecalis* bacteraemia incidence, stratified by acquisition type (community-vs hospital-onset), three most common resistance phenotypes (based on tetracycline, high-level gentamicin, and ciprofloxacin), and the six most common reference clusters with assigned clonal complexes (colored areas). Only clusters with ≥5 index isolates are shown for each phenotype; ST778 and 103 were merged. Points show observed rates; lines and shaded areas represent estimates and 95% confidence intervals from *loess* regression. Isolates were classified as community-onset (COB) if obtained within 48 hours of admission, and hospital-onset (HOB) if obtained between 48 hours and discharge. (B) Midpoint-rooted phylogenetic tree of *E. faecalis* index isolates from unique bacteraemia cases. Inner rings one and two show sequence clusters and sequence types (ST), respectively, with the legends in order from most to least common. The second outermost ring (ring three) illustrates the phylogenetic relationships with common AMR phenotypes. The outermost ring (ring four) indicates whether isolates were classified as community-onset or hospital-onset. (C) Incidence rate of hospital-onset *E. faecium* ST117 bacteraemia per 100,000 patient days, shown overall (left) and stratified by the six most common resistance phenotypes (right), based on vancomycin, tetracycline, and high-level gentamicin susceptibility. Points and bars show observed rates; lines and shaded areas represent estimates and 95% confidence intervals from segmented regression. (D) Midpoint-rooted phylogenetic tree of *E. faecium* index isolates from unique bacteraemia cases. Rings are annotated in the same manner described in (B).

Highly resistant *E faecalis* isolates (resistant to all three key antibiotics) were rare and clustered mainly within ST6 (N=47), ST103 (N=36), and ST778 (N=22), with declining incidence of ST103 in HOB (Appendix p.28).

### Trends in *E faecium* were mainly driven by ST117

For *E faecium*, incidence trends were primarily driven by ST117, a globally recognized hospital-adapted lineage within clade A1 of CC17 (Figure 1D) and dominated by a hospital-onset subclade resistant to vancomycin (Figure 3C-D). The incidence of this subclade in HOB increased between 2006-2013 (AAPC=36·7%, [3·3%, 80·9%], p=0·0286), followed by a sharp decline from 2014 to 2022 (AAPC=−26·9%, [−37·0%, −15·3%], p=0·0001). These trends occurred alongside changes in IPC (Appendix p.4-5): the decline in incidence followed the expansion of VRE rectal swab screening to all medical and surgical admissions between 2011 and 2015, after which screening was restricted to three select high-risk units and subsequently phased out. Despite the discontinuation of routine rectal swab screening, the incidence of VRE *E faecium* bacteraemia continued to decline after 2015, coinciding with sustained improvements in hand hygiene compliance.

### Increase of ceftriaxone and ciprofloxacin resistance in *Enterobacterales*

The population structure of both *Enterobacterales* species was highly diverse (Figure 1D, 4D), with *K pneumoniae* exhibiting the highest Shannon strain diversity index (80% of cases represented by 554 strains), and *E coli* the second highest (80% of cases represented by 27 strains). COB incidence increased for both species, driven primarily by phylogenetically diverse, susceptible strains (Appendix p.29). Fully susceptible *E coli* (susceptible to cefazolin, ceftriaxone, ciprofloxacin, gentamicin, and trimethoprim/sulfamethoxazole) increased from 2006-2018 (AAPC=2·8% [1·7%, 4·0%], p<0·0001) before declining from 2019-2022 (AAPC=−5·5% [−11·0%, 0·0%], p=0·0062). *K pneumoniae* showed a similar pattern between 2006-2019 (AAPC=3·7% [2·3%, 5·2%], p<0·0001).

In *E coli,* ceftriaxone and ciprofloxacin resistance were strongly linked: 42·6% (1548/3637) of ciprofloxacin-resistant isolates were also ceftriaxone resistant, and 78·3% (1548/1978) of ceftriaxone-resistant isolates were ciprofloxacin resistant. Ciprofloxacin-ceftriaxone-resistant *E coli* rose mainly in COB, driven by ST131 (AAPC=36·8% [21·7%, 53·8%], p<0·0001) between 2006–2011, and by ST1193, which emerged in 2008 and increased steadily thereafter (AAPC=17·7% [9·0%, 30·0%], p<0·0001) (Figure 4A-B).

**Figure 4.**
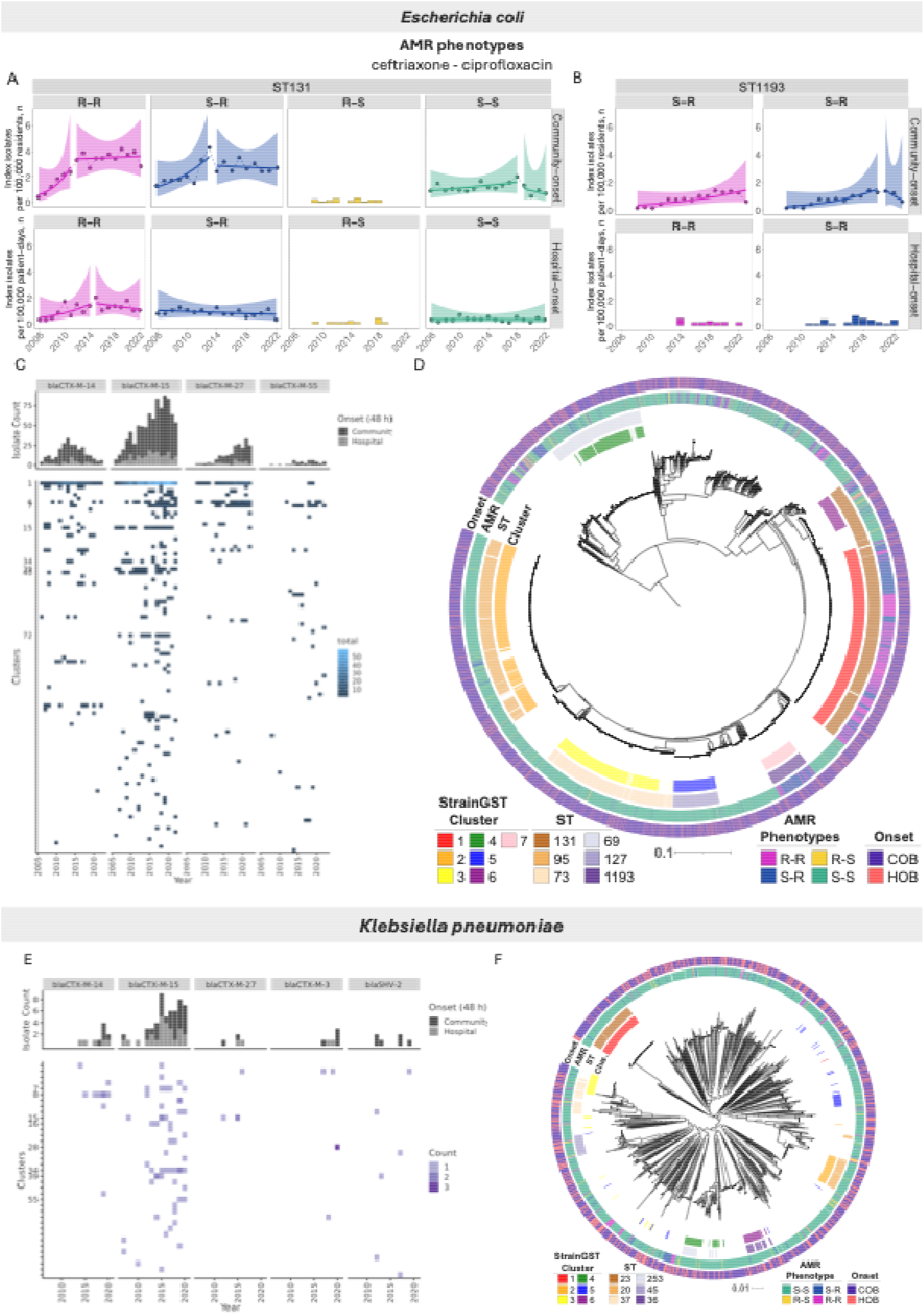
Bacteraemia Incidence and Genomic Context of Ceftriaxone and Ciprofloxacin Resistance in Major *E coli* and *K pneumoniae* strains. (A) Incidence rate of *E. coli* ST131 bacteraemia stratified by acquisition type (community-vs hospital-onset) and ceftriaxone–ciprofloxacin resistance profile. Points represent observed 30-day index isolates per 100,000 Calgary residents/patient days. Lines and shaded areas represent rates and 95% confidence intervals estimated by loess regression. Bar plots show observed counts when regression was not performed due to limited sample size. Colors denote resistance profiles. (B) Incidence rate of *E. coli* ST1193 bacteraemia, similarly stratified by acquisition type and resistance profile, with plotting conventions as in (A). (C) Number of 30-day index *E. coli* isolates with CTX-M β-lactamases (top) and their distribution across sequence clusters. Clusters are labeled if more than ten isolates encoding an ESBL were identified. (D) Midpoint-rooted phylogenetic tree of *E coli* index isolates from bacteraemia cases. Rings one and two show sequence clusters and sequence types (ST), respectively, with the legends in order from most to least common. Ring three illustrates the phylogenetic relationships with common AMR phenotypes. Ring four indicates whether isolates were classified as community-onset or hospital-onset. (E) Number of 30-day index *K pneumoniae* isolates with CTX-M β-lactamases (top) and corresponding sequence cluster distribution. Clusters are labeled if more than three isolates encoding an ESBL were identified. (F) Midpoint-rooted phylogenetic tree of *K pneumoniae* index isolates. Rings are annotated in the same manner described in (D). Isolates were classified as community-onset (COB) if obtained within 48 hours of admission, and hospital-onset (HOB) if obtained between 48 hours and discharge.

Ceftriaxone resistance was primarily due to CTX-M-type ESBLs. Within ST131 and ST1193, the CTX-M-27 allele emerged (Figure 4C). In ST131, CTX-M-27 emerged in 2008 and increased from 0·15 to 1·1 per 100,000 residents by 2022 (AAPC=23·8%, [17·4%, 30·5%], *p*<0·0001), while CTX-M-15 increased from 0·2 in 2006 to 2·2 in 2022 (AAPC=9·8%, [6·1%, 13·7%], *p*<0·0001). CTX-M-14 expanded from 0·2 in 2006 to 1·7 in 2013 (AAPC=53·6%, [31·9, 79·0], *p*<0·0001) before declining thereafter (AAPC=−15·9%, [−21·3%, −10·2%], *p*<0·0001). These CTX-M variants were associated with distinct ST131 subclades (Appendix p.22). In ST1193, CTX-M-27 appeared in 2013 and increased to 3 isolates in 2022 (12·0% of 25 isolates), while CTX-M-15, first detected in 2015, peaked at 7 isolates (28%) in 2017 and 2019, before declining to 3 in 2022. In both ST131 and ST1193, CTX-M-15- and CTX-M-27-positive isolates had higher ceftazidime MICs than CTX-M-14 (Appendix p.40). Among ST131 isolates, CTX-M-14 MICs reached 16 µg/mL, whereas 11·1% of CTX-M-15-positive and 2·8% of CTX-M-27-positive isolates had MICs ≥32 µg/mL.

Unlike *E coli*, the dynamics of ceftriaxone-resistant *K pneumoniae* in Calgary were not driven by any one strain. Similar to *E coli*, ESBL-producing *K pneumoniae* increased between 2006–2019 (Figure 4E), with ceftriaxone– ciprofloxacin resistance rising overall (AAPC=19·2% [9·9, 29·3], p<0·0001). *K pneumoniae* isolates encoding CTX-M-15 in Calgary were more closely related to global CTX-M-15 encoding strains compared to susceptible *K pneumoniae* strains from Calgary. We did not detect ST258, a globally prevalent and clinically significant lineage. The hypervirulent ST23 (4·9% of isolates), associated with severe invasive infections, was mostly community-onset, susceptible to key antibiotics, and showed only a modest, non-significant increase (AAPC=2·3% [−4·4, 9·4]).

### Prescribing patterns and associated resistance

Declines in ciprofloxacin resistance among Gram-positive bacteria coincided with reduced fluoroquinolone prescribing (Figure 5). Inpatient prescribing fell from 11·4 to 3·6 DDD per 100 patient-days between 2006–2022 (AAPC=−5·1%, [−6·5%, −3·7%], p<0·0001), and community prescribing in CHZ dropped from 61·6 in 2010 to 23·0 in 2022 (AAPC=−6·8%, [−8·1%, −5·4%], p<0·0001). Declines in macrolide and lincosamide prescribing paralleled reductions in erythromycin and clindamycin resistance in *S aureus* (Figure 5). These trends occurred alongside key infection prevention initiatives in the CHZ: an Antimicrobial Stewardship (AS) Program with an active AS Committee promoting evidence-based prescribing from 2002 onwards, and an Infection Prevention and Control Hand Hygiene Program established in 2011, which introduced standardised training, infrastructure improvements, and compliance monitoring against a provincial target of 90% (Appendix p.6).

**Figure 5.**
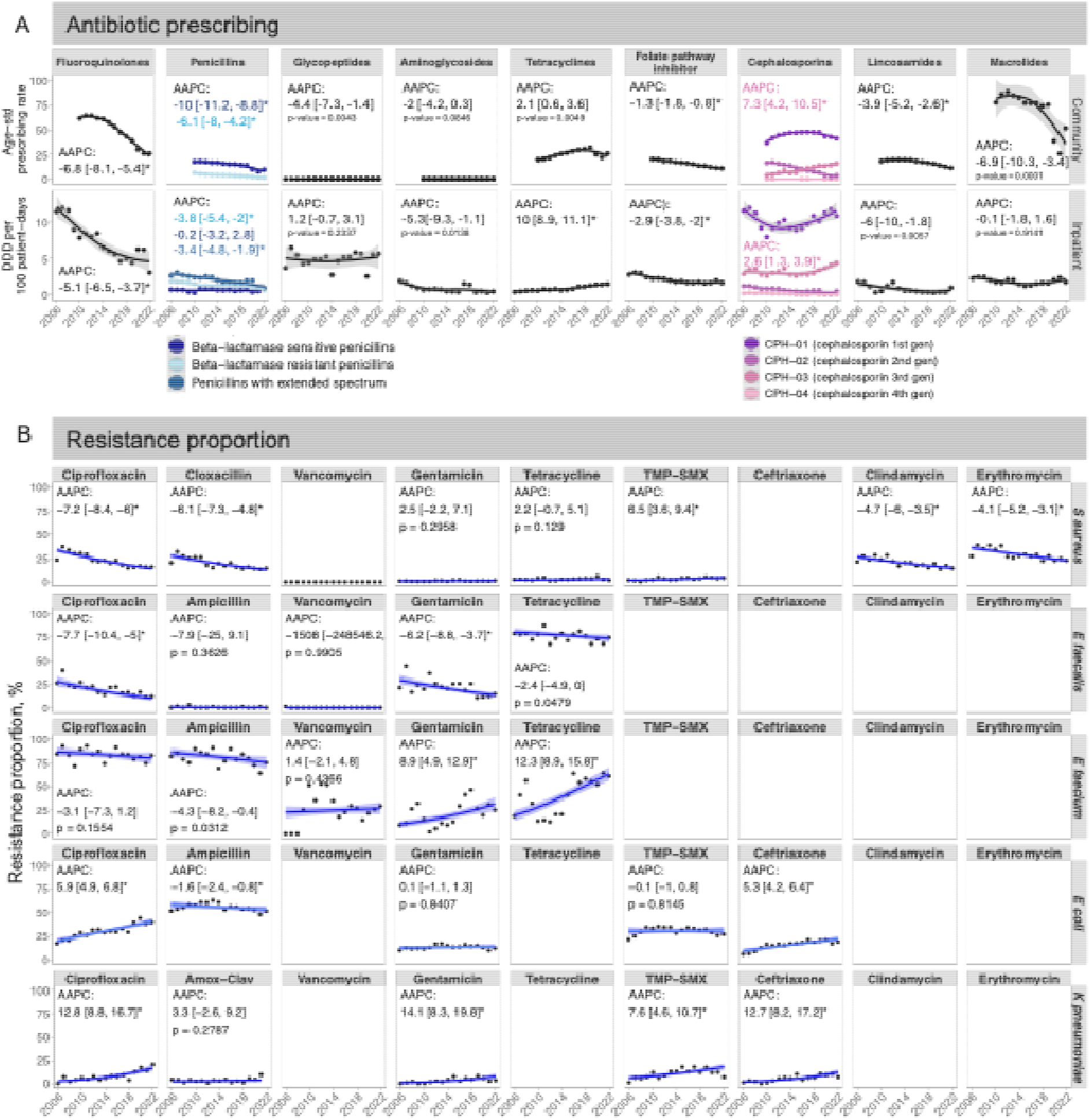
Temporal trends in antibiotic prescribing and resistance proportion across five target species and corresponding key antibiotic classes from 2006-2022. (A) Community and inpatient antibiotic prescribing of key antibiotic classes. Community prescribing is reported as age-standardized prescribing rate in the Calgary Health Zone from 2010 to 2022; inpatient prescribing as defined daily doses (DDD) per 100 patient-days from 2006 to 2022 in hospitals of the Calgary Health Zone. Data is shown as points. Lines and shaded bands are smoothing curves using the locally estimated scatterplot smoothing method and corresponding 95% confidence intervals. Average annual percentage change (AAPC) for time trend analyses are given next to the curves. For cephalosporins, time trends are only reported for third-generation cephalosporins. (B) Proportion of 30-day index isolates that were resistant to key antibiotic classes for five target species from 2006 to 2022. Data is shown as points. Lines and shaded bands are time trend estimates and 95% confidence intervals from Binomial regression. Abbreviations for antimicrobials:, TMP-SMX = trimethoprim-sulfamethoxazole, Amox-Clav = amoxicillin-clavulanate.

In contrast, ciprofloxacin resistance increased in *E coli* and *K pneumoniae*. Despite overall declines, outpatient fluoroquinolone prescribing increased among patients with *E coli* bacteraemia between 2006–2017 (Appendix p.37-38), and fluoroquinolone prescribing 30-days prior to the bacteraemia was associated with resistance in *E coli* (Appendix pp.39). Ceftriaxone resistance also increased, paralleling higher cephalosporin prescribing (Figure 5A). Third-generation cephalosporin inpatient use increased from 2·9 to 5·1 DDD per 100 patient days between 2006– 2022 (AAPC=2·6%, [1·3%, 3·9%], p=0·0001), while community prescribing grew from 3·8 in 2010 to 15·8 in 2022 (AAPC=7·3%, [4·2%, 10·5%], p<0·0001).

Carbapenem inpatient prescribing increased over the study period (Appendix p.41), though carbapenem resistance remained rare, peaking at three isolates in *E coli* and two isolates per year in *K pneumoniae* (Appendix p.40).

## Discussion

From 2006 to 2022, the incidence of bacteraemia, AMR, and strain dynamics varied substantially among five major pathogens in the CHZ, reflecting pathogen-specific interactions between antibiotic exposure, clonal expansion, and resistance evolution. Notably, we observed few pandemic-associated shifts in resistance trends in our setting, suggesting that observed patterns were shaped primarily by longer-term prescribing and strain dynamics.

Despite prevailing expectations that increasing bacteraemia incidence would be accompanied by proportional increases in AMR, increases in community-onset bacteraemia were largely driven by expansion of pre-existing strains susceptible to many first- and second-line antibiotics in our data. This pattern mirrors European reports of increasing MSSA and declining MRSA prevalence,^27–30^ but extends beyond methicillin to three additional important antibiotics in *S aureus*. In our study, these shifts were independent of patient age and sex, though other factors driving the increase in MSSA, such as changing population demographics in the region^31^ and importation of strains from other countries, warrant further investigation. Our findings highlight that reducing overall infection incidence, not just resistant infections, is critical to limiting antibiotic pressure and ultimately the emergence of resistance.^32^

The decline in multidrug-resistant MRSA likely reflects a combination of hospital-based infection control measures, antimicrobial stewardship, and intrinsic evolutionary dynamics of *S aureus*. Sustained reductions in fluoroquinolone prescribing in both inpatient and outpatient settings may have reduced selective pressure favouring certain MRSA lineages.^33^ Hospital-acquired MRSA lineages carrying large type-II SCCmec elements have been shown to offset the metabolic cost of resistance by reducing toxin expression, making them reliant on vector-mediated transmission through healthcare workers and vulnerable hosts rather than virulence-driven community transmission.^34^ While the MRSA decline began in 2006, the province-wide Hand Hygiene Program established in 2011 likely reinforced these trends by disrupting hospital transmission.^35,36^ Consistent with this interpretation, VRE bacteraemia continued to decline after 2015 despite discontinuation of routine rectal swab screening, indicating that sustained hand hygiene compliance, rather than active surveillance alone, drove the reduction.

The systematic integration of genomic data allowed us to disentangle clonal expansion from polyclonal spread – a distinction critical for intervention design. When incidence was driven by clonal expansion, such as *S aureus* CC5 or *E coli* ST131, targeted strategies (e.g., strain-specific diagnostics, screening, or vaccination strategies) may be most effective.^37^ In contrast, polyclonal expansion of MSSA suggests the need for broader measures, including reducing prescribing across multiple antibiotic classes, general infection control strategies such as environmental cleaning or universal decolonization, and community-level interventions, such as improved outpatient stewardship and strengthened infection prevention in long-term care centres.

Genomic surveillance also revealed ongoing evolution within a dominant clone. In *E coli* ST131, we observed the rise of CTX-M-27 alongside a decline in CTX-M-14. This shift is concerning, as CTX-M-27 genes are frequently carried on plasmids harbouring resistance determinants against other antibiotic classes, including fluoroquinolones and carbapenems.^38–40^ Consistent with previous findings, isolates from our study carrying CTX-M-27 showed higher ceftazidime MICs than those with CTX-M-14, suggesting ceftazidime use may be selecting for this allele.^41^ This exemplifies how resistance evolution continues even within successful clones, necessitating continued genomic monitoring.

Our results highlight the complexity of use-resistance relationships across different settings, pathogens, and host populations. Despite overall reductions in fluoroquinolone prescribing in both community and inpatient settings, we observed contrasting trends between bacterial species. While fluoroquinolone resistance declined in Gram-positive species, it increased in *E coli* and *K pneumoniae*. This pattern coincided with increased outpatient fluoroquinolone prescribing specifically among patients with *E coli* bacteraemia, suggesting heightened selection pressure in this group.^42^ Furthermore, the increase in third-generation cephalosporin prescribing, likely substituted for fluoroquinolones, may have inadvertently co-selected for fluoroquinolone resistance through linked resistance mechanisms, thereby undermining the intended benefits of stewardship efforts.

Our study has several limitations. First, the observed dynamics may have been influenced by other factors that we were unable to assess, including importation of strains from outside the CHZ, temporal variations in diagnostic and reporting practices, shifts in patient populations (beyond those we studied), and changes in IPC (beyond those reported here). Second, we focused exclusively on data from bacteraemia cases. The inclusion of information on bacterial colonization or clinical infections would enhance our analysis and provide a more comprehensive understanding of AMR dynamics. It is well-established that the strains responsible for invasive disease may differ from those associated with colonization.^43^ Third, we classified hospital-onset bacteraemia using a 48-hour cutoff, which may misclassify some cases, such as infections acquired in healthcare settings but detected early, or community-acquired infections that present after 48 hours. Fourth, community antibiotic prescribing data covered only part of the study period, limiting our ability to fully capture exposure patterns, although it still represents a substantial proportion of the reporting years. Lastly, resistance profiling relied on AMRFinderPlus; while most resistance phenotypes were well explained by known markers, unexplained cases remained, such as high-level gentamicin resistance in *Enterococcus* species, where the major genetic determinant of resistance was not always present in assembled genomes despite evidence for its presence in raw sequencing reads.

In conclusion, integrating genomic surveillance with clinical and prescribing data provided critical insights into the multifactorial drivers of AMR. Effective mitigation will likely require coordinated, multifaceted interventions, including genomic surveillance, community-focused strategies to reduce overall infection incidence, and stewardship programmes that explicitly address co-selection pressures. As resistance mechanisms continue to evolve and diversify, so too must our interventions to combat them.

## Data Availability

Individual-level patient data cannot be provided due to Alberta Health Services privacy practices. Summary and artificial data that represents the original data will be provided along with a data dictionary that defines each field in the dataset and supporting documentation (statistical and analytic code) are published on Github (https://github.com/tm-pham/LSARP_amr_genomics.git). Genomics data are available from www.resistanceDB.org

https://www.resistancedb.org/

## Contributors

Conceptualisation: IAL, YHG, AME, TMP, TM, JS

Data curation: SW, AME, BW, TMP, JS, TM, MEV-T, BD

Investigation: TMP, JS, TM, BW, DBG, KH, AW, AM, AUL, TR, GR, MM, JP, LPA, MEV-T

Methodology: TMP, TM, JS, YHG, AME, JC, IAL

Formal analysis: TMP, TM, JS

Funding acquisition: IAL

Project administration: IAL, RN, NS, TDF

Software: TMP, TM, JS, RS

Supervision: IAL, YHG, AME

Validation: TM, JS

Visualisation: TMP, TM, JS

Writing – original draft: TMP

Writing – review & editing: TMP, TM, JS, AME, YHG, JC, IAL, RS

All authors revised the content of the manuscript critically and approved the final version. All authors had full access to all the data in the study and had final responsibility for the decision to submit for publication.

## Consortium Authorship

The Precision Infection Management and Epidemiology (PRIME) Consortium consists of members who directly contributed to this work: Thi Mui Pham PhD^1,2,3,*^, Joshua T. Smith PhD^*,4^, Tatum D. Mortimer PhD^*,5^, Daniel B. Gregson MD^6,7^, Bruce Walker^4^, John Conly MD DSc^7,8,,9^, Karen Hope MSc^9^, Bruce Dalton Bsc^10^, Andriy Plakhotnyk Bsc^6^, Sören Wacker PhD^6^, Rauf Salamzade PhD^4^, Anika Westlund Bsc^6^, Rory Gilliland MSc^6^, Alikhan Mansuri Msc^6^, Annegret Ulke-Lemée PhD^6^, Mario E. Valdés-Tresanco PhD^6^, Thomas Rydzak PhD^6^, Gopal Ramamourthy PhD, Nithya Swaminathan MPH ^4^, Rachel Newmiller BS^4^, Troy D. Feener MSc^6^, Maryam Mapar PhD^6^, Jenna Poelzer^6^, Ryan A. Groves MSc^6^, Luis Ponce Alvarez Msc^6^, M. Ethan MacDonald PhD PEng^11^, Hallgrimur Benediktsson^12,13^, Fiona Clement PhD^14^, Sergei Y. Noskov PhD^15^, Yonatan H. Grad MD Prof^2,3,†^, Ashlee M. Earl PhD^4,†^, Ian Lewis PhD^6, †^

^1^Department of Epidemiology, Harvard T.H. Chan School of Public Health, 02115, Boston, MA, USA

^2^Center for Communicable Disease Dynamics, Harvard T.H. Chan School of Public Health, 02115, Boston, MA, USA

^3^Department of Immunology and Infectious diseases, Harvard T.H. Chan School of Public Health, 02115, Boston, MA, USA

^4^Infectious Disease and Microbiome Program, Broad Institute of MIT & Harvard, Cambridge, MA, 02142, USA

^5^Population Health, University of Georgia, Athens, GA, 30602, USA

^6^Alberta Centre for Advanced Diagnostics, Department of Biological Sciences, University of Calgary, Calgary, Alberta, Canada

^7^Department of Pathology and Laboratory Medicine, Cumming School of Medicine, University of Calgary and Alberta Health Services

^8^Department of Medicine (Infectious Diseases), Cumming School of Medicine, University of Calgary and Alberta Health Services

^9^Infection Prevention and Control, Alberta Health Services, Calgary, Alberta, Canada

^10^Pharmacy Services, Alberta Health Services, Calgary, Alberta, Canada

^11^Department of Biomedical Engineering, Schulich School of Engineering, University of Calgary, Calgary, Alberta, Canada

^12^Department of Pathology and Laboratory Medicine, University of Calgary, Calgary, Alberta, Canada

^13^Alberta Precision Laboratories, Calgary, Alberta, Canada

^14^Department of Community Health Sciences, Cumming School of Medicine, University of Calgary and Alberta Health Services

^15^Department of Biological Sciences, University of Calgary, Calgary, Alberta, Canada

## Acknowledgments

The work described in this manuscript is a result of the intellectual, scientific, and foundational contributions from the following individuals: Gregory Tyrrell, Marcello Tonelli, Tanis Dingle, Michael Parkins, Otto G. Vanderkooi, Edward L. Huttlin, Joe Harrison, Diego Nobrega, Morgan Hepburn, Nadia Monych, Yeganeh Khaniani, Aja List, Jason Lam, Lindsey Paul, Nick Fitzgerald, Sean English, Jimmy Harold, Matthew Rogers, Owen Conroy, Colin MacKenzie, Racheal Apata, Madeline Kline. We thank Nic Van Bavel for proofreading the main text and providing valuable editorial suggestions.

