## Supplementary materials for "Trends in incidence and antimicrobial resistance for five major causes of bacteraemia in a Canadian metropolitan area, 2006–22: a genomic and antimicrobial use cohort study"

### S1 Data

#### S1.1 Study cohort

The sources for this study are clinical, microbiology, and genomic data from positive blood cultures classified as *Staphylococcus aureus, Enterococcus faecalis, Enterococcus faecium, Escherichia coli,* and *Klebsiella pneumoniae* between Jan 1, 2006 and Oct 14, 2022 in the Calgary Health Zone (CHZ). The area was a centralized health district consisting of 14 hospitals serving inpatient and outpatient health services to the residents of Calgary in Alberta, Canada.[1] Information on age and biological sex was available from birth certificates. We removed five isolates where information on age and biological sex was not available. We defined the pre-COVID-19 period from Jan 1, 2006 to Dec 31, 2019, and the COVID-19 period from Jan 1, 2020 to Oct 14, 2022.

#### S1.2 Antimicrobial prescribing data

Community antimicrobial prescribing data was obtained from Pharmaceutical Information Network and provided by Alberta Health Services as age-standardized unique dispensations per 100,000 Calgary residents.[2] Drug dispensations for antibiotics were extracted by Anatomical Therapeutic Chemical (ATC) code J01.[3] Outputs stratified by age and geography that were non-zero and have less than or equal to five (≤5) unique or total dispensations were not reported. Unique and total dispensations are estimated based on the definitions described by Alberta Health, Analytics and Performance Reporting Branch:

- Unique Dispensation: total people with at least one filled prescription of antimicrobials by ATC name stratified by age and AHS Zones
- Total Dispensations: Sum of all filled prescriptions for antimicrobials by ATC name (see Supplementary Information) stratified by age and AHS Zones.

Inpatient antimicrobial prescribing data was obtained from the electronic medical administration records and pharmacy system in the Calgary Zone of Alberta Health Services on all key antibiotic classes (aminoglycosides, carbapenems, fluoroquinolones, folate-path inhibitors, glycopeptides, lincosamides, macrolides, penicillins, tetracyclines, first, second, third, and fourth generation cephalosporins, carbapenems) and converted to Defined Daily Doses (DDDs) as previously described.[4]

**Table S1.** ATC classification codes for selected key antibiotic classes.

| Antibiotic Class | ATC Code |
|---|---|
| Aminoglycosides | J01GB |
| Carbapenems | J01DH |
| Fluoroquinolones | J01MA |
| Folate-pathway inhibitors | J01E |
| Glycopeptides | J01XA |
| Lincosamides | J01FF |
| Macrolides | J01FA |
| Penicillins | J01C |
| Tetracyclines | J01AA |
| 1st generation cephalosporins | J01DB |
| 2nd generation cephalosporins | J01DC |
| 3rd generation cephalosporins | J01DD |
| 4th generation cephalosporins | J01DE |

#### S1.3 Antimicrobial stewardship and Infection control protocol information

##### S1.3.1 MRSA policies

From 2006 to 2011, MRSA admission screening (nasal, rectal, and axillary/inguinal sites) was conducted for patients with a known history of MRSA, for all admissions to critical care units, and for patients directly transferred from a health care facility outside the Calgary Zone who had been hospitalized for more than 72 hours.

In 2011, following a prevalence survey, screening was expanded to include all medical and surgical admissions, in addition to the previously targeted groups. Patients identified as MRSA positive were placed on contact precautions (gown and gloves for all patient or environmental contact, and placement in a single room or designated bed space if available).

In 2015, universal screening was discontinued and replaced with a risk-based approach at admission. Screening was then limited to individuals who had been hospitalized for more than 24 hours in the prior six months, those receiving haemodialysis, or those with a history of incarceration.

##### S1.3.2 VRE policies

From 2006 to 2011, routine admission screening for vancomycin-resistant Enterococci (VRE) by rectal swab was performed for patients with a known history of VRE, all admissions to critical care units, and patients transferred from health care facilities outside the Calgary Zone after hospital stays longer than 72 hours. In 2011, screening was expanded to include all medical and surgical admissions. Patients identified as VRE positive were placed on contact precautions (gown and gloves for all patient or environmental contact, and a single room or designated bed space if available).

In 2015, screening protocols were revised to discontinue routine admission screening, restricting testing to high-risk units, specifically critical care, vascular surgery, and bone marrow transplant wards. Screening on these units continued variably until early 2021, when it was fully discontinued. In June 2017, contact precautions for patients colonized with VRE were discontinued in the renal medicine and transplant units at Foothills Medical Centre, Calgary. By 2021, contact precautions for all VRE-positive patients were fully withdrawn, with precautions thereafter applied only on a case-by-case basis.

##### S1.3.3 ESBL-GNB policies

Throughout the study period, no routine or targeted screening for ESBL-producing Gram-negative bacteria (ESBL-GNBs) was conducted in the Calgary Health Zone. Identification occurred only through clinical cultures obtained as part of routine care. When ESBL GNB were detected, antimicrobial susceptibility test results were reviewed to determine whether isolates demonstrated resistance across three or more antimicrobial classes (e.g., aminoglycosides, β-lactams, and fluoroquinolones). Such extensive multidrug resistance was uncommon. Consequently, the use of enhanced infection prevention measures, such as contact precautions or placement in single rooms, was infrequent for ESBL GNB during the study period.

##### S1.3.4 CPO policies

No active screening for carbapenemase-producing organisms (CPOs) was implemented prior to September 29, 2015. From that date onward, adult and pediatric patients admitted to acute care facilities were assessed for CPO screening eligibility based on defined risk criteria, primarily recent hospitalization (>24 hours) or receipt of hemodialysis outside of Canada within the preceding six months. Screening typically involved rectal swabs, with additional sampling of wounds, urine, or sputum when clinically indicated. CPOs remained rare, and the Calgary Health Zone continued to be a low-prevalence setting over the study period, consistent with findings from local prevalence assessments. Pre-emptive isolation was not used for inpatients identified as being at risk; contact precautions in a private room were implemented only after laboratory confirmation. For outpatients colonized with CPOs, additional precautions were generally not applied, except in oncology and hematology settings.

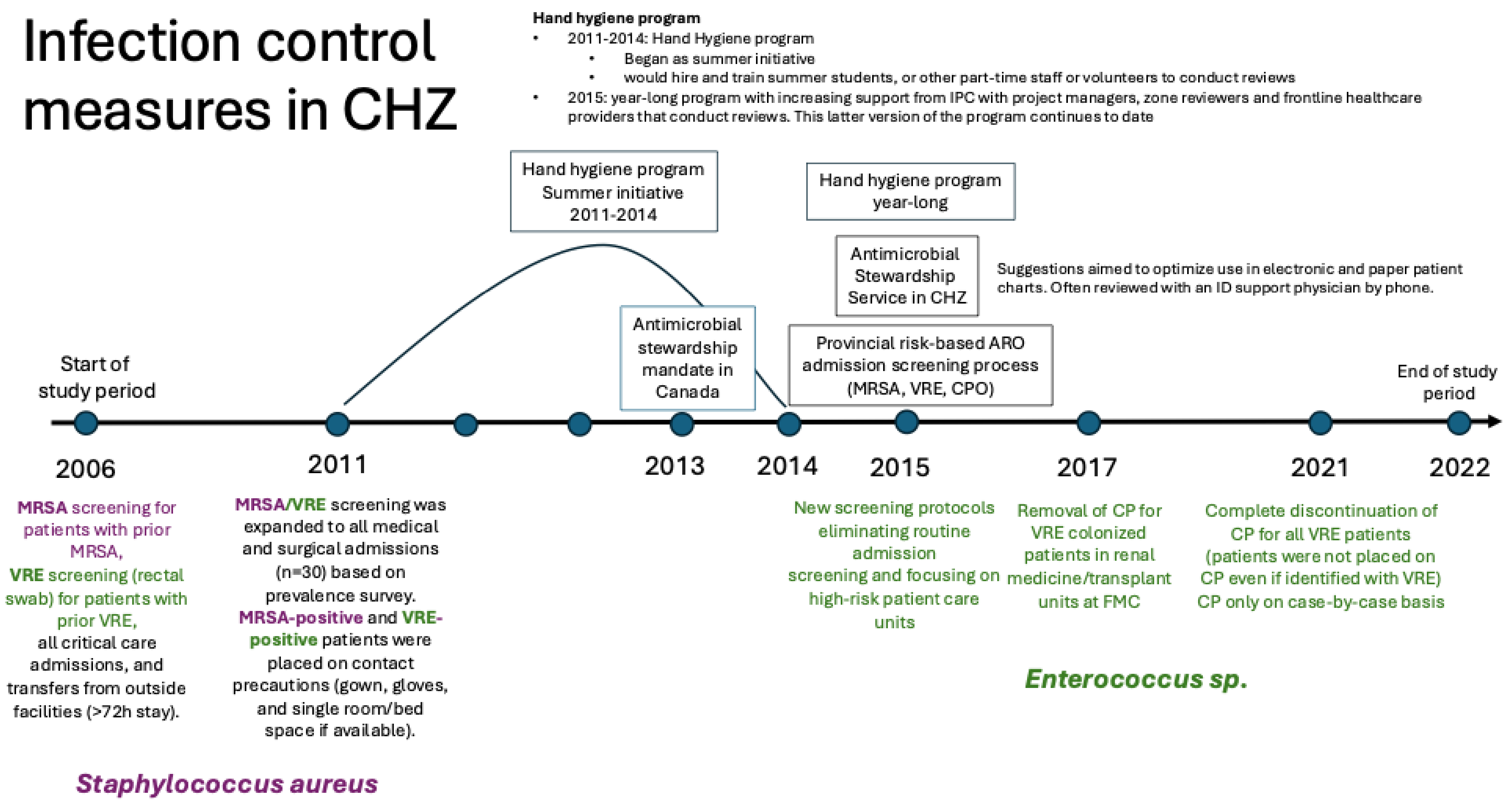

**Figure S 1 Timeline of major infection prevention, screening, and antimicrobial stewardship measures targeted for *Staphylococcus aureus* and *Enterococcus* species in the Calgary Health Zone (CHZ), 2006–2022. The figure summarizes and visualizes key system-level infection control interventions implemented during the study period, including hand hygiene initiatives, antimicrobial stewardship activities, and organism-specific screening and contact precaution policies.**

##### S1.3.5 Hand hygiene compliance

The Alberta Health Services IPC Hand Hygiene Program was established in the Calgary Health Zone in 2011, first established in the Calgary Health Zone in 2011, evolving from a summer student initiative and early digital tools into a province-wide system with dedicated staff, standardized training, and integration into AHS reporting and accreditation requirements. Infrastructure improvements, such as new sinks and product installations, were introduced in 2012, and by 2014 the program expanded with dedicated project managers and coordinators in each zone. In 2016, program leadership was restructured, with data collection and reporting centralized through the Clean Hands platform. The program provides a structured framework to improve and monitor hand hygiene compliance as a key patient safety measure, with the overarching goal of reducing healthcare-associated infections through consistent, proper practices. Accountability is embedded at multiple levels, local (front-line staff and site-based reviewers), zone and provincial programs (project managers, directors, and committees), and executive leadership. Compliance is assessed through direct observations based on the "Four Moments for Hand Hygiene," with data captured in the Clean Hands System (iPad app, paper tool, and portal). Results are reported at the unit, site, zone, and provincial levels, against a provincial target of 90% compliance. Education, competency checks, and quality assurance processes ensure data reliability, while resource allocation is guided by performance and planning metrics. More details can be found in the Infection Prevention and Control Hand Hygiene Program guide provided by Alberta Health Services.[5]

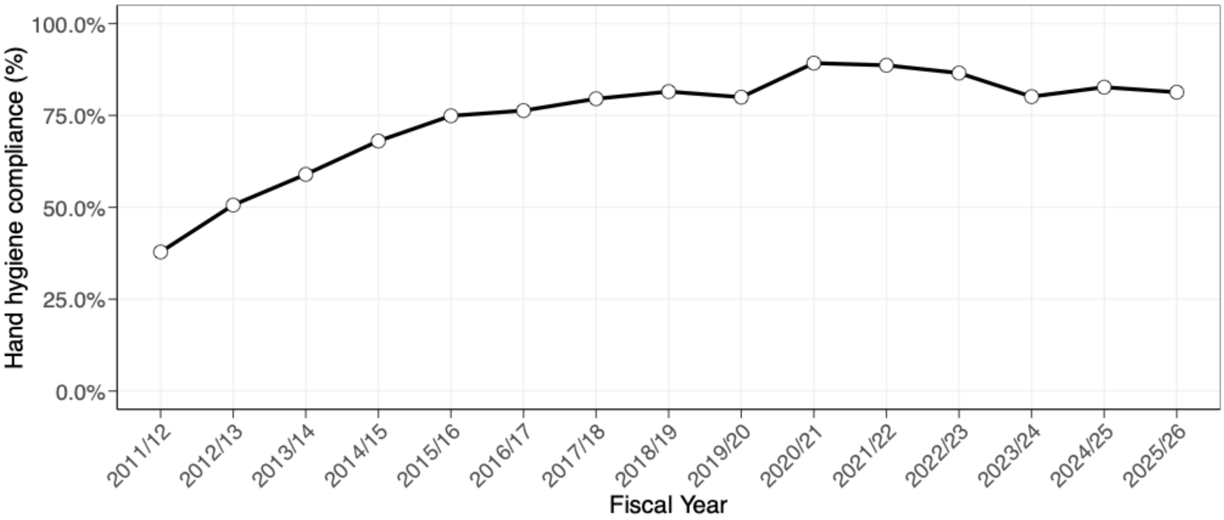

**Figure S2. Hand hygiene compliance in the Calgary Health Zone, fiscal years 2011/12–2025/26. Compliance was calculated using the number of divided by the total number of observations.**

##### S1.3.6 Antimicrobial Stewardship in the Calgary Health Zone

The Antimicrobial Stewardship Program (ASP) in the Calgary Health Zone has been active since 2002, making it one of the earlier established programs in western Canada.[6] The program was staffed by 2 FTE infectious diseases pharmacists and a 0.3 FTE physician leader, and was initially centred at Foothills Medical Centre (FMC), the largest hospital in Alberta, with outreach extending to all other adult acute care sites in the zone, including the Peter Lougheed Centre, Rockyview General Hospital, and South Health Campus. Key interventions included standardized antimicrobial order sets, cascade susceptibility reporting, prospective audit and feedback, and educational sessions for prescribers, all aimed at promoting optimal antimicrobial use. An active Stewardship Committee provided multidisciplinary oversight, including microbiology input. The Calgary program later helped inform the development of the broader Alberta Health Services provincial stewardship infrastructure, which was formalized with the creation of the provincial Antimicrobial Stewardship Committee in 2011.

### S2 Statistical analysis

#### S2.1 Segmented regression analyses

To estimate time trends and changes in trends for incidence of infections we performed segmented regression models using the segmented package in R.[7,8] Since the package did not support negative-binomial regression, we performed segmented Poisson regression. We selected the number of breakpoints using the *selgmented* and *davies.test* functions according to the Bayesian Information Criterion (BIC) and using the Bonferroni correction. In cases where the model with no break points showed the best fit according to BIC, we performed negative-binomial regression to account for overdispersion.

#### S2.2 Comparing proportions

We compared proportions of cases in different age groups in different sequence types using the Z-test and the function prop.test in R. We performed one-sided tests, depending on the comparison of the age groups.

#### S2.3 Shannon diversity index

We used the Shannon diversity index [9] to estimate the diversity of strains for each organism overall and for each phenotype within each organism, and was calculated as follows:

$$H' = -\sum_{i=1}^{R} p_i \ln p_i$$

where $R$ is the number of different strains (strainGST reference clusters) the dataset of interest contains. The calculations were performed using the R package vegan.

### S3 Resistance phenotypes

Phenotypes were defined using pathogen-specific panels of first- and second-line antibiotics, referred to as key antibiotics. For each target organism, we report the most common phenotype, which accounted for at least 95% of all 30-day index isolates.

#### S3.1 *Staphylococcus aureus*

**Table S2. Most common phenotypes for *Staphylococcus aureus.* Antimicrobial susceptibility test (AST) results were used to identify whether a culture was resistant to a specific antibiotic. Interpretation of ASTs was based on the reported minimum inhibitory concentrations and categorised into susceptible (S), intermediate (I), or resistant (R). We grouped intermediate and resistant in our analyses. We report the most common phenotypes covering at least 95% of index isolates.**

| Cloxacillin | Ciprofloxacin | Erythromycin | Clindamycin | Number of 30-day index isolates | Percentage (%) | Cumulative percentage (%) |
|---|---|---|---|---|---|---|
| S | S | S | S | 4543 | 62.5 | 62.5 |
| S | S | R | R | 734 | 10.1 | 72.6 |
| R | R | R | R | 516 | 7.1 | 79.7 |
| R | R | R | S | 440 | 6.1 | 85.7 |
| S | R | S | S | 320 | 4.4 | 90.1 |
| R | S | S | S | 141 | 1.9 | 92.1 |
| R | R | S | S | 123 | 1.7 | 93.8 |
| S | R | R | R | 110 | 1.5 | 95.3 |

#### S3.2 *Enterococcus faecalis*

**Table S3. Most common phenotypes for *Enterococcus faecalis.* Antimicrobial susceptibility test (AST) results were used to identify whether a culture was resistant to a specific antibiotic. Interpretation of ASTs was based on the reported minimum inhibitory concentrations and categorised into susceptible (S), intermediate (I), or resistant (R). We grouped intermediate and resistant in our analyses. We report the most common phenotypes covering at least 95% of index isolates.**

| Tetracycline | Gentamicin | Ciprofloxacin | Number of 30-day index isolates | Percentage (%) | Cumulative percentage (%) |
|---|---|---|---|---|---|
| R | S | S | 748 | 46.4 | 46.4 |
| S | S | S | 352 | 21.8 | 68.3 |
| R | R | S | 165 | 10.2 | 78.5 |
| R | R | R | 157 | 9.7 | 88.3 |
| R | S | R | 109 | 6.8 | 95.0 |
| S | S | R | 26 | 1.6 | 96.6 |

#### S3.3 *Enterococcus faecium*

**Table S4. Most common phenotypes for *Enterococcus faecium.* Antimicrobial susceptibility test (AST) results were used to identify whether a culture was resistant to a specific antibiotic. Interpretation of ASTs was based on the reported minimum inhibitory concentrations and categorised into susceptible (S), intermediate (I), or resistant (R). We grouped intermediate and resistant in our analyses. We report the most common phenotypes covering at least 95% of index isolates.**

| Vancomycin | Tetracycline | Gentamicin | Number of 30-day index isolates | Percentage (%) | Cumulative percentage (%) |
|---|---|---|---|---|---|
| S | S | S | 312 | 39.0 | 39.0 |
| R | S | S | 137 | 17.1 | 56.1 |
| S | R | S | 136 | 17.0 | 73.0 |

| | | | | | |
|---|---|---|---|---|---|
| S | R | R | 103 | 12.9 | 85.9 |
| R | R | R | 38 | 4.7 | 90.6 |
| R | R | S | 37 | 4.6 | 95.3 |
| S | S | R | 17 | 2.1 | 97.4 |

#### S3.4 *Escherichia coli*

**Table S5. Most common phenotypes for *Escherichia coli.* Antimicrobial susceptibility test (AST) results were used to identify whether a culture was resistant to a specific antibiotic. Interpretation of ASTs was based on the reported minimum inhibitory concentrations and categorised into susceptible (S), intermediate (I), or resistant (R). We grouped intermediate and resistant in our analyses. We report the most common phenotypes covering at least 95% of index isolates.**

| Cefazolin | Ceftriaxone | Ciprofloxacin | Gentamicin | Trimethoprim/ Sulfamethoxazole | Number of 30-day index isolates | Percentage (%) | Cumulative percentage (%) |
|---|---|---|---|---|---|---|---|
| S | S | S | S | S | 5214 | 45.9 | 45.9 |
| S | S | S | S | R | 857 | 7.5 | 53.4 |
| S | S | R | S | S | 702 | 6.2 | 59.6 |
| R | S | S | S | S | 640 | 5.6 | 65.2 |
| R | R | R | R | R | 487 | 4.3 | 69.5 |
| R | R | R | S | R | 482 | 4.2 | 73.8 |
| R | S | S | S | R | 425 | 3.7 | 77.5 |
| S | S | R | S | R | 407 | 3.6 | 81.1 |
| R | R | R | S | S | 327 | 2.9 | 84.0 |
| R | R | R | R | S | 251 | 2.2 | 86.2 |
| R | S | R | S | S | 222 | 2.0 | 88.1 |
| R | S | R | S | R | 196 | 1.7 | 89.9 |
| S | S | R | R | R | 156 | 1.4 | 91.2 |
| R | R | S | S | S | 154 | 1.4 | 92.6 |
| S | S | S | R | R | 110 | 1.0 | 93.5 |
| R | S | R | R | R | 106 | 0.9 | 94.5 |
| R | R | S | S | R | 81 | 0.7 | 95.2 |

#### S3.5 *Klebsiella pneumoniae*

**Table S6. Most common phenotypes for *Klebsiella pneumoniae.* Antimicrobial susceptibility test (AST) results were used to identify whether a culture was resistant to a specific antibiotic. Interpretation of ASTs was based on the reported minimum inhibitory concentrations and categorised into susceptible (S), intermediate (I), or resistant (R). We grouped intermediate and resistant in our analyses. We report the most common phenotypes covering at least 95% of index isolates.**

| Cefazolin | Ceftriaxone | Ciprofloxacin | Gentamicin | Piperacillin/ Tazobactam | Trimethoprim/ Sulfamethoxazole | Number of 30-day index isolates | Percentage (%) | Cumulative percentage (%) |
|---|---|---|---|---|---|---|---|---|
| S | S | S | S | S | S | 1968 | 79.8 | 79.8 |
| S | S | S | S | S | R | 97 | 3.9 | 83.7 |
| R | S | S | S | S | S | 74 | 3.0 | 86.7 |
| S | S | R | S | S | S | 48 | 1.9 | 88.7 |
| S | S | R | S | S | R | 29 | 1.2 | 89.8 |
| R | R | R | R | R | R | 25 | 1.0 | 90.8 |
| R | R | R | R | S | R | 21 | 0.9 | 91.7 |
| R | S | S | S | R | S | 17 | 0.7 | 92.4 |

| | | | | | | | | |
|---|---|---|---|---|---|---|---|---|
| R | R | R | S | S | R | 16 | 0.6 | 93.0 |
| R | S | S | S | S | R | 11 | 0.4 | 93.5 |

### S4 Genomic analyses

#### S4.1 Assembly Pipeline

Whole-genome sequencing was performed at the Broad Institute of MIT and Harvard for all isolates. Sequencing libraries were prepared using the Nextera XT DNA Library Prep Kit (Illumina, San Diego, CA, USA), and 151 bp paired-end reads were generated using the Illumina HiSeqX platform. Reads were processed and quality-controlled using the seQuoia computational pipeline (https://github.com/broadinstitute/seQuoia). Adaptors were trimmed using TrimGalore (v 0.6.5). Draft assemblies were generated for each isolate using Unicycler v0.4.6 with SPAdes v3.13.0 [10] and Pilon v1.23 correction.[11] Assemblies were annotated using Prokka v1.14.6.[10] Species-level taxonomic assignments were determined using Centrifuge v1.0.4_beta as a quality control measure.[12] Each isolate was matched to a closely related reference genome using StrainGST from the Strain Genome Explorer (StrainGE) toolkit v1.3.1. Variant calling and quantification of genomic differences were performed with Pilon v1.23.[11]

#### S4.2 Quality Control

We assessed assembly quality using several metrics based on read quality, assembly length, assembly completeness, and strain composition with species-specific thresholds (Table S2). Additionally, we identified contaminated runs as those with >1% of Centrifuge hits belonging to a different genus than the target organism or >5% of centrifuge hits belonging to an organism of the same genus but different species as the target organism.

**Table S7. Assembly quality metrics. SA = Staphylococcus aureus. EFM = Enterococcus faecium, EFS = Enterococcus faecalis, EC = Escherichia coli, KP = Klebsiella pneumoniae.**

| Metric | SA | EFM | EFS | EC | KP |
|---|---|---|---|---|---|
| **Genome Size (+/- 20%)** | 2730000 | 2840000 | 2960000 | 5000000 | 5390000 |
| **GC Content (+/- 10%)** | 36.0 | 40.6 | 39.9 | 52.4 | 58.5 |
| **Annotated genes (+/- 20%)** | 2530 | 2740 | 2840 | 4700 | 5000 |
| **Maximum assembly contigs** | 200 | 300 | 200 | 300 | 300 |
| **Minimum assembly contig N50** | 25000 | 15000 | 25000 | 25000 | 25000 |
| **Minimum sequencing coverage** | 20 | 20 | 20 | 20 | 20 |
| **Minimum trimmed read length** | 120 | 120 | 120 | 120 | 120 |
| **Minimum mean base quality** | 30 | 30 | 30 | 30 | 30 |
| **Max StrainGST strains (score>0.2)** | 1 | 1 | 1 | 1 | 1 |

#### S4.3 Typing

##### S4.3.1 StrainGST

StrainGST typing consists of two steps. First, we used the reference database creation pipeline for each for the four genera under study. This entails downloading and k-merizing all complete genomes for each genus from NCBI (as of May 2022 for *S. aureus*, June 2022 for *Enterococcus*, September 2022 for *E. coli*, and October 2022 for *K. pneumoniae*) and choosing representative genomes for each genus with a Jaccard similarity lower than 0.90 compared to other reference genomes. In the second step, we k-merized our sample reads and compared them to the genus-specific database generated in the first step. This identifies a reference most genetically similar to each sample. We used the default k-mer of 23.

##### S4.3.2 MLST

Identification of sequence types was confirmed with Ariba v2.14.5[13] and species-specific scheme databases of seven housekeeping genes. These databases were obtained from

- https://pubmlst.org/efaecalis
- https://pubmlst.org/efaecium
- https://pubmlst.org/escherichia
- https://bigsdb.pasteur.fr/klebsiella
- https://pubmlst.org/saureus.

##### S4.3.3 Identification of antimicrobial resistance associated genes and alleles

We used AMRFinderPlus v 3.12.8 with database version 2024-05-02.2 to identify the presence of previously described variants and genes associated with antimicrobial resistance.[14] For each species, we used an organism specific database specified by the --organism flag.

###### *S4.3.3.1* Concordance of antimicrobial resistance genotypes and phenotypes in *Staphylococcus aureus*

The concordance between antimicrobial resistance genotypes and phenotypes in *Staphylococcus aureus* was generally high, indicating that genetic determinants were strong predictors of observed resistance profiles. For most antimicrobial classes, the presence of resistance-associated genes correlated well with phenotypic resistance determined by susceptibility testing (Table S8).

**Table S8. *Staphylococcus aureus* antimicrobial susceptibility genotype and phenotype summary.**

| Antibiotic | Proportion of isolates with no AMR markers that are phenotypically susceptible | Proportion of phenotypically resistant isolates with at least one AMR marker |
|---|---|---|
| Cloxacillin | 0.997 | 0.989 |
| Ciprofloxacin | 0.997 | 0.996 |
| Clindamycin | 0.974 | 0.892 |
| Erythromycin | 0.981 | 0.959 |

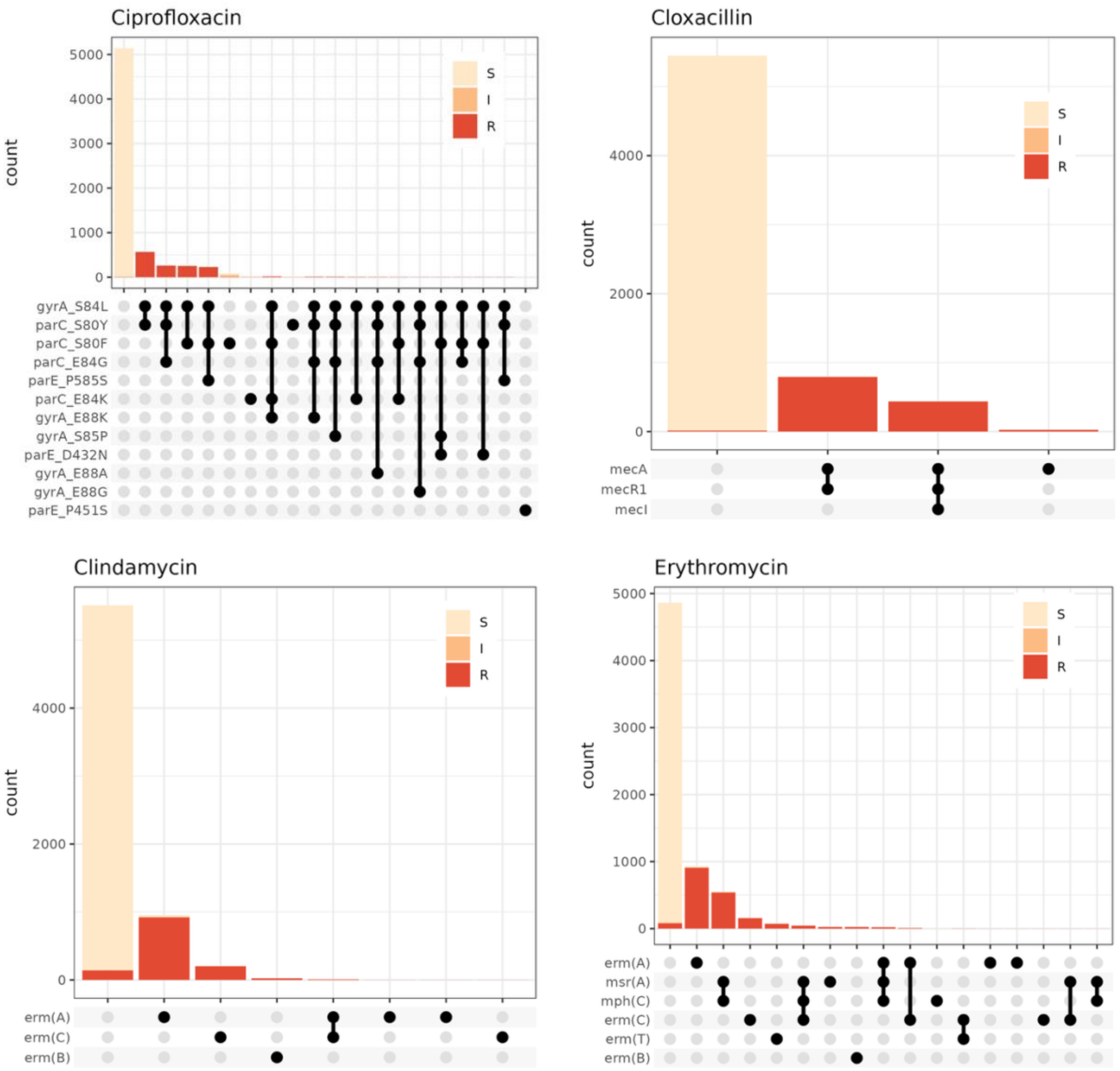

**Figure S3.** ***Staphylococcus aureus*** **antimicrobial resistance genotypes and phenotypes. The presence (black) or absence (gray) of known antimicrobial resistance determinants was identified with AMRFinderPlus. The stacked bar charts represent the number of isolates sequenced with each genotype that were phenotypically susceptible (beige), intermediate (orange), or resistant (red) to each antimicrobial.**

##### *S4.3.3.2* Concordance of antimicrobial resistance genotypes and phenotypes in *Enterococcus faecalis*

**Table S9.** ***Enterococcus faecalis*** **antimicrobial susceptibility genotype and phenotype summary.**

| Antibiotic | Proportion of isolates with no AMR markers that are phenotypically susceptible | Proportion of phenotypically resistant isolates with at least one AMR marker |
|---|---|---|
| Ciprofloxacin | 0.976 | 0.975 |
| Gentamicin | 0.826 | 0.261 |
| Tetracycline | 0.966 | 0.989 |

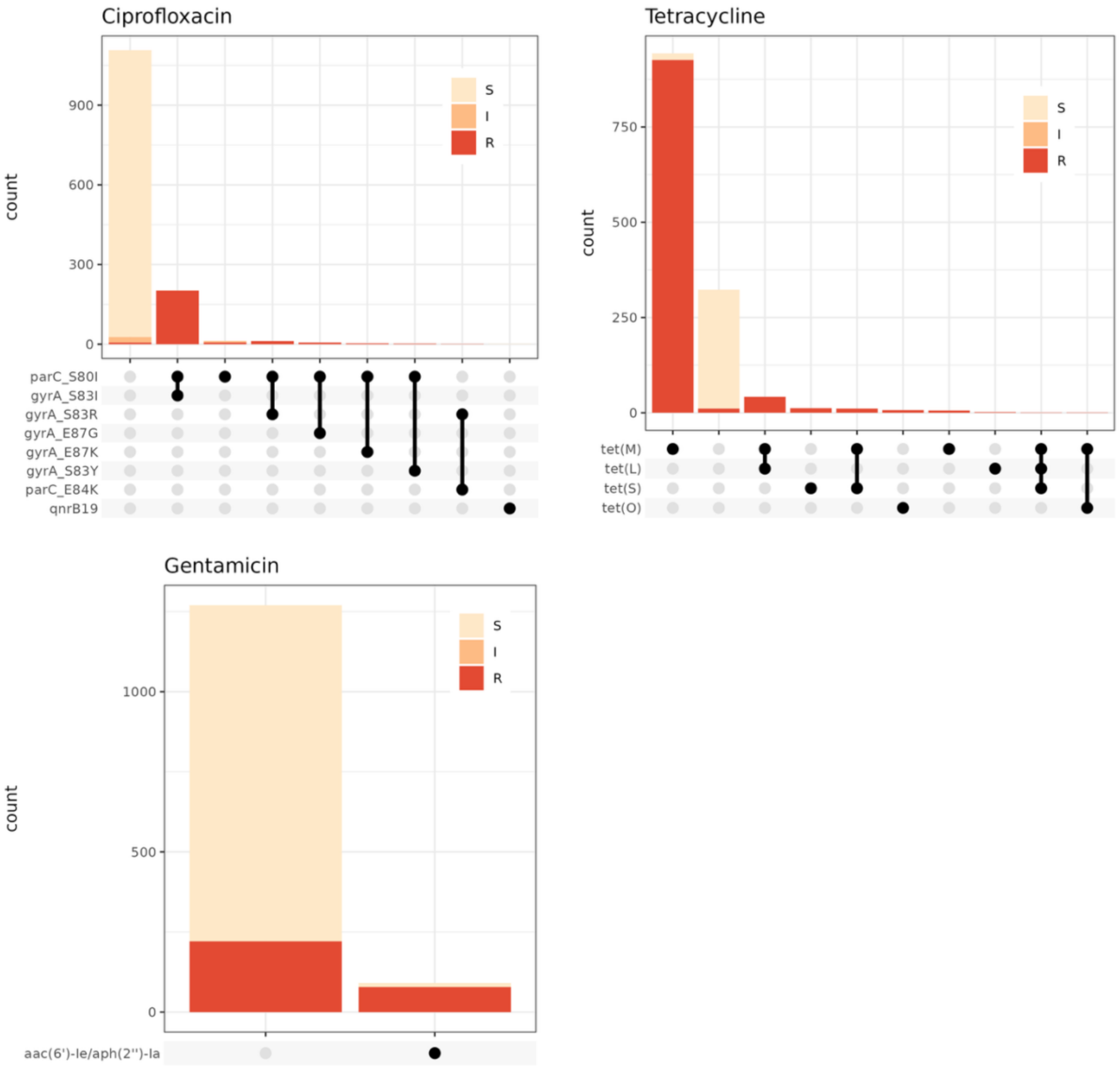

**Figure S4. *Enterococcus faecalis* antimicrobial resistance genotypes and phenotypes. The presence (black) or absence (gray) of known antimicrobial resistance determinants was identified with AMRFinderPlus. The stacked bar charts represent the number of isolates sequenced with each genotype that were phenotypically susceptible (beige), intermediate (orange), or resistant (red) to each antimicrobial.**

##### *S4.3.3.3* Concordance of antimicrobial resistance genotypes and phenotypes in *Enterococcus faecium*

**Table S10. *Enterococcus faecium* antimicrobial susceptibility genotype and phenotype summary.**

| Antibiotic | Proportion of isolates with no AMR markers that are phenotypically susceptible | Proportion of phenotypically resistant isolates with at least one AMR marker |
|---|---|---|
| Ciprofloxacin | 0.753 | 0.981 |
| Gentamicin | 0.833 | 0.197 |
| Tetracycline | 0.994 | 0.991 |
| Vancomycin | 0.998 | 0.994 |

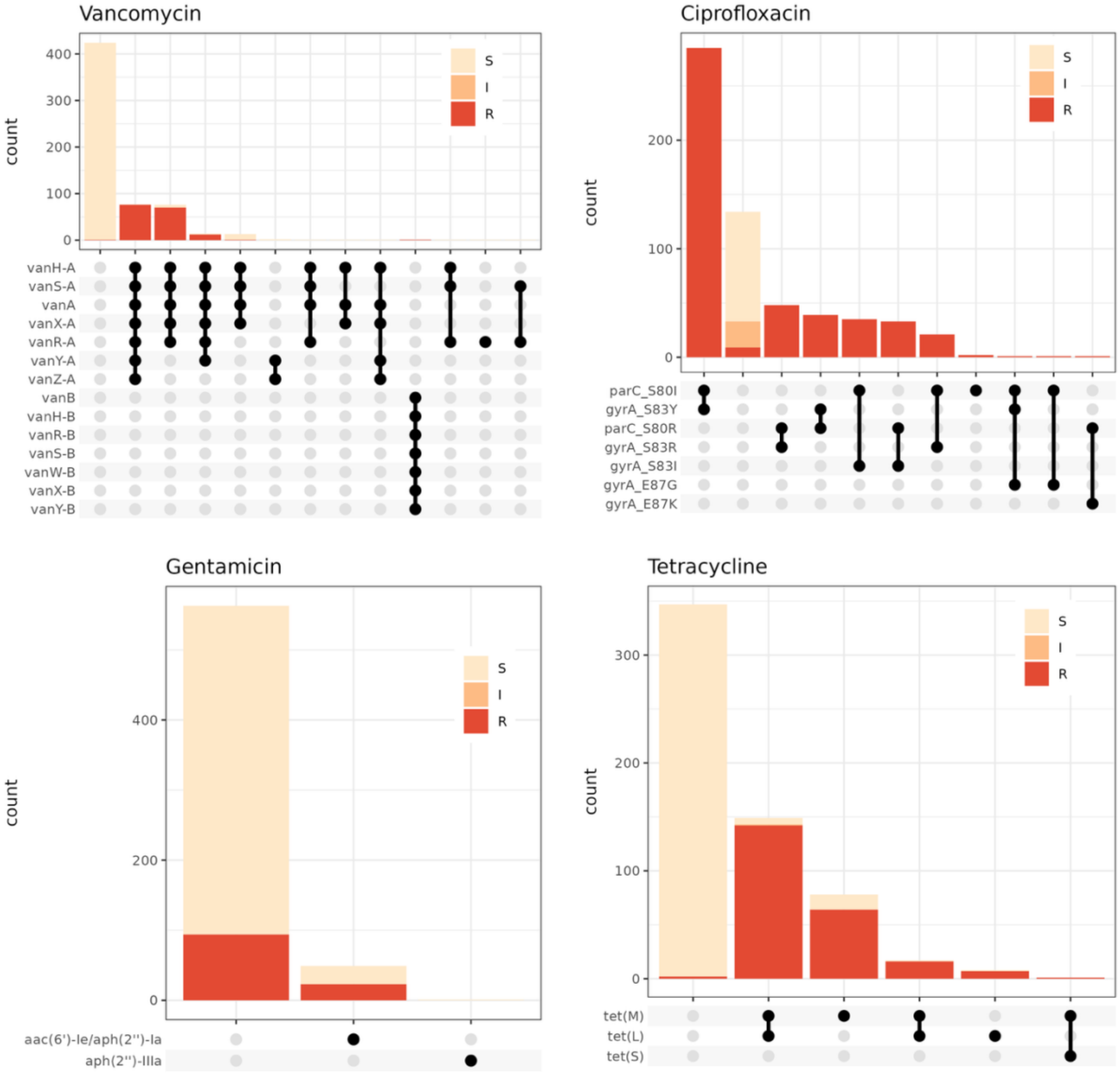

**Figure S5.** ***Enterococcus faecium*** **antimicrobial resistance genotypes and phenotypes. The presence (black) or absence (gray) of known antimicrobial resistance determinants was identified with AMRFinderPlus. The stacked bar charts represent the number of isolates sequenced with each genotype that were phenotypically susceptible (beige), intermediate (orange), or resistant (red) to each antimicrobial.**

###### S4.3.3.4 Assembly and identification of high-level gentamicin resistance determinants in *Enterococcus*

We observed an unexpectedly high number of *Enterococcus* isolates with phenotypic high-level gentamicin resistance that did not encode known determinants (i.e. *aac*(6′)Ie-*aph*(2″)Ia). *aac*(6′)Ie-*aph*(2″)Ia has been associated with Tn*5281* transposons in *Enterococcus.* Since repeat-containing mobile elements may be more difficult to assemble using short reads, we additionally investigated the presence of *aac*(6′)Ie-*aph*(2″)Ia using ARIBA v2.14.5 with the CARD database, which detects the presence of resistance-associated genes and alleles directly from reads rather than assembled genomes.

We compared the coverage of *aac*(6′)Ie-*aph*(2″)Ia detected by ARIBA to the median coverage across the genome using reads mapped to the closed StrainGST reference genome, which we will refer to as total coverage. We found that sequencing coverage of *aac*(6′)Ie-*aph*(2″)Ia is lower than other parts of the genome in *E. faecium*. Among *E. faecium* isolates with high-level gentamicin resistance and a copy of *aac*(6′)Ie-*aph*(2″)Ia in the genome assembly, the median coverage of *aac*(6′)Ie-*aph*(2″)Ia was 41.1% of total coverage. Additionally, among *E. faecium* isolates with high-level gentamicin resistance and without a copy of *aac*(6′)Ie-*aph*(2″)Ia in the genome assembly, *aac*(6′)Ie-*aph*(2″)Ia coverage from ARIBA ranged from 0%-53.3% (median: 20.1%) of total coverage.

We found similar results for *E. faecalis*. Among isolates with high-level gentamicin resistance and *aac*(6′)Ie-*aph*(2″)Ia in the genome assembly, *aac*(6′)Ie-*aph*(2″)Ia had a median coverage of 27.5% of total coverage, suggesting that sequencing coverage of *aac*(6′)Ie-*aph*(2″)Ia is low compared to other parts of the genome in this species as well. Among isolates with high-level gentamicin resistance and without a copy of *aac*(6′)Ie-*aph*(2″)Ia in the genome assembly, *aac*(6′)Ie-*aph*(2″)Ia coverage from ARIBA ranged from 0%-47.1% (median: 14.6%) of total coverage.

In addition to gentamicin-resistant isolates that did not encode *aac*(6′)Ie-*aph*(2″)Ia, we also identified susceptible isolates with *aac*(6′)Ie-*aph*(2″)Ia in both *E. faecium* and *E. faecalis* using AMRFinderPlus (Figure S5-S6) and ARIBA. ARIBA identified *aac*(6′)Ie-*aph*(2″)Ia in 33.9% (172/507) *E. faecium* susceptible to gentamicin, albeit at lower coverage (median: 13.8% of total coverage). In *E. faecalis*, ARIBA identified *aac*(6′)Ie-*aph*(2″)Ia in only 5.2% (55/1061) of gentamicin susceptible isolates (median: 13.3% of total coverage). Identification of this gene in susceptible genomes could be a result of read contamination across barcodes, changes in expression of *aac*(6′)Ie-*aph*(2″)Ia, or variation in MIC measurements around the breakpoint. Precise gentamicin MIC values were unavailable for these isolates, so we could not assess if susceptible isolates encoding *aac*(6′)Ie-*aph*(2″)Ia had MICs close to the breakpoint (≥500 µg/mL). While unknown genetic determinants of gentamicin resistance or susceptibility may be contributing to discordance between genotypes and phenotypes, we conclude that limitations of high-throughput short-read sequencing have also impacted the agreement between high-level gentamicin resistance phenotypes and the presence of *aac*(6′)Ie-*aph*(2″)Ia in our study, specifically low sequencing depth for *aac*(6′)Ie-*aph*(2″)Ia compared to other parts of *Enterococcus* genomes.

##### S4.3.3.5 Concordance of antimicrobial resistance genotypes and phenotypes in *Escherichia coli*

**Table S11. *Escherichia coli* antimicrobial susceptibility genotype and phenotype summary.**

| Antibiotic | Proportion of isolates with no AMR markers that are phenotypically susceptible | Proportion of phenotypically resistant isolates with at least one AMR marker |
|---|---|---|
| Ciprofloxacin | 0.998 | 0.999 |
| Ceftriaxone | 0.996 | 0.981 |
| Gentamicin | 0.995 | 0.974 |
| Trimethoprim-sulfamethoxazole | 0.996 | 0.993 |

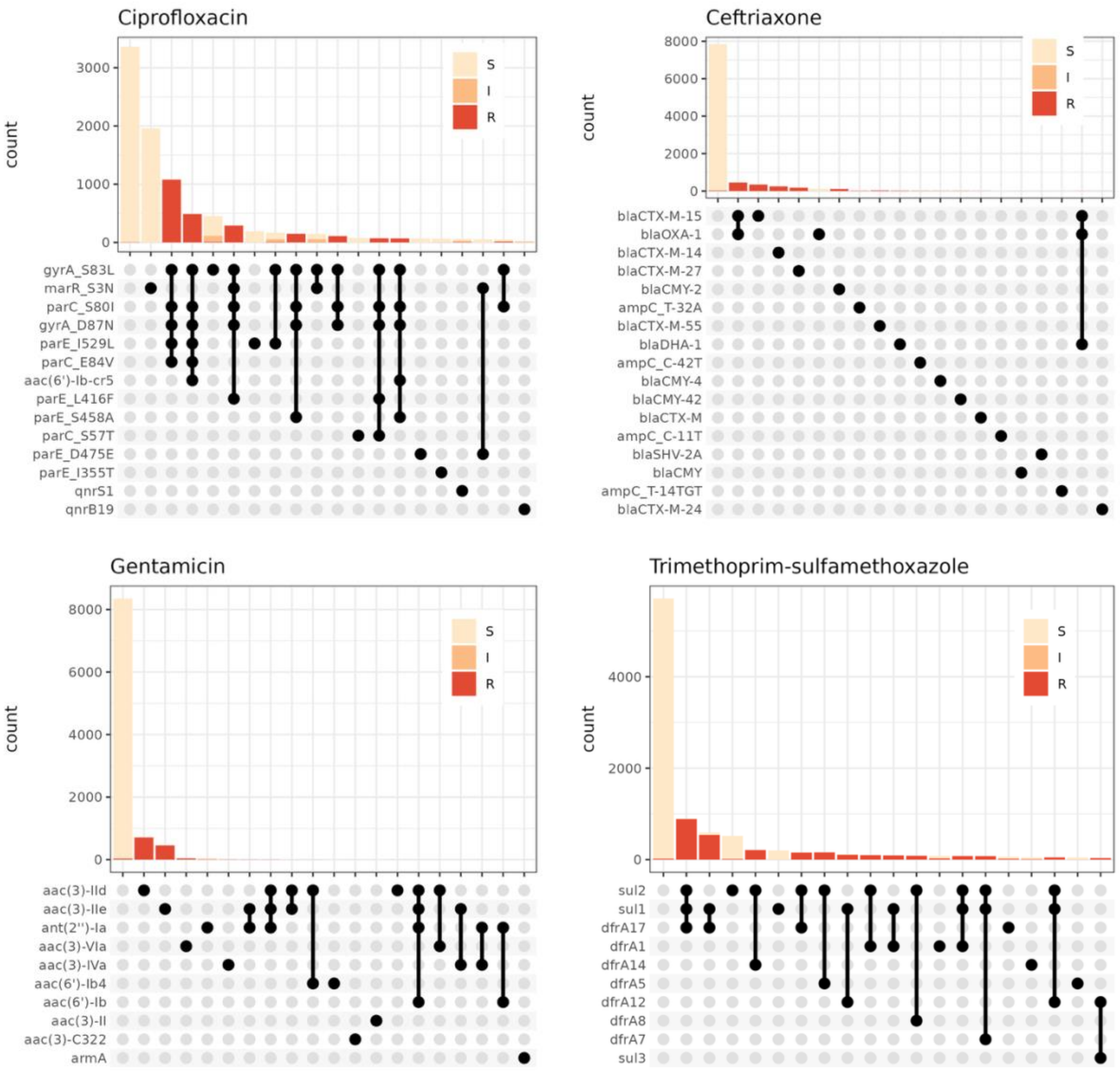

**Figure S6.** ***Escherichia coli*** **antimicrobial resistance genotypes and phenotypes. The presence (black) or absence (gray) of known antimicrobial resistance determinants was identified with AMRFinderPlus. The stacked bar charts represent the number of isolates sequenced with each genotype that were phenotypically susceptible (beige), intermediate (orange), or resistant (red) to each antimicrobial.**

##### S4.3.3.6 Concordance of antimicrobial resistance genotypes and phenotypes in *Klebsiella pneumoniae*

**Table S12.** ***Klebsiella pneumoniae*** **antimicrobial susceptibility genotype and phenotype summary.**

| Antibiotic | Proportion of isolates with no AMR markers that are phenotypically susceptible | Proportion of phenotypically resistant isolates with at least one AMR marker |
|---|---|---|
| Ciprofloxacin | 1.0 | 1.0 |
| Ceftriaxone | 0.991 | 0.892 |
| Gentamicin | 1.0 | 1.0 |
| Piperacillin-tazobactam | 0.977 | 0.970 |

| | | |
|---|---|---|
| Trimethoprim-sulfamethoxazole | 0.992 | 0.957 |

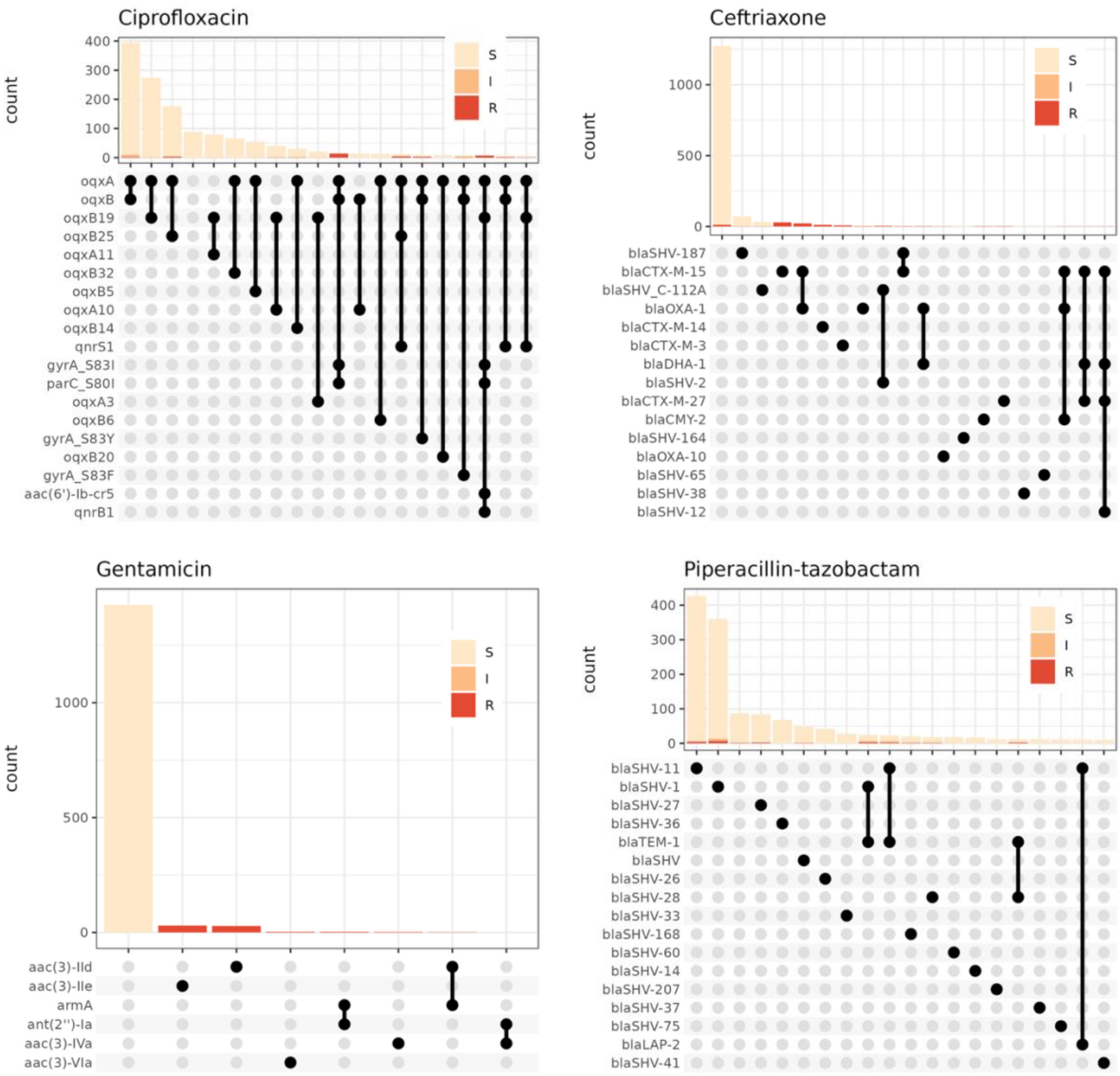

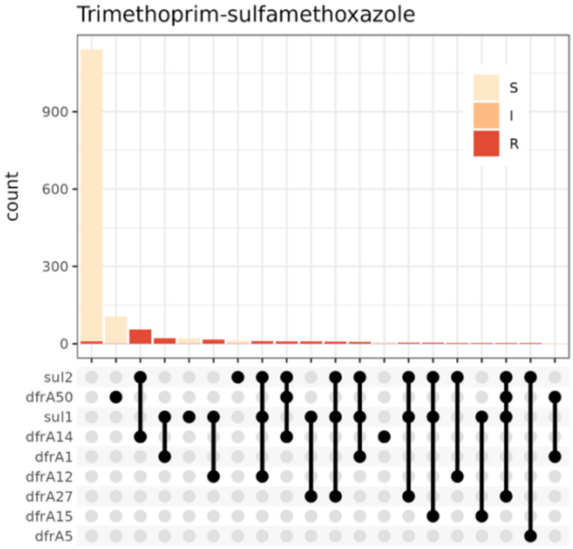

**Figure S7. *Klebsiella pneumoniae* antimicrobial resistance genotypes and phenotypes. The presence (black) or absence (gray) of known antimicrobial resistance determinants was identified with AMRFinderPlus. The stacked bar charts represent the number of isolates sequenced with each genotype that were phenotypically susceptible (beige), intermediate (orange), or resistant (red) to each antimicrobial.**

###### S4.3.3.7 Carbapenemases

Isolates encoding carbapenemases were rare in our dataset. Two *Escherichia coli* isolates encoded $bla_{OXA-181}$, collected in 2019 and 2020. Three isolates encoded $bla_{NDM-5}$, collected in 2016, 2018, and 2022, and one isolate encoded $bla_{NDM-1}$, collected in 2019. No *Klebsiella pneumoniae* isolates encoded a carbapenemase.

#### S4.4 Phylogenetics

For each bacterial lineage, we generated a core genome alignment to a reference genome with BWA v.0.7.18. We used Pilon v.1.24 to call SNPs and create a full-length genome alignment, masking positions with an allele frequency <0.9, mapping quality <10, or low coverage. The full-length alignment was used as input to Gubbins v.3.3.5 to remove recombination. The resulting core SNP alignment was further filtered to include only variable sites using SNP-sites v.2.5.1. The final clean alignment was used to build a maximum likelihood phylogenetic tree with IQTree v.2.0.6. We employed the ModelFinder algorithm to identify the most suitable nucleotide substitution model. Based on ModelFinder's results, we selected the general time reversible (GTR) model for nucleotide substitution along with the FreeRate model for rate heterogeneity, using three categories. Additionally, we utilized the UFBoot2 algorithm with 1,000 bootstrap replicates, as integrated in IQTree. The phylogenetic trees were then visualized through the online tool Interactive Tree of Life (IToL).

##### S4.5 Phylogenetic trees

###### S4.5.1 *Staphylococcus aureus*

In *S aureus,* the fully resistant phenotype (R-R-R-R) was predominantly observed within clonal complex CC5, which encompassed diverse antibiotic resistance phenotypes (Figure S8**Error! Reference source not found.**). Specifically, the R-R-R-R isolates clustered into two phylogenetically distinct sublineages, corresponding to sequence types ST225 and ST105. Conversely, isolates from ST5 were largely susceptible to all or most key antibiotic classes, underscoring significant phenotypic variation within CC5. Community-onset and hospital-onset cases appeared interspersed throughout the phylogeny, indicating no clear clustering by bacteremia onset.

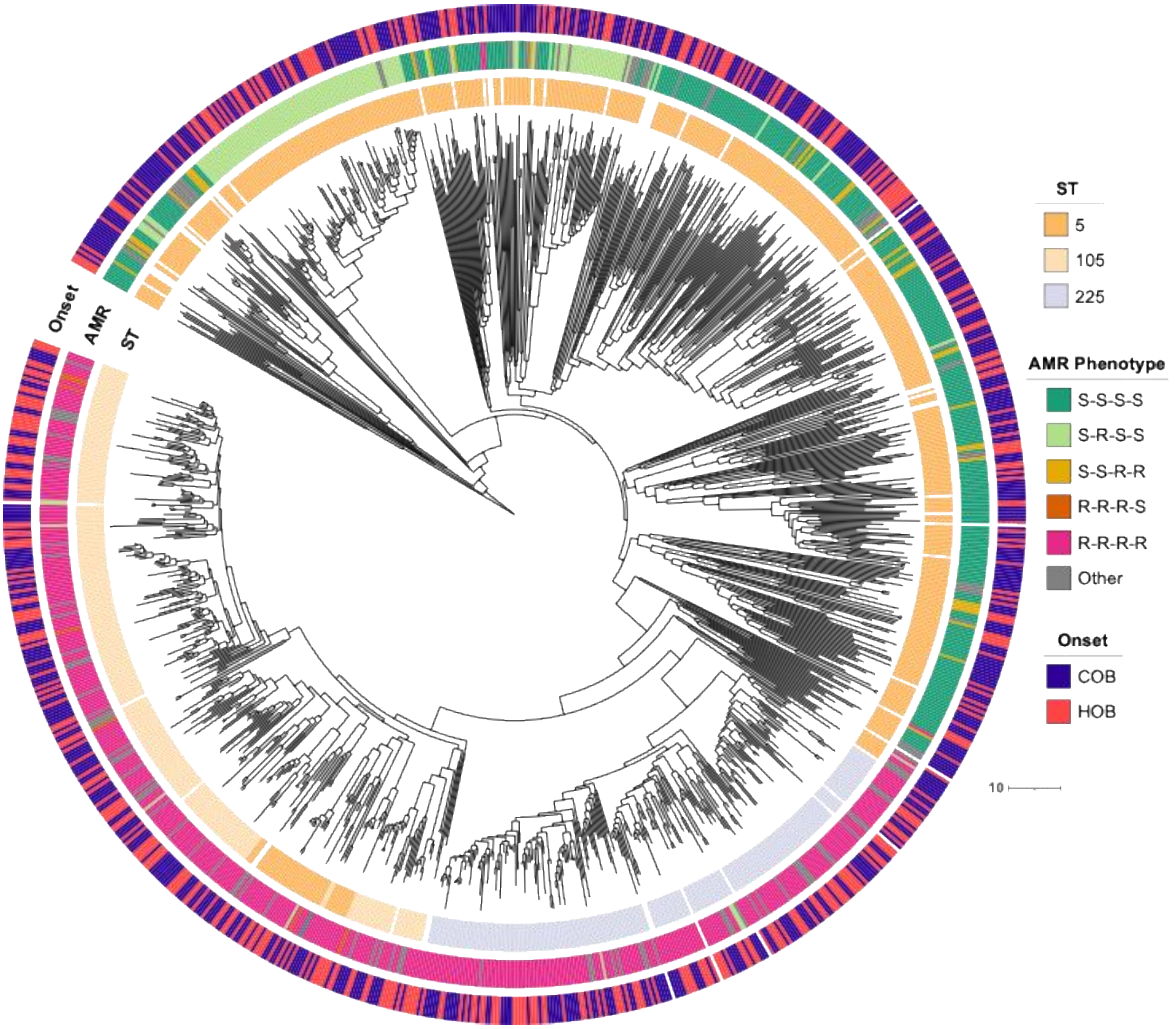

**Figure S8. Phylogeny of clonal complex 5 isolates from Calgary. Recombination-removed whole-genome phylogenetic tree of all isolates assigned to *S. aureus* StrainGST cluster 2. Inner ring shows sequence type (ST) representation. Second ring shows AMR phenotypes. Outer ring shows hospital-onset bacteremia (HOB) vs. community-onset bacteremia (COB).**

##### S4.5.2 *Enterococcus faecalis*

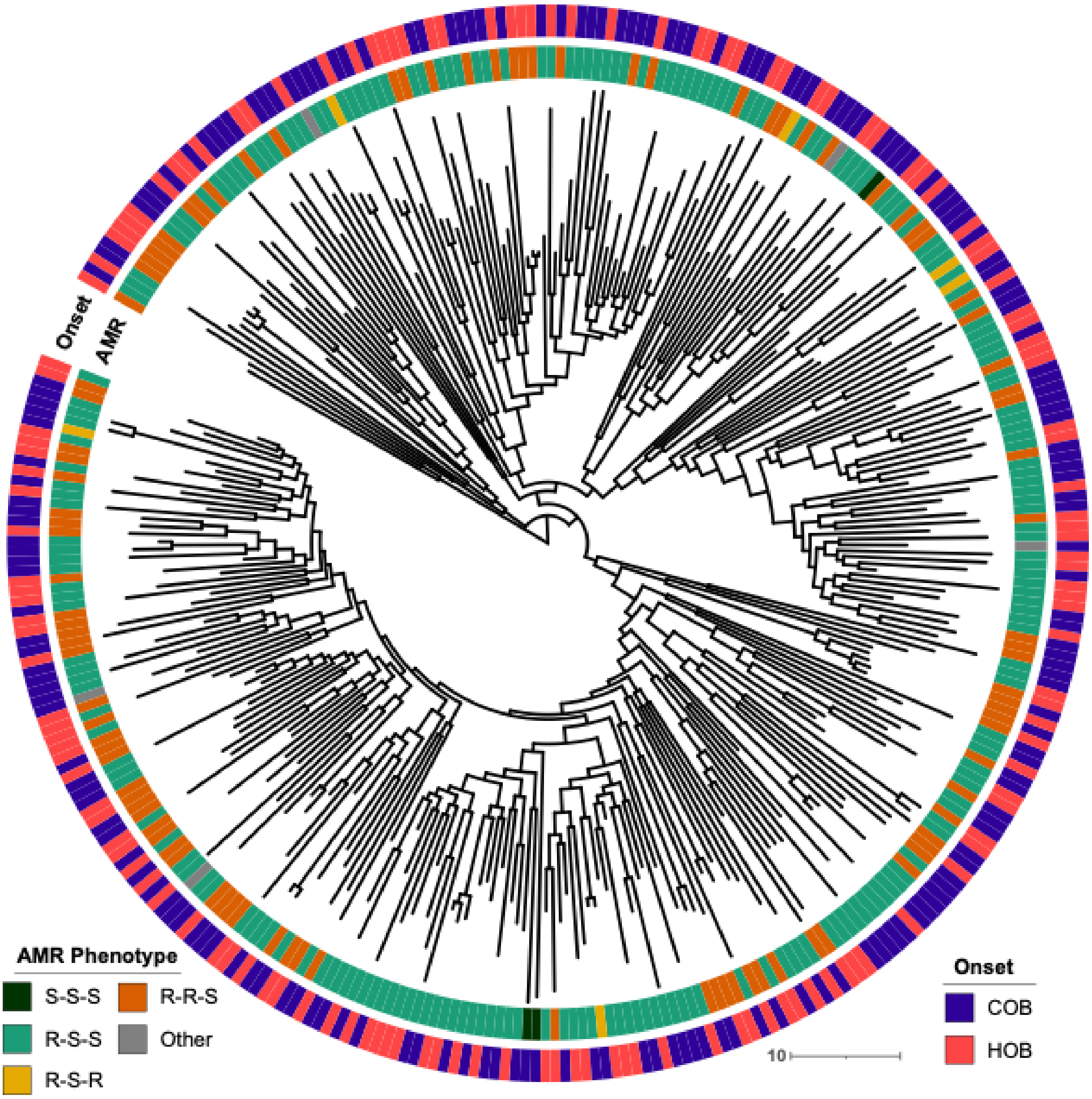

**Figure S9. Phylogenetic tree for *Enterococcus faecalis* ST179. Recombination-removed whole-genome phylogenetic tree of all isolates assigned to *E. faecalis* StrainGST cluster 1, which represents all ST179 isolates. Inner ring shows AMR phenotypes. Outer ring shows hospital-onset bacteremia (HOB) vs. community-onset bacteremia (COB).**

#### S4.5.3 *Enterococcus faecium*

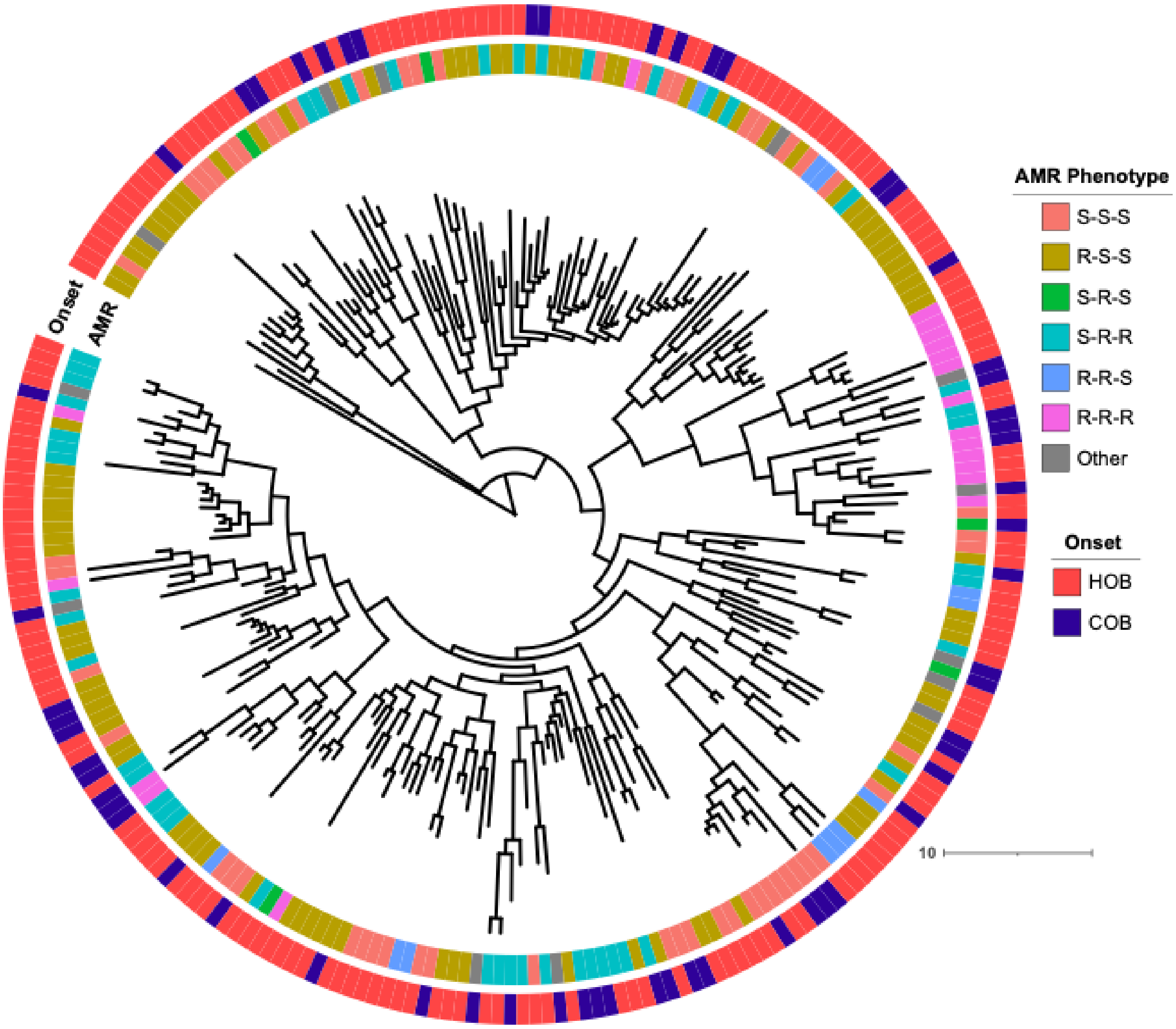

**Figure S10. Phylogenetic tree for *Enterococcus faecium* ST117. Recombination-removed whole-genome phylogenetic tree of all isolates assigned to *E. faecium* StrainGST cluster 1, which broadly represents sequence type 117 and closely related uncommon sequence types. Inner ring shows AMR phenotypes. Outer ring shows hospital-onset bacteremia (HOB) vs. community-onset bacteremia (COB).**

#### S4.5.4 *Escherichia coli*

In *E coli*, the predominant sequence type was ST131. We constructed a recombination-filtered whole-genome phylogenetic tree for all isolates assigned to *E coli* StrainGST cluster 1, corresponding to ST131 Clade C. The phylogeny shows no distinct clustering of hospital-onset versus community-onset bacteremia cases. Most ST131 isolates exhibited resistance to either ceftriaxone or ciprofloxacin, highlighting widespread antimicrobial resistance within this lineage.

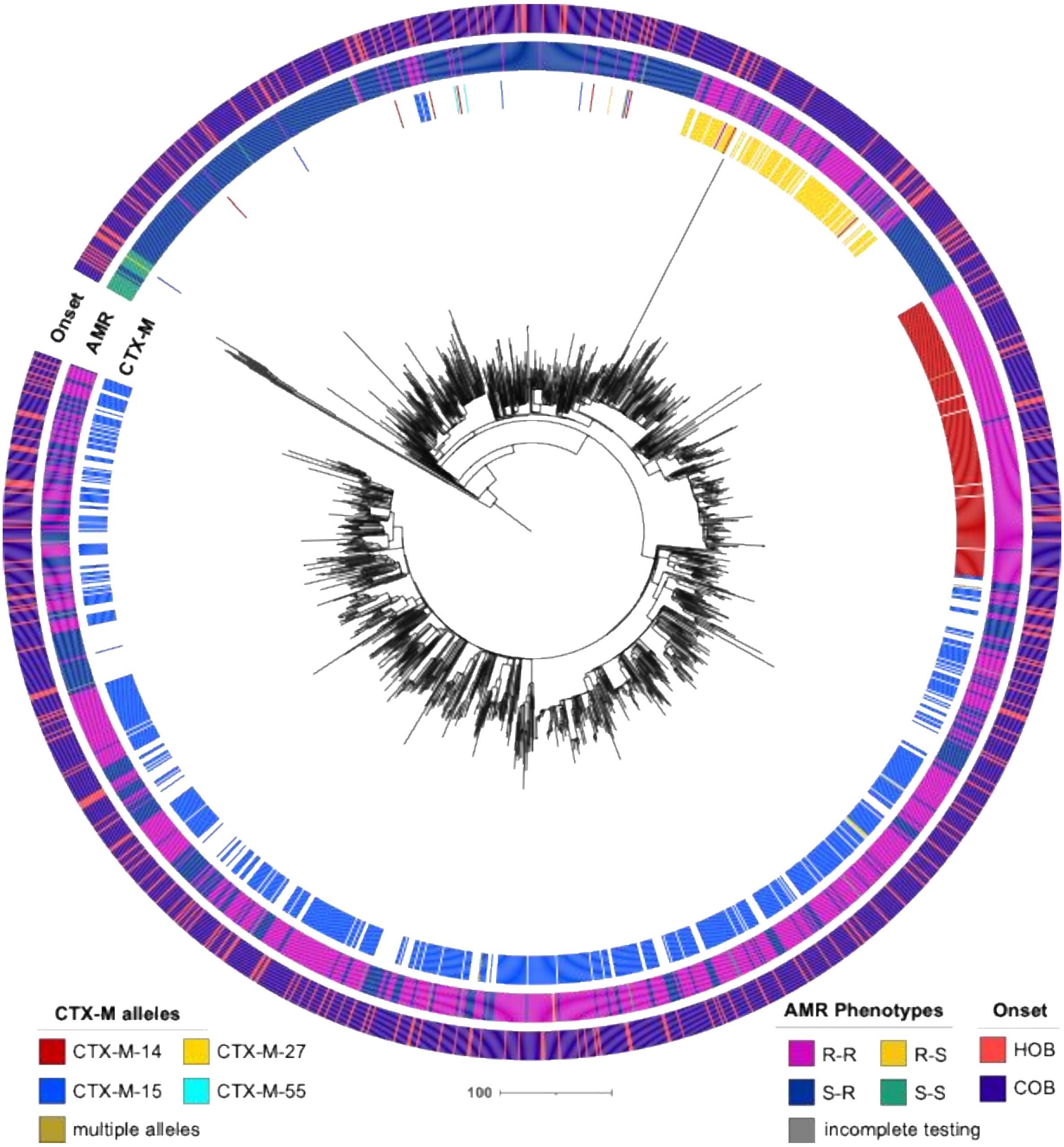

**Figure S11. Phylogenetic tree of *Escherichia coli* ST131 Clade C. Recombination-removed whole-genome phylogenetic tree of all isolates assigned to *E. coli* StrainGST cluster 1, which represents sequence type (ST) 131 Clade C. Inner ring shows presence of CTX-M alleles. Middle ring shows AMR phenotypes (ceftriaxone-ciprofloxacin). Outer ring shows hospital-onset bacteremia (HOB) vs. community-onset bacteremia (COB).**

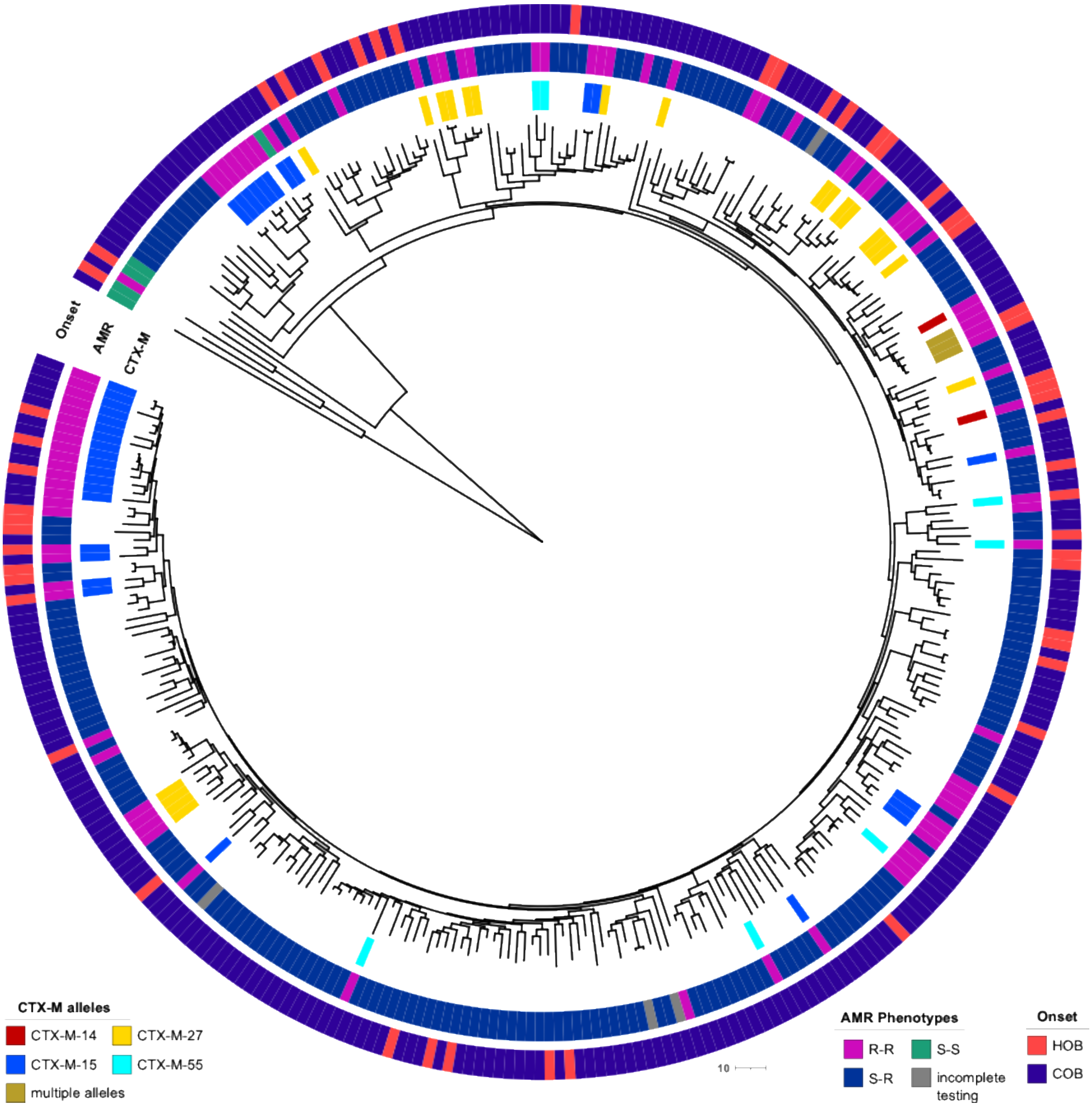

**Figure S12. Phylogenetic tree of *Escherichia coli* ST1193 lineage. Recombination-removed whole-genome phylogenetic tree of all isolates assigned to *E. coli* StrainGST cluster 7, which represents sequence type (ST) 1193. Inner ring shows presence of CTX-M alleles. Middle ring shows AMR phenotypes (ceftriaxone-ciprofloxacin). Outer ring shows hospital-onset (HOB) vs community-onset bacteremia (COB).**

#### *S4.5.5 Klebsiella pneumoniae*

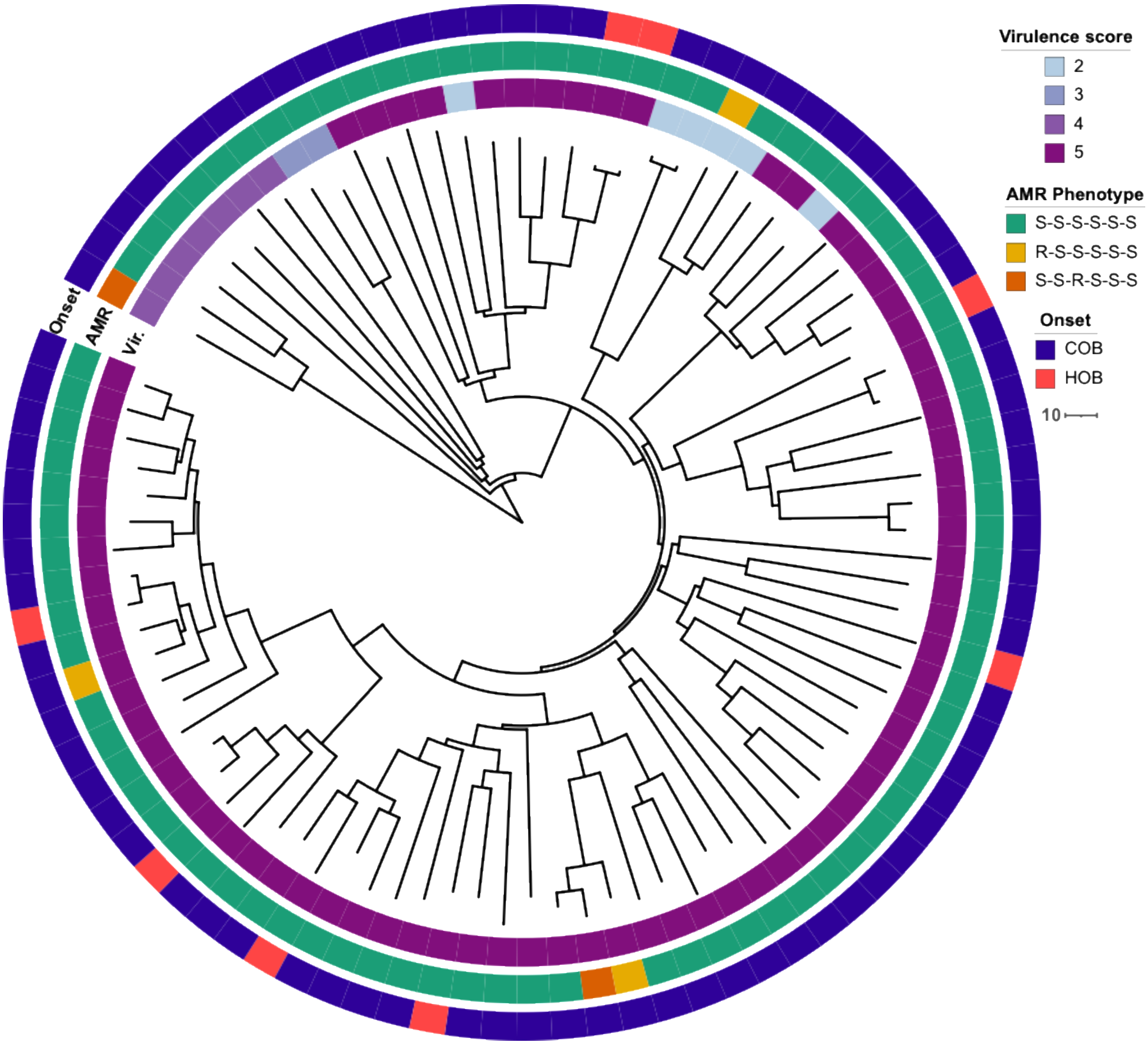

**Figure S 13. Phylogenetic tree of *Klebsiella pneumoniae* ST23 lineage. Recombination-removed whole-genome phylogenetic tree of all isolates assigned to *K. pneumonaie* StrainGST cluster 1, which represents sequence type (ST) 23. Inner ring shows virulence score as predicted by Kleborate. Middle ring shows AMR phenotypes. Outer ring shows hospital-onset bacteremia (HOB) vs. community-onset bacteremia (COB).**

### S5 Time trend analyses

#### S5.1 Overall time trend stratified by onset

**Table S13. Time trend estimates for bacteremia incidence stratified by onset for five target organisms. Estimates for the average annual percentage change (AAPC), 95% confidence intervals, and p-values are given.**

| Organism | Time period | AAPC Estimate (%) | AAPC 95% CI (%) | p-value |
|---|---|---|---|---|
| Community-onset | | | | |
| Total | 2006-2018 | 4.5 | (4.0, 5.0) | < 0.0001 |
| | 2019-2022 | -10.3 | (-12.5, -8.0) | < 0.0001 |
| *Staphylococcus aureus* | 2006-2017 | 4.8 | (3.7, 5.8) | < 0.0001 |
| | 2018-2022 | -4.1 | (-7.1, -1.0) | < 0.0001 |
| *Enterococcus faecalis* | 2006-2022 | 2.8 | (1.4, 4.2) | < 0.0001 |
| *Enterococcus faecium* | 2006-2011 | 17.7 | (1.2, 36.8) | 0.0342 |
| | 2012-2022 | -1 | (-5.1, 3.3) | 0.0307 |
| *Klebsiella pneumoniae* | 2006-2022 | 4.8 | (3.6, 6.0) | < 0.0001 |
| *Escherichia coli* | 2006-2018 | 4.3 | (3.6, 5.0) | < 0.0001 |
| | 2019-2022 | -7.6 | (-10.6, -4.4) | < 0.0001 |
| Hospital-onset | | | | |
| Total | 2006-2022 | -1.5 | ( -2.1, -0.8) | < 0.0001 |
| *Staphylococcus aureus* | 2006-2010 | -9.8 | (-14.7, -4.6) | 0.0003 |
| | 2011-2022 | 0.1 | (-1.3, 1.6) | 0.0004 |
| *Enterococcus faecalis* | 2006-2022 | -1.8 | (-3.6, 0.1) | 0.0631 |
| *Enterococcus faecium* | 2006-2012 | 2.9 | (-4.0, 10.4) | 0.4125 |
| | 2013-2022 | -5.1 | (-8.6, -1.4) | 0.0454 |
| *Klebsiella pneumoniae* | 2006-2022 | 2 | (0.2, 3.8) | 0.0321 |
| *Escherichia coli* | 2006-2022 | -0.3 | (-1.2, 0.7) | 0.6125 |

#### S5.2 Phenotype incidence time trends

**Table S14. Phenotypic incidence trends for five target bacterial species.** Time trends were estimated using segmented Poisson regression. p-values < 0.0001 are indicated by a * and otherwise reported up to four decimals.

| Organism/Phenotype | **Incidence time trend estimates**<br>Average Annual Percentage Change (95% confidence interval) | | | |
|---|---|---|---|---|
| | **Community-onset** | | **Hospital-onset** | |
| ***Staphylococcus aureus***<br>Phenotype: Cloxacillin – Ciprofloxacin – Erythromycin - Clindamycin | | | | |
| S-S-S-S<br>N = 4542 | 2006 - 2018 | 2019 - 2022 | 2006 - 2008 | 2009 - 2022 |
| | - 5.7%<br>(4.3%, 7.1%)* | -3.9<br>(-7.6, -0.1)* | -26.9<br>(-37.9, -14.0)<br>p-value = 0.0002 | -1.8<br>(0.2, 3.3)* |
| R-R-R-R<br>N = 518 | -10.2%<br>(-13.2%, -7.1%)* | | -16.0%<br>(-19.3%, -12.7%)* | |
| ***Enterococcus faecalis***<br>Phenotype: Tetracycline – Gentamicin - Ciprofloxacin | | | | |
| R-S-S<br>N = 748 | - 3.6% (1.6%, 5.6%)<br>p-value = 0.0003 | | 0.1% (-2.3%, 2.6%) | |
| S-S-S<br>N = 352 | - 3.2% (0.5%, 6.1%)<br>p-value = 0.0209 | | 0.9% (-2.8%, 4.8%)<br>p-value = 0.6487 | |
| R-R-S, N = 165<br>(< 2013, >=2013) | -0.03 (-4.5, 4.7) | | 2006 - 2012 | 2013-2022 |
| | | | - 30.2%<br>(2.3%, 65.7%)*<br>p-value = 0.0189 | -11.6%<br>(-21.2%, -0.9%)<br>p-value = 0.0053 |
| R-R-R, N = 157 | -5.3% (-9.5%, -0.9%)<br>p-value = 0.0196 | | -11.3%<br>(-15.7%, -6.6%)* | |
| ***Enterococcus faecium*** | | | | |
| vancomycin susceptible<br>N = 581 | - 3.8% (1%, 6.7%)<br>p-value = 0.0076 | | -3.1% (-5.3%, -1.0%)<br>p-value = 0.0042 | |
| vancomycin resistant<br>N = 219 | 2006-2012 | 2013-2022 | 2006-2012 | 2013-2022 |
| | 41.1%<br>(-9.9%, 121.1%) | -5.4%<br>(-16.3%, 6.8%) | - 36.1%<br>(7.2%, 73.0%)<br>p-value = 0.0115 | -8.9%<br>(-14.7%, -2.6%)<br>p-value = 0.0062 |
| ***Escherichia coli***<br>Phenotype: Cefazolin – Ceftriaxone – Ciprofloxacin – Gentamicin – Trimethoprim/Sulfamethoxazole | | | | |
| S-S-S-S-S<br>N = 3625 | 2006 - 2018 | 2019 - 2022 | 0.7% (-1.1%, 2.6%)<br>p-value = 0.4548 | |
| | 2.8%<br>(1.7%, 4.0%)* | -5.5%<br>(-11.0%, 0.0%)<br>p-value = 0.0062 | | |
| R-R-R-R-R<br>N = 418 | 2006 - 2013 | 2014 - 2022 | 2006 - 2014 | 2015 - 2022 |
| | - 29.7%<br>(14.6%, 46.8%)* | -5.6%<br>(-9.6%, -1.5%)* | 14.9%<br>(-1.1%, 33.6%) | -9.9%<br>(-18.6%, -0.3%)<br>p-value = 0.0085 |
| Ceftriaxone-Ciprofloxacin-resistant<br>N = 1341 | 2006 - 2013 | 2014 - 2022 | 2006 - 2014 | 2015-2022 |
| | - 25.0%<br>(16.6%, 34.0%)* | 1.6%<br>(-0.7%, 4.1%) | - 16.7%<br>(5.5%, 29.2%)<br>p-value = 0.0028 | -3.6%<br>(-8.9%, 2.1) |

| | | |
|---|---|---|
| ***Klebsiella pneumoniae***<br>Phenotype: Cefazolin – Ceftriaxone – Ciprofloxacin – Gentamicin - Piperacillin/Tazobactam - Trimethoprim/Sulfamethoxazole | | |
| S-S-S-S-S-S<br>N = 1686 | 3.7% (2.3%, 5.2%)* | 0.2% (-2.1%, 2.5%)<br>p-value = 0.8582 |
| Ceftriaxone-Ciprofloxacin-resistant<br>N = 62 | - 15.7% (2.1%, 31.3%)<br>p-value = 0.0228 | - 15.9% (1.9%, 31.9%)<br>p-value = 0.0251 |

* p-value < 0.0001

#### S5.3 Strain-phenotype incidence time trends for Gram-positive organisms

**Table S15. Time trend estimates for strain-phenotype incidence for Gram-positive species over the study period Jan 1, 2006-Oct 14, 2022.** Time trends were estimated using segmented Poisson regression. p-values < 0.0001 are indicated by a star(*) and otherwise reported up to four decimals.

| **Organism/ Most prevalent strains and phenotypes** | **Incidence time trend estimates**<br>Average Annual Percentage Change (95% confidence interval) | | |
|---|---|---|---|
| | **Community-onset** | | **Hospital-onset** |
| ***Staphylococcus aureus***<br>Phenotype: Cloxacillin – Ciprofloxacin – Erythromycin - Clindamycin | | | |
| **CC30**<br>S-S-S-S<br>N = 882 | 2006 - 2016 | 2017 - 2022 | -6.9% (-10.0%, -3.7%)* |
| | - 4.7%<br>(1.0%, 8.5%)<br>p-value = 0.0117 | -12.8%<br>(-18.1%, -7.1%)* | |
| **CC5** | | | |
| S-S-S-S,<br>N = 417 | 2006 - 2019 | 2020 - 2022 | -1.2% (-4.8%, 2.7%)<br>p-value = 0.5473 |
| | - 8.4%<br>(4.8%, 12.1%)* | -28.1%<br>(-47.1%, -2.3%)<br>p-value = 0.0091 | |
| R-R-R-R,<br>N = 414 | -13.0%<br>(-16.6%, -9.2%)* | | -18.6% (-22.6%, -14.4%)* |
| **CC8** | | | |
| S-S-S-S,<br>N = 258 | 2006 - 2019 | 2020 - 2022 | 1.1% (-3.9%, 6.3%)<br>p-value = 0.6806 |
| | - 11.7%<br>(6.7%, 17.0%)* | -50.4%<br>(-70.7%, -16.2%)<br>p-value = 0.0025 | |
| R-R-R-S<br>N = 380 | -3.0% (-6.0%, 0.1%)<br>p-value = 0.0576 | | -6.4% (-11.7%, -0.7%)<br>p-value = 0.0276 |
| **CC45** | | | |
| S-S-S-S<br>N = 507 | 2006 - 2018 | 2019 - 2022 | -3.0% (-6.0%, 0.2%)<br>p-value = 0.0686 |
| | - 7.6%<br>(4.0%, 11.4%)* | -13.4%<br>(-28.0%, 4.1%)<br>p-value = 0.0225 | |
| S-S-R-R<br>N = 130 | 2006 - 2018 | 2019 - 2022 | 5.5% (-1.4%, 12.9%)<br>p-value = 0.1210 |
| | 6.9%<br>(-0.5%, 14.9%)<br>p-value = 0.0689 | -27.5%<br>(-46.2%, -2.1%)<br>p-value = 0.0135 | |

| | | | | |
|---|---|---|---|---|
| **CC15**<br>S-S-S-S<br>N = 568 | 2006 - 2019 | 2020 - 2022 | 0.7 (-2.7%, 4.2%)<br>p-value = 0.6777 | |
| | - 7.0%<br>(4.1%, 9.9%)* | ¯ -34.4%<br>(-51.0%, -12.1%)<br>p-value = 0.0011 | | |
| **CC97**<br>S-S-S-S<br>N = 330 | 2006 - 2015 | 2016 - 2022 | - 6.2% (1.4%, 11.3%)<br>p-value = 0.0109 | |
| | - 22.1%<br>(13.5%, 31.4%)* | -9.9%<br>(-17.4%, -1.6%)* | | |
| ***Enterococcus faecalis***<br>Phenotype: Tetracycline – Gentamicin - Ciprofloxacin | | | | |
| **ST179** | | | | |
| R-S-S<br>N = 207 | - 8.2% (3.3%, 13.2%)<br>p-value = 0.0008 | | 4.2% (-0.7%, 9.3%)<br>p-value = 0.0924 | |
| R-R-S<br>N = 98 | -6.6% (-16.7%, 4.7%)<br>p-value = 0.2410 | | 2006-2014 | 2015-2022 |
| | | | 13.2%<br>(-2.7%, 31.7%)<br>p-value = 0.1080 | -28.1%<br>(-44.1%, -7.4%)<br>p-value = 0.0025 |
| **ST40** (R-S-S)<br>N = 103 | -4.4% (-9.2%, 0.7%)<br>p-value = 0.0886 | | -2.9% (-9.5%, 4.1)<br>p-value = 0.4019 | |
| **ST16** (R-S-S)<br>N = 74 | 2.3% (-4.1%, 9.1%)<br>p-value = 0.4920 | | -4.5% (-11.4%, 3.0%)<br>p-value = 0.2360 | |
| **ST6** (R-R-R)<br>N = 47 | -5.4% (-12.4%, 2.0%)<br>p-value = 0.1502 | | -5.8% (-15.5%, 5.0%)<br>p-value = 0.2788 | |
| **ST103** (R-R-R)<br>N = 36 | -12.5% (-34.6%, 17.2)<br>p-value = 0.3717 | | -17.7% (-27.2%, -7.0%)<br>p-value = 0.0017 | |
| **ST778** (R-R-R)<br>**N=22** | No statistical time trend analysis due to small sample size | | | |
| ***Enterococcus faecium*** | | | | |
| **ST117**<br>(vancomycin-resistant)<br>N = 114 | -<br>Numbers too small | | 2006 - 2012 | 2013 - 2022 |
| | | | - 36.7%<br>(3.3%, 80.9%)<br>p-value = 0.0286 | -26.9%<br>(-37.0%, -15.3%)<br>p-value = 0.0001 |

#### S5.4 Strain-phenotype incidence time trends for Gram-negative organisms

**Table S16. Time trend estimates for strain-phenotype incidence for Gram-negative species over the study period Jan 1, 2006 - Oct 14, 2022. Time trends were estimated using segmented Poisson regression.**

<table>
<tr><th rowspan="2">Organism/<br>Most prevalent strains and phenotypes</th><th colspan="4">Incidence time trends<br>Average Annual Percentage Change (95% confidence interval)</th></tr>
<tr><th colspan="2">Community-onset</th><th colspan="2">Hospital-onset</th></tr>
<tr><td colspan="5">Escherichia coli<br>Phenotype: Ceftriaxone – Ciprofloxacin</td></tr>
<tr><td colspan="5">ST131</td></tr>
<tr><td rowspan="2">R-R<br>N = 909</td><td>2006 - 2011</td><td>2012 - 2022</td><td>2006-2014</td><td>2015-2022</td></tr>
<tr><td>- 36.8%<br>(21.7%, 53.8%)*</td><td>0.6%<br>(-1.9%, 3.1%)</td><td>- 15.4%<br>(4.3%, 27.7%)<br>p-value = 0.0055</td><td>4.9%<br>(-12.7%, 3.5%)</td></tr>
<tr><td rowspan="2">S-R<br>N = 778</td><td>2006 - 2013</td><td>2014 - 2022</td><td colspan="2" rowspan="2">-2.6%<br>(-6.1%, 1.1%)</td></tr>
<tr><td>14.8%<br>(8.4%, 21.6%)*</td><td>-2.6%<br>(-6.5%, 0.8%)</td></tr>
<tr><td rowspan="2">S-S<br>N = 341</td><td>2006 - 2018</td><td>2019 - 2022</td><td colspan="2" rowspan="2">-0.6%<br>(-6.4%, 5.5%)</td></tr>
<tr><td>- 4.5%<br>(0.9%, 8.2%)<br>p-value = 0.0135</td><td>-21.6%<br>(-37.8%, -1.1%)<br>p-value = 0.0164</td></tr>
<tr><td>ST95<br>S-S<br>N = 1281</td><td colspan="2">-0.7%<br>(-2.0%, 0.7%)</td><td colspan="2">-3.2%<br>(-7.0%, 0.7%)</td></tr>
<tr><td>ST73<br>S-S<br>N = 969</td><td colspan="2">-0.1%<br>(-1.7%, 1.5%)</td><td colspan="2">-4.0%<br>(-7.5%, -0.4%)<br>p-value = 0.0291</td></tr>
<tr><td rowspan="2">ST69<br>S-S<br>N = 619</td><td>2006 - 2018</td><td>2019 - 2022</td><td colspan="2" rowspan="2">-4.5%<br>(-8.6%, -0.3%)<br>p-value = 0.0364</td></tr>
<tr><td>- 3.0%<br>(0.3%, 5.8%)<br>p-value = 0.0262</td><td>-14.5%<br>(-27.3%, 0.5%)</td></tr>
<tr><td rowspan="2">ST127<br>S-S<br>N = 352</td><td>2006 - 2017</td><td>2018 - 2022</td><td colspan="2" rowspan="2">-3.5%<br>(-8.6%, 1.8%)</td></tr>
<tr><td>2.6%<br>(-1.4%, 6.8%)</td><td>-13.8%<br>(-26.0%, 0.3%)</td></tr>
<tr><td colspan="5">ST1193</td></tr>
<tr><td rowspan="2">S-R<br>N = 221</td><td>2006 - 2019</td><td>2020 - 2022</td><td colspan="2" rowspan="2">-<br>(numbers too small)</td></tr>
<tr><td>- 17.4%<br>(11.1%, 24.0%)*</td><td>-29.8%<br>(-49.6%, -2.1%)*</td></tr>
<tr><td>R-R<br>N = 87</td><td colspan="2">- 17.7%<br>(9.0%, 30.0%)*</td><td colspan="2">-<br>(numbers too small)</td></tr>
<tr><td colspan="5">Klebsiella pneumoniae (2006-2019)<br>Phenotype: Ceftriaxone – Ciprofloxacin</td></tr>
<tr><td>ST23<br>S-S<br>N = 86</td><td colspan="2">2.3%<br>(-4.4%, 9.4%)</td><td colspan="2">-<br>(numbers too small)</td></tr>
</table>

* p-value < 0.0001

### S6 Additional analyses

#### S6.1 Time trend analyses for susceptible phenotypes in *Staphylococcus aureus*

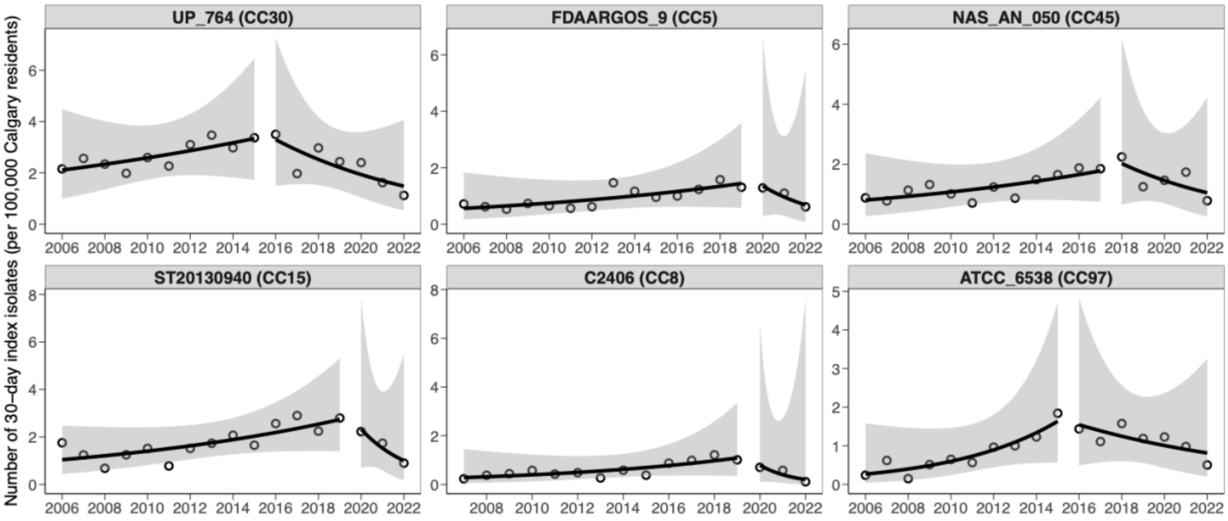

**Figure S14. Incidence of *S aureus* bacteremia for susceptible phenotypes stratified by major StrainGST sequence clusters. Points represent observed 30-day index isolates per 100,000 Calgary residents/patient days. Lines and shaded areas represent rates and 95% confidence intervals estimated by segmented regssion.**

#### S6.2 Association of *S aureus* bacteremia incidence with patient characteristics

We evaluated whether temporal trends in *S aureus* community-onset bacteremia incidence differed according to patient characteristics, such as biological sex, age groups, substance-related disorders, and resistance phenotype.

##### S6.2.1 Association with biological sex

Generally, (community-onset) incidence varied by biological sex (Figure S15) and phenotype. To assess whether increasing trends were associated with a specific patient population, a multivariable negative-binomial log-linear model was fitted to the annual number of *S aureus* community-onset bacteremia episodes, with the log of the Calgary population size and the number of patient-days as offset. Fixed effects included calendar year (continuous), biological sex (reference = female), phenotype of the index isolate, and all two- and three-way interaction terms (Year × Sex × Phenotype). The fully susceptible phenotype (S-S-S-S) served as the phenotype reference category.

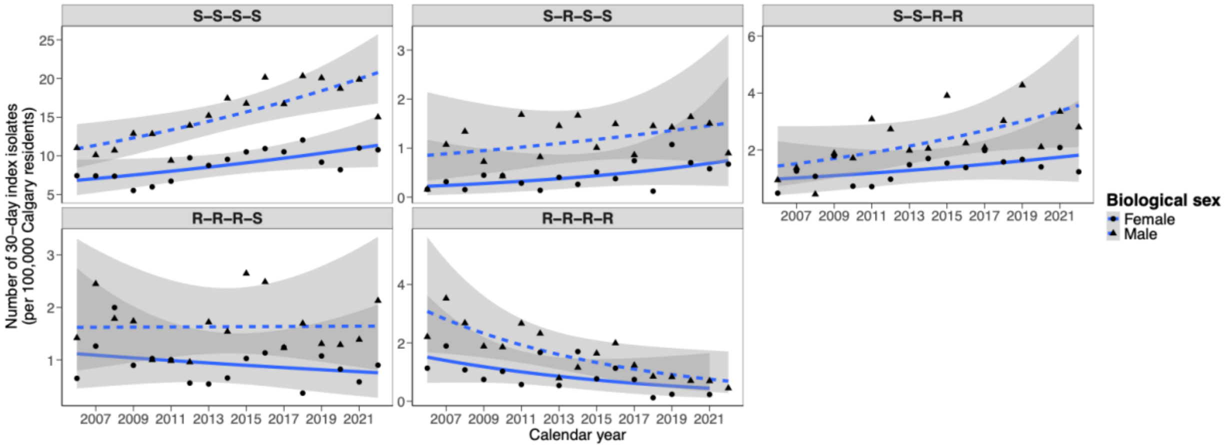

**Figure S15. Community-onset bacteremia incidence rates for *S aureus* stratified by phenotypes and biological sex.** Each panel shows the annual incidence (per 100,000 person-years) of *S aureus* bacteremia with a specific resistance phenotype, from 2007 to 2021. Lines represent fitted values from negative binomial regression models stratified by biological sex (female: solid line and circle, male: dashed line and triangle), with shaded areas indicating 95% confidence intervals. Resistance (R) or susceptibility (S) to four antibiotics (cloxacillin, ciprofloxacin, erythromycin, and clindamycin) is denoted in phenotype labels (e.g., S–S–S–S).

The global Wald test for the four Year × Sex × Phenotype coefficients showed no evidence that the sex-specific time trend differed by phenotype ($\chi^2 = 3.2$, df = 5, p = 0.6648). Point estimates for these interaction slopes were all close to the null, and their 95 % confidence intervals spanned unity (Table S2). These results indicate that the year-to-year changes in *S aureus* community-onset bacteremia incidence were similar in males and females across all resistance phenotypes and that it appears to be driven by factors other than the biological sex.

**Table S17. Sex-specific temporal trends in *Staphylococcus aureus* bacteremia incidence, stratified by and resistance phenotype.** Annual male-to-female incidence-rate ratios (IRR) per calendar year are shown for each resistance phenotype within community-onset (blood culture obtained ≤ 48 h after admission). Estimates derive from negative-binomial log-linear models that included calendar year (continuous), biological sex, phenotype, and all two- and three-way interaction terms, with the log of the population at risk as an offset. The fully susceptible phenotype (S-S-S-S) served as the reference and therefore is not listed. Values are the exponentiated Year × Sex × Phenotype coefficients, accompanied by Wald 95 % confidence intervals, z-statistics, and p-values. An IRR > 1 indicates that incidence in males is increasing faster (or decreasing more slowly) over time than in females for the specified phenotype, whereas an IRR < 1 indicates the opposite.

| Onset (48h) | Phenotype | Annual IRR* | 95% CI† | z-statistic | p-value |
|---|---|---|---|---|---|
| Community-onset | S-R-S-S | 0.95 | (0.88, 1.02) | -1.427 | 0.154 |
| | S-S-R-R | 1.009 | (0.96, 1.06) | 0.375 | 0.708 |
| | R-R-R-S | 1.016 | (0.96, 1.07) | 0.578 | 0.563 |
| | R-R-R-R | 0.981 | (0.92, 1.04) | -0.614 | 0.539 |

*IRR = Incidence rate ratio (males vs females)
†Confidence interval

##### S6.2.2 Association with age groups

Similar to our previous analysis, we fit a multivariable negative-binomial log-linear to the annual number of *S aureus* bacteremia episodes, with the log of the Calgary population size and the number of patient-days as offset for community-onset and hospital-onset bacteremia, respectively. Fixed effects included calendar year (continuous), age group (reference =00-09 age group), phenotype of the index isolate, and all two- and three-way interaction terms (Year × Age group × Phenotype). The fully susceptible phenotype (S-S-S-S) served as the phenotype reference category.

Point estimates for these interaction slopes were all close to the null, and their 95 % confidence intervals spanned unity.

##### S6.2.3 Association with substance-related disorders

To assess whether increasing trends in the S-S-S-S phenotype is associated with patients with substance-related disorders (SRD), we performed a multivariable negative-binomial log-linear model fitted to the annual number of S aureus community-onset bacteremia episodes. Fixed effects included calendar year (continuous), SRD indicator (reference = no SRD), phenotype of the index isolate, and all two- and three-way interaction terms (Year × Sex × Phenotype). The fully susceptible phenotype (S-S-S-S) served as the phenotype reference category. Patients with SRD were identified using diagnostic codes. In Table S18, we summarize the relevant diagnostic categories, including substance induced mental disorders, dependence, non-dependent use, poisoning events, and laboratory findings, that were used to identify patients with substance-related conditions in the dataset. Patients who had any of these diagnostic codes were classified as "At least one SRD diagnostic code" whereas patients who didn't as "No SRD".

The vast majority of patients were classified as patient without SRD (Figure S16). The global Wald test for the four Year × Drug_use × Phenotype coefficients showed no evidence that the sex-specific time trend differed by phenotype ($\chi^2 = 6.19$, df = 4, p = 0.1855). Point estimates for these interaction slopes were all close to the null, and their 95 % confidence intervals spanned unity (Table S19). These results indicate that the year-to-year changes in *S aureus* community-onset bacteremia incidence were similar in patients with and without substance-related disorders across all resistance phenotypes and that it appears to be driven by other factors.

**Table S18. Diagnostic codes for substance-related disorders and poisonings, as classified in ICD-9-CM and ICD-10-CM.**

| Code or Range | Description |
|---|---|
| 2920 | Drug-induced delirium |
| 29211 | Amphetamine intoxication delirium |
| 29212 | Cocaine intoxication delirium |
| 2922 | Drug-induced persisting dementia |
| 29281–29285 | Drug-induced psychotic disorders: amphetamine (81), cocaine (82), hallucinogens (83), opioids (84), phencyclidine (85) |
| 29289 | Other drug-induced mental disorders |
| 2929 | Unspecified drug-induced mental disorder |
| 30400–30403 | Opioid dependence: unspecified (00), continuous (01), episodic (02), in remission (03) |
| 30420–30423 | Cannabis dependence: unspecified (20), continuous (21), episodic (22), in remission (23) |
| 30440–30443 | Sedative, hypnotic, or anxiolytic dependence: unspecified (40), continuous (41), episodic (42), in remission (43) |
| 30460–30463 | Cocaine dependence: unspecified (60), continuous (61), episodic (62), in remission (63) |
| 30470–30473 | Amphetamine dependence: unspecified (70), continuous (71), episodic (72), in remission (73) |
| 30480–30483 | Hallucinogen dependence: unspecified (80), continuous (81), episodic (82), in remission (83) |
| 30490–30493 | Other/unspecified drug dependence: unspecified (90), continuous (91), episodic (92), in remission (93) |
| 30550–30553 | Opioid abuse: unspecified (50), continuous (51), episodic (52), in remission (53) |
| 30560–30563 | Sedative, hypnotic, or anxiolytic abuse: unspecified (60), continuous (61), episodic (62), in remission (63) |
| 30570–30573 | Amphetamine abuse: unspecified (70), continuous (71), episodic (72), in remission (73) |
| 30580–30583 | Hallucinogen abuse: unspecified (80), continuous (81), episodic (82), in remission (83) |
| 30590–30593 | Other/unspecified drug abuse: unspecified (90), continuous (91), episodic (92), in remission (93) |
| E8500 | Accidental poisoning by heroin |
| 9650 | Poisoning by opiates and related narcotics |
| 970 | Poisoning by alcohol |
| F11.0–F11.9 | Opioid-related disorders: use through other conditions/remission |
| F12.0–F12.9 | Cannabis-related disorders |
| F14.0–F14.9 | Cocaine-related disorders |
| F15.0–F15.9 | Other stimulant-related disorders |
| F19.0–F19.9 | Other psychoactive substance-related disorders |
| Z7151 | Opioid abuse counseling |
| Z7152 | Cocaine abuse counseling |
| Z722 | Drug use behavior |
| R781–R783 | Finding of opioids, narcotics, or hallucinogens in blood |
| R78.50–R78.59 | Findings of benzodiazepines in specimen |
| R78.6–R78.7 | Findings of antidepressants |
| R78.9 | Finding of other drugs, unspecified |
| T400–T404, T406 | Poisoning by narcotics: opium (T400), heroin (T401), methadone (T402), other (T403–T404), unspecified (T406) |

| | |
|---|---|
| T405 | Poisoning by cocaine |
| T408 | Poisoning by other psychoactive substances |
| T409 | Poisoning by unspecified narcotics |

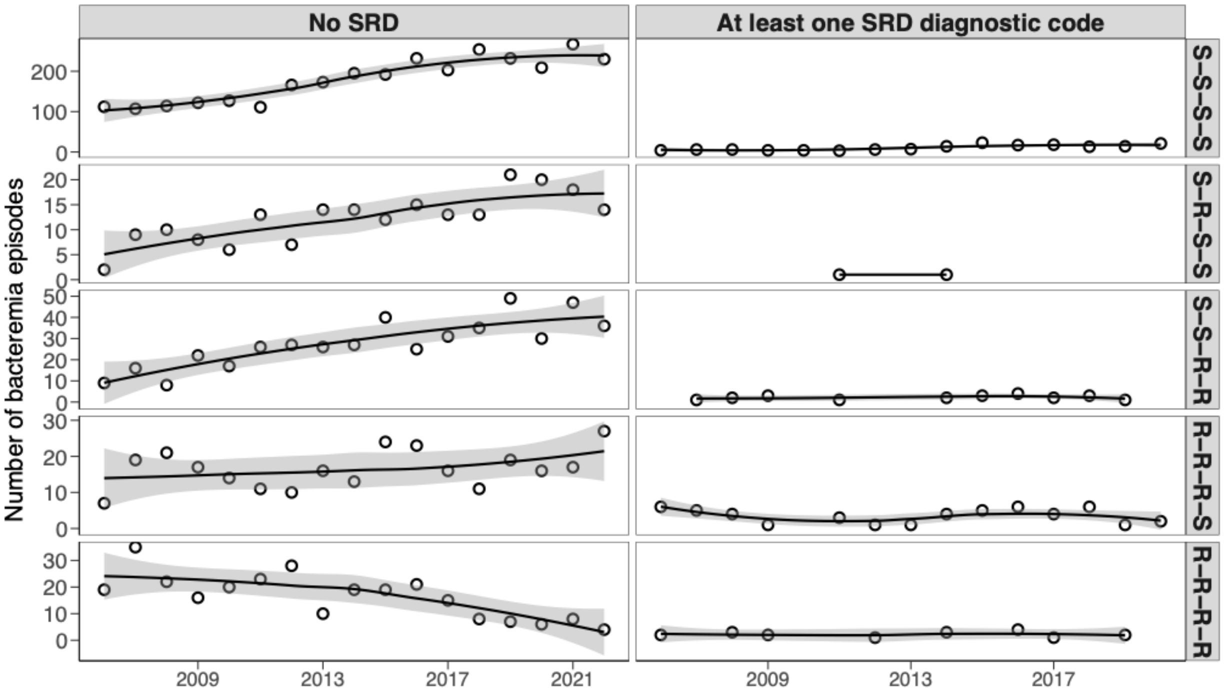

**Figure S16. Annual number of community-onset *Staphylococcus aureus* bacteremia by resistance phenotype and substance-related disorder (SRD) status, Calgary 2006–2022.** Each facet shows one resistance phenotype – SRD combination with calendar year on the x-axis and number of episodes on the y-axis. Points denote the absolute number of bacteremia episodes; lines are fitted lines using locally estimated scatterplot smoothing (loess); grey bands are corresponding 95% confidence intervals.

**Table S19. Substance-related disorder-specific temporal trends in *Staphylococcus aureus* bacteremia incidence, stratified by resistance phenotype.** Annual incidence-rate ratios (IRR) for patients with vs without substance-related disorder (SRD) per calendar year are shown for each resistance phenotype within community-onset (blood culture obtained ≤ 48 h after admission). Estimates derive from negative-binomial log-linear models that included calendar year (continuous), substance-related disorder indicator, phenotype, and all two- and three-way interaction terms. The fully susceptible phenotype (S-S-S-S) served as the reference and therefore is not listed. Values are the exponentiated Year × Sex × Phenotype coefficients, accompanied by Wald 95 % confidence intervals, z-statistics, and p-values. An IRR > 1 indicates that incidence in patients with SRD is increasing faster (or decreasing more slowly) over time than in patients without SRD for the specified phenotype, whereas an IRR < 1 indicates the opposite.

| Onset (48h) | Phenotype | Annual IRR* | 95% CI† | z-statistic | p-value |
|---|---|---|---|---|---|
| Community-onset | S-R-S-S | 0.874 | (0.30, 2.57) | -0.285 | 0.775 |
| | S-S-R-R | 0.886 | (0.79, 1.00) | -2.022 | 0.043 |
| | R-R-R-S | 0.926 | (0.84, 1.01) | -1.656 | 0.098 |
| | R-R-R-R | 1.008 | (0.89, 1.14) | 0.122 | 0.903 |

*IRR = Incidence rate ratio (males vs females)
†Confidence interval

#### S6.2.4 Association with short-term migration

Our data indicate that the vast majority of incident *Staphylococcus aureus* bacteremia cases (~92%, N = 6689/7271) occur within non-short-term-migratory resident populations. Inward short-term migration accounted for less than 5% of total cases during the fiscal year of their blood culture. These findings suggest that phenotypic temporal trends in S. aureus bacteremia incidence are not associated with short-term migration.

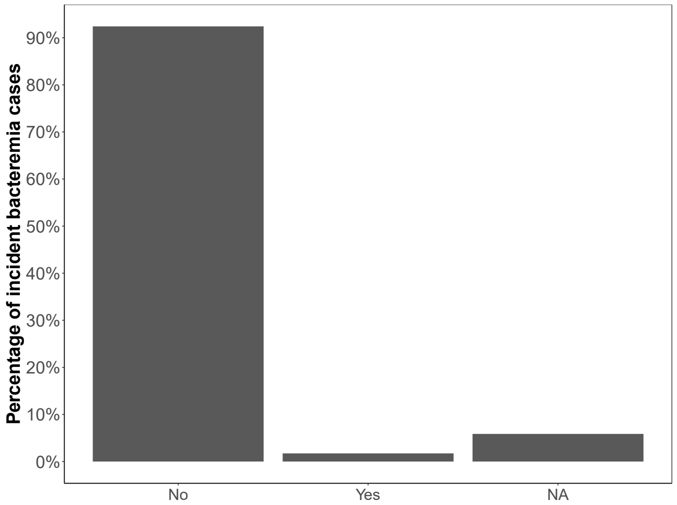

**Figure S17. Percentage of Incident *Staphylococcus aureus* Bacteraemia Cases by Inward Short-Term Migration Status.** This bar chart illustrates the percentage of incident *S aureus* bacteraemia cases involving patients who engaged in short-term migration into the province within the fiscal year of their blood culture.

#### S6.3 Association of *E faecalis* bacteremia incidence with patient characteristics

To determine whether there is a difference in age distributions between ST40 and ST179, we performed a statistical test for testing the null hypothesis that the age proportions are the same. We did not have the statistical power to determine whether incidence trends were driven by certain age groups for the *E faecalis* bacteremia dataset.

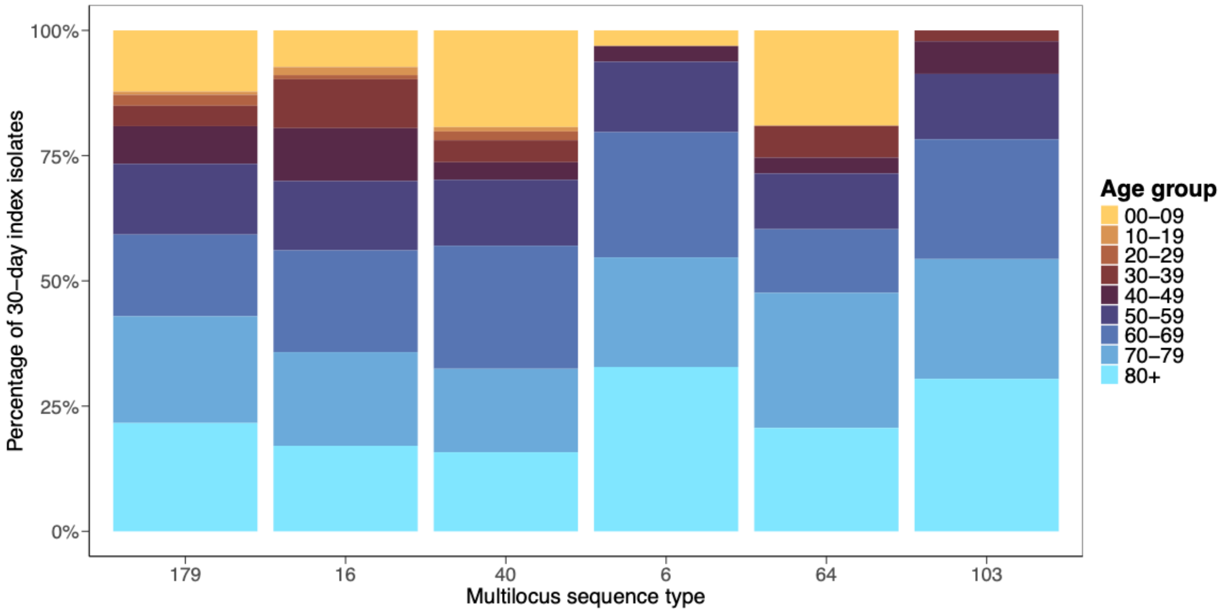

**Figure S18. Age distribution for six most common *E faecalis* multilocus sequence types. Colours represent different age groups.**

#### S6.4 Shannon Diversity index

##### *S6.4.1* Shannon Diversity index for five target pathogens

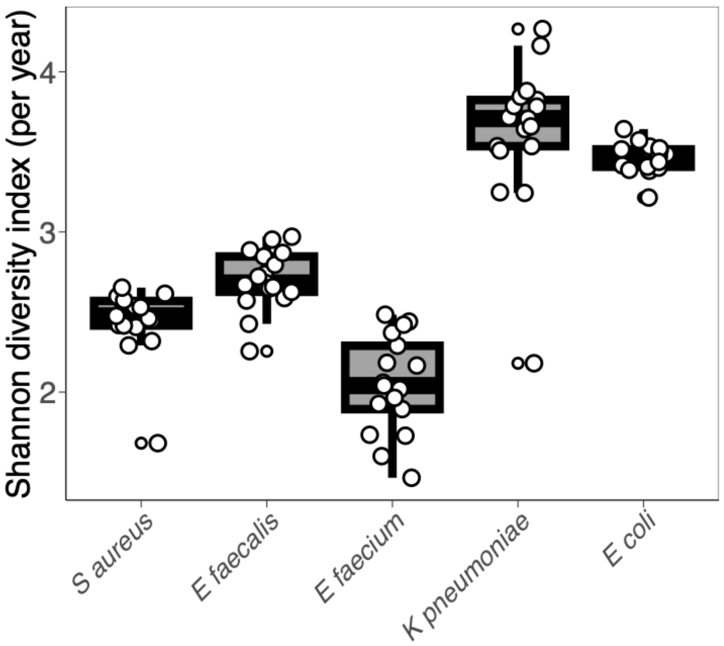

**Figure S19. Shannon diversity index for five target bacterial pathogens. Boxplot of distribution of Shannon diversity index based on the number of StrainGST reference cluster per organism. Points represent the Shannon diversity index per species per year.**

##### *S6.4.2* Diversity index for *E faecalis* antibiograms

**Table S20. Shannon diversity index for *E faecalis* bacteremia cases stratified by antibiogram.**

| Antibiogram | Shannon diversity index |
|---|---|
| R-S-S | 2.66 |
| S-S-S | 4.04 |
| R-R-S | 0.84 |
| R-R-R | 1.91 |
| R-S-R | 2.77 |
| S-S-R | 2.28 |
| S-R-R | 1.63 |

#### S6.5 Analysis of Calgary and international K pneumoniae isolates encoding CTX-M-15

Antimicrobial resistant *K pneumoniae* are often described as local or global problem clones.[15] To differentiate between acquisition of CTX-M-15 by previously susceptible strains endemic to Calgary and importation of resistant *K pneumoniae* strains, we compared the genomic diversity of *K pneumoniae* encoding CTX-M-15 in our dataset to all *K pneumoniae* data available in NCBI's Sequencing Read Archive as of January 16, 2024 with linked paired-end Illumina reads with geographic location and collection date metadata. All data was downloaded and assigned a StrainGST cluster with the same reference database used for Calgary isolates.[16] We additionally used AMRFinderPlus results produced by the AlltheBacteria[17] project to identify the presence or absence of CTX-M-15.[14] Of 44,624 *K. pneumoniae* isolates analysed, 17,702 (39.7%) encoded CTX-M-15. The majority of Calgary isolates were part of StrainGST clusters that commonly encoded CTX-M-15 in the global dataset (Table S13). One cluster (55) only contained two isolates encoding CTX-M-15 in the global dataset. However, phylogenetic analysis using ParSNP[18] suggests that CTX-M-15 encoding isolates in Calgary are more closely related to global CTX-M-15 encoding isolates from Asia than to susceptible isolates from the same cluster in Calgary (Figure S17). While CTX-M-15-encoding isolates in Calgary were in the same clusters as globally sampled isolates, not all global problem clones were sampled in Calgary. For example, despite 14.5% (2558/17,702) of CTX-M-15-encoding isolates in the global dataset belonging to cluster 31 (ST147), no Calgary isolates encoding CTX-M-15 belonged to this StrainGST cluster.

**Table S21. CTX-M-15 encoding *K pneumoniae* in Calgary and global dataset.**

| StrainGST cluster | Most common ST of CTX-M-15 positive isolates | Number of CTX-M-15 positive isolates from Calgary | Number of CTX-M-15 negative isolates from Calgary | Number of CTX-M-15 positive isolates in Global Dataset |
|---|---|---|---|---|
| 34 | 307 | 9 | 3 | 2311 (13%) |
| 7 | 45 | 6 | 32 | 444 (2.5%) |
| 15 | 11 | 4 | 18 | 1869 (11%) |
| 39 | 25 | 3 | 8 | 56 (0.32%) |
| 55 | 592 | 3 | 5 | 2 (0.011%) |
| 16 | 405 | 3 | 17 | 219 (1.2%) |

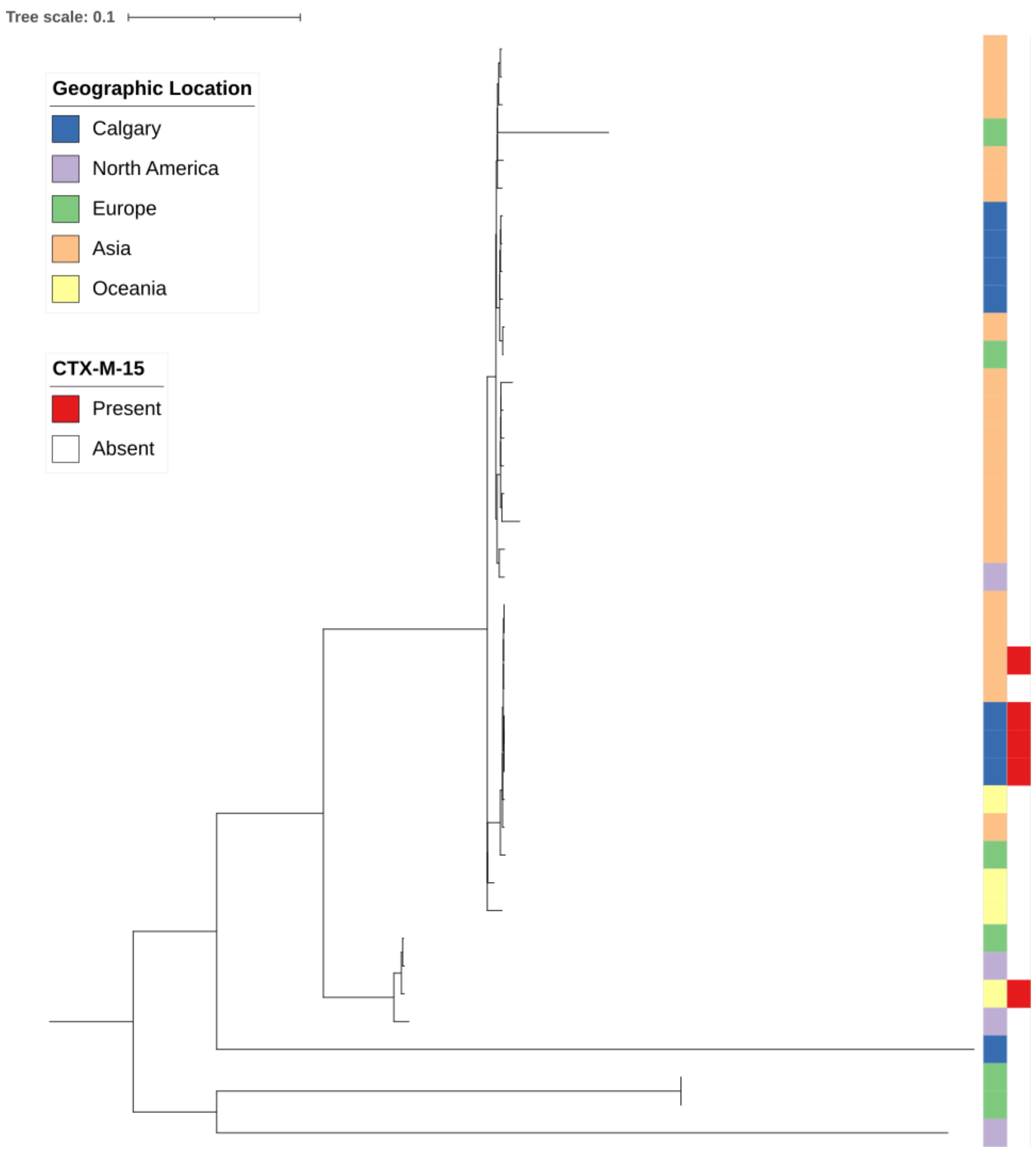

**Figure S20. Phylogeny of cluster 55 isolates from Calgary and global dataset. Whole genome alignment and maximum likelihood phylogeny of all isolates assigned to StrainGST cluster 55 were generated with ParSNP. Tips are annotated with geographic location: Calgary (blue, isolates sequenced in this study), North America (purple), Europe (green), Asia (orange), Oceania (yellow). Tips representing isolates encoding CTX-M-15 are annotated with red.**

#### S6.6 Fluoroquinolone resistance in *Escherichia coli*

##### S6.6.1 Fluoroquinolone outpatient antibiotic use in *E coli* bacteremia patients

While overall community- and inpatient fluoroquinolone prescribing declined (see main text Figure 5), the rate of outpatient fluoroquinolone prescribing in *Escherichia coli* bacteremia patients increased from 2006 to 2017.

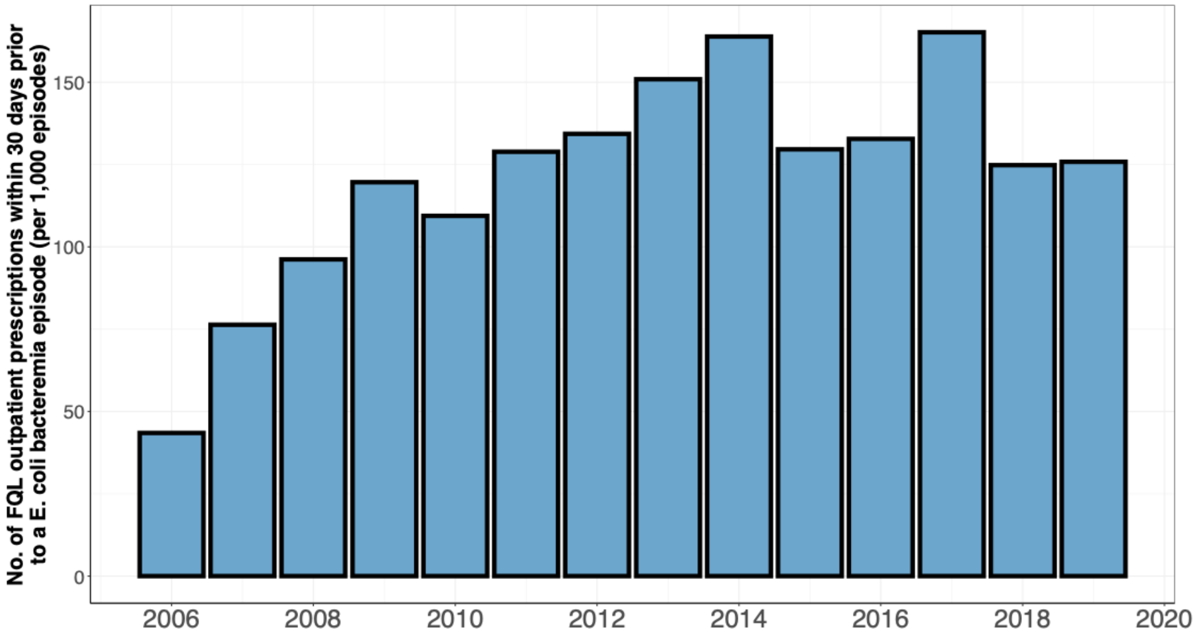

**Figure S21. Temporal trends in outpatient fluoroquinolone prescribing in *E coli* bacteremia patients, Jan 7, 2006–Dec 31, 2019. Each bar represents the number of fluoroquinolone prescriptions dispensed in the 30 days preceding an *E coli* bacteremia episode, standardized per 1,000 bacteremia episodes.**

##### S6.6.2 Association of prior antibiotic use with resistance phenotype

We conducted a logistic regression analysis of Escherichia coli bacteremia isolates to identify factors associated with ciprofloxacin resistance (0 = susceptible, 1 = intermediate or resistant). The primary exposure was fluoroquinolone prescribing, measured as either annual inpatient prescribing rates or outpatient exposure in the 30 days preceding the bacteremia episode (number of fluoroquinolone prescriptions dispensed in the outpatient setting). The model was adjusted for year, age, biological sex, Charlson Weighted Index of Comorbidities, onset of infection (hospital- vs. community-onset), episode number, and co-resistance to other antibiotics (ampicillin, cefazolin, ceftriaxone, and trimethoprim-sulfamethoxazole).

Analyses were performed using Bayesian logistic regression implemented in the brms package in R version 4.4.0. Four Markov chain Monte Carlo (MCMC) chains were run with 10,000 iterations per chain, including 1,000 warm-up iterations. We used adapt_delta = 0.9 and max_treedepth = 15 to ensure stable sampling. Posterior distributions were used to estimate adjusted log odds ratios and 95% credible intervals.

Recent outpatient fluoroquinolone exposure was strongly associated with ciprofloxacin resistance (adjusted odds ratio = 1.5, 95% credibility interval: 1.0, 1.9), while inpatient fluoroquinolone prescribing showed little to no association. Co-resistance to other agents, particularly ampicillin, cefazolin, and ceftriaxone, was also associated with higher odds of ciprofloxacin resistance. Other patient-level factors, such as age, sex, comorbidity burden, infection onset, and episode number, had smaller effect sizes with confidence intervals crossing the null. These findings indicate that recent outpatient fluoroquinolone use is a key driver of ciprofloxacin resistance among E. coli bacteremia cases in our setting (Figure S22).

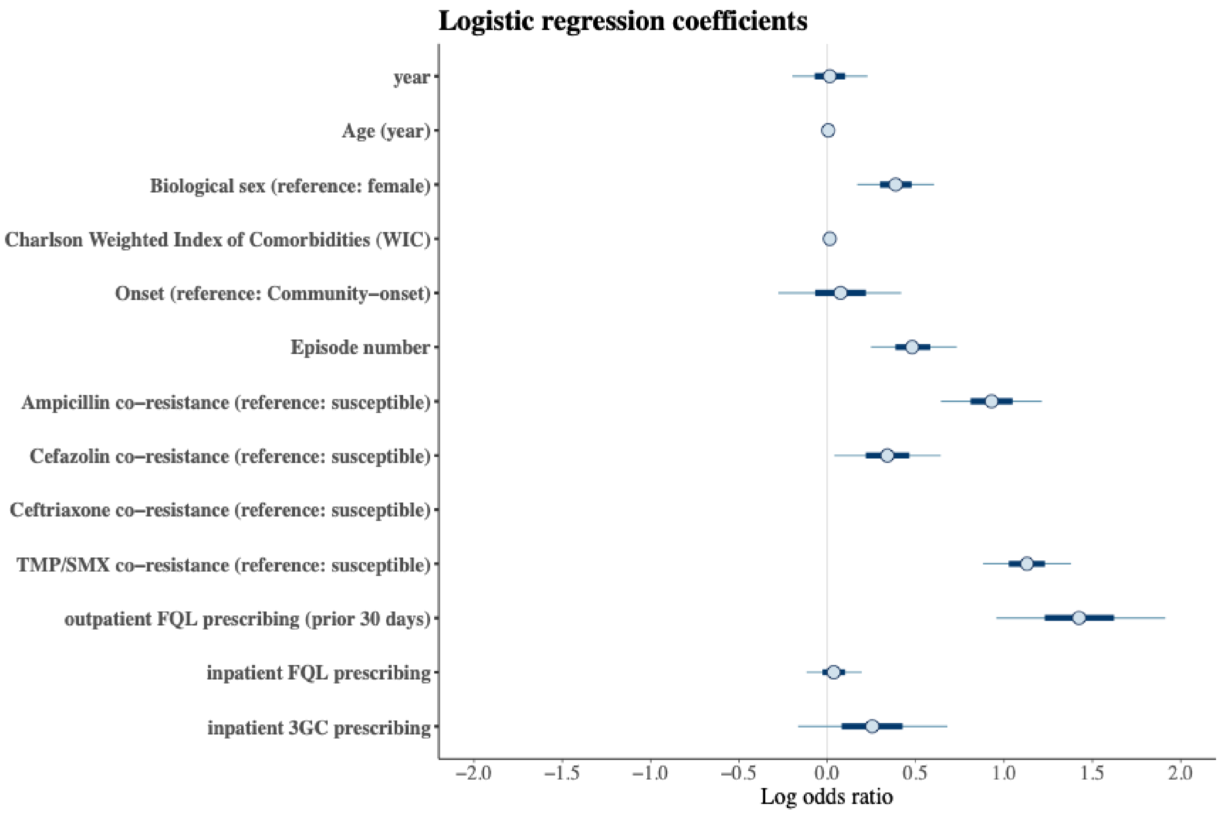

**Figure S22. Logistic regression results for ciprofloxacin resistance in *E coli* bacteremia patients. The log odds ratio for covariates is presented on the y-axis. The Covariates are given on the y-axis. Points represent the estimates, thick blue bars the 80% and thin black bars the 95% credibility intervals. FQL = fluoroquinolone, 3GC = third-generation cephalosporins, TMP/SMX = Trimethoprim/Sulfamethoxazole**

#### S6.7 MIC distributions for ceftazidime and ceftriaxone among *E coli* ST131 bacteremia

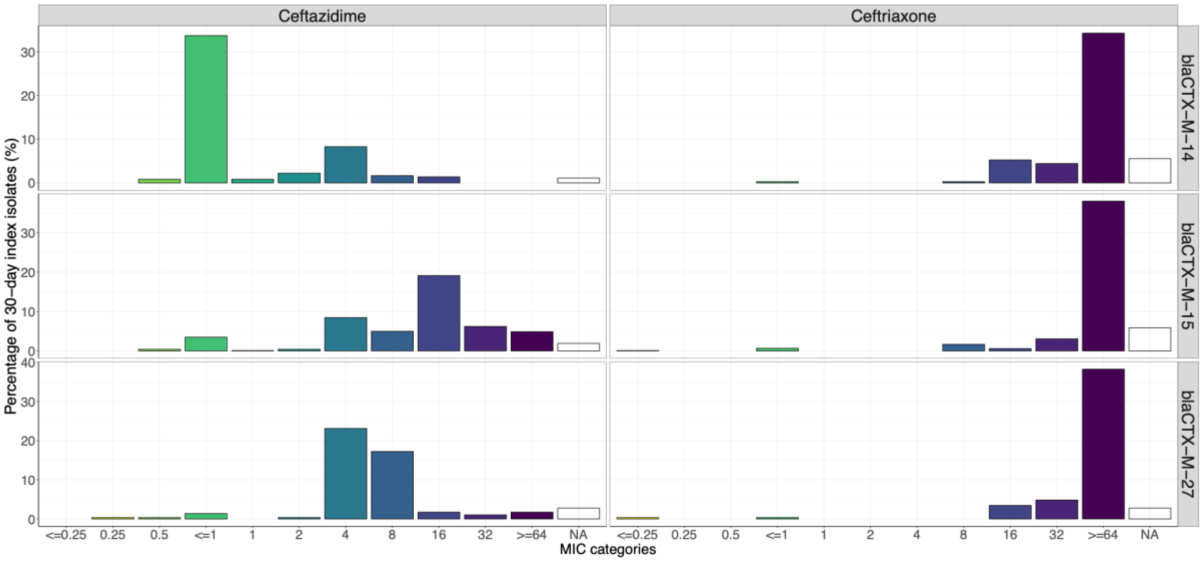

**Figure S23. Minimum inhibitory concentration (MIC) distributions for ceftazidime and ceftriaxone among *Escherichia coli* ST131 bacteremia isolates, stratified by the most common CTX-M β-lactamase variants (CTX-M-14, CTX-M-15, and CTX-M-27). MIC values are expressed in μg/mL. Each panel shows the percentage of isolates within each MIC category. Isolates with missing susceptibility results are labeled as "NA".**

#### S6.8 Carbapenem prescribing and resistance in Enterobacterales species

Carbapenem resistance remained rare in both *K pneumoniae* and *E coli* (Figure S24-23).

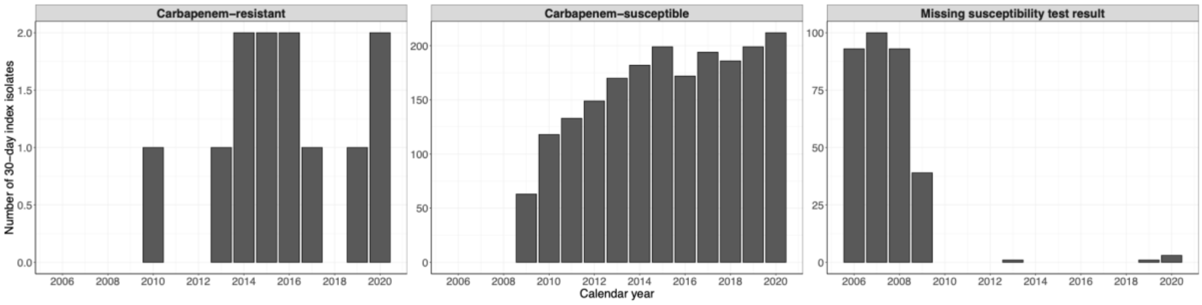

**Figure S24. Carbapenem resistance for *Klebsiella pneumoniae,* Jan 1, 2006 - Dec 31, 2022.** Bars indicate the total number of 30-day index isolates classified as carbapenem-resistant (intermediate or resistant), carbapenem-susceptible, or with missing susceptibility test results.

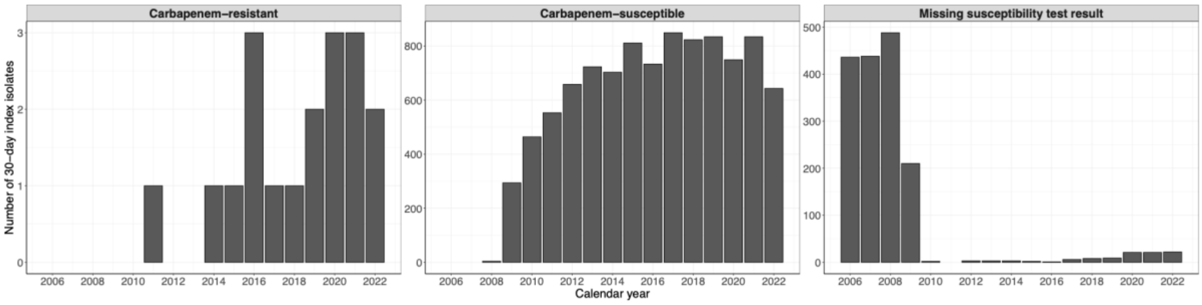

**Figure S25. Carbapenem resistance for *Escherichia coli,* Jan 1, 2006 - Oct 4, 2022. Bars indicate the total number of 30-day index isolates classified as carbapenem-resistant (intermediate or resistant), carbapenem-susceptible, or with missing susceptibility test results.**

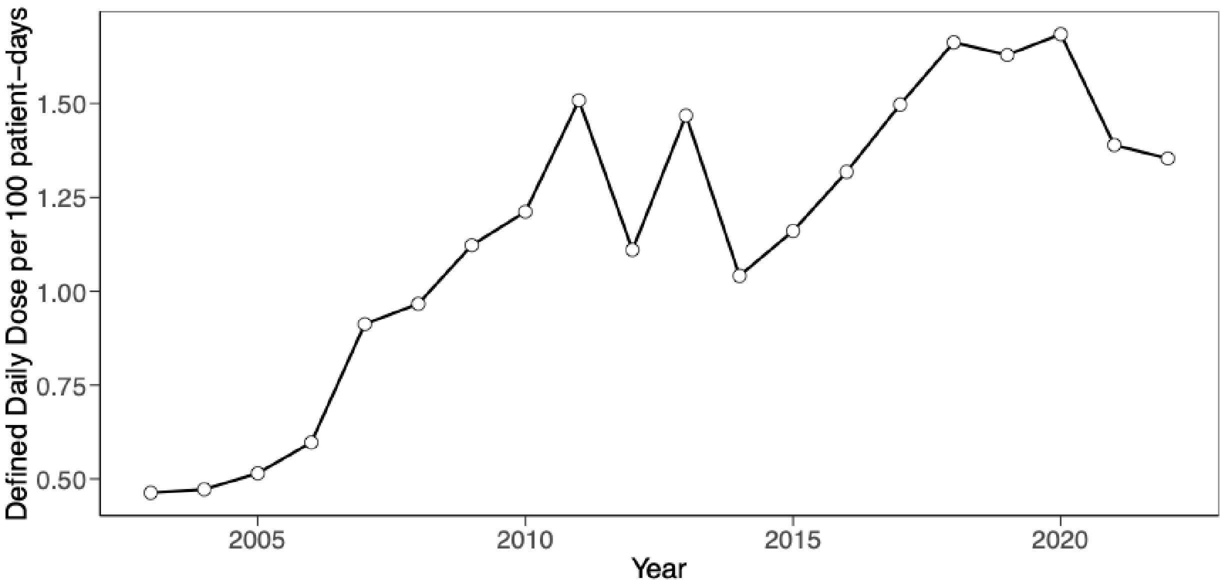

**Figure S26. Inpatient carbapenem prescribing rates in hospitals of the Calgary Health Zone, 2004-2022. Points represent defined daily dose (DDD) per 100 patient-days.**

5 Alberta Health Services. Infection Prevention and Control Hand Hygiene Program. 2023.

6 A Provincial Antimicrobial Stewardship Program: Successes and Challenges in Alberta – National Collaborating Centre for Infectious Diseases. https://nccid.ca/publications/provincial-antimicrobial-stewardship-program-successes-challenges-alberta/ (accessed March 8, 2026).

7 Muggeo VMR. Estimating regression models with unknown break-points. *Stat Med* 2003; **22**: 3055–71.

8 Muggeo V. Segmented: An R Package to Fit Regression Models With Broken-Line Relationships. *R News* 2008; **8/1**. https://www.researchgate.net/publication/234092680_Segmented_An_R_Package_to_Fit_Regression_Models_With_Broken-Line_Relationships (accessed Dec 1, 2024).

9 Shannon CE. A mathematical theory of communication. *The Bell System Technical Journal* 1948; **27**: 379–423.

10 Bankevich A, Nurk S, Antipov D, *et al.* SPAdes: A new genome assembly algorithm and its applications to single-cell sequencing. *Journal of Computational Biology* 2012; **19**: 455–77.

11 Walker BJ, Abeel T, Shea T, *et al.* Pilon: An Integrated Tool for Comprehensive Microbial Variant Detection and Genome Assembly Improvement. *PLoS One* 2014; **9**: e112963.

12 Kim D, Song L, Breitwieser FP, Salzberg SL. Centrifuge: Rapid and sensitive classification of metagenomic sequences. *Genome Res* 2016; **26**: 1721–9.

13 Hunt M, Mather AE, Sánchez-Busó L, *et al.* ARIBA: Rapid antimicrobial resistance genotyping directly from sequencing reads. *Microb Genom* 2017; **3**: e000131.

14 Feldgarden M, Brover V, Gonzalez-Escalona N, *et al.* AMRFinderPlus and the Reference Gene Catalog facilitate examination of the genomic links among antimicrobial resistance, stress response, and virulence. *Scientific Reports 2021 11:1* 2021; **11**: 1–9.

15 Wyres KL, Lam MMC, Holt KE. Population genomics of Klebsiella pneumoniae. *Nature Reviews Microbiology 2020 18:6* 2020; **18**: 344–59.

16 van Dijk LR, Walker BJ, Straub TJ, *et al.* StrainGE: a toolkit to track and characterize low-abundance strains in complex microbial communities. *Genome Biol* 2022; **23**: 1–27.

17 Hunt M, Lima L, Anderson D, *et al.* AllTheBacteria - all bacterial genomes assembled, available and searchable. *bioRxiv* 2024; : 2024.03.08.584059.

18 Kille B, Nute MG, Huang V, Kim E, Phillippy AM, Treangen TJ. Parsnp 2.0: scalable core-genome alignment for massive microbial datasets. *Bioinformatics* 2024; **40**. DOI:10.1093/BIOINFORMATICS/BTAE311.
